# The demographic and geographic impact of the COVID pandemic in Bulgaria and Eastern Europe in 2020

**DOI:** 10.1101/2021.04.06.21254958

**Authors:** Antoni Rangachev, Georgi K. Marinov, Mladen Mladenov

## Abstract

**Background:** The COVID-19 pandemic followed a unique trajectory in Eastern Europe compared to other heavily affected regions, with most countries there only experiencing a major surge of cases and deaths towards the end of 2020 after a relatively uneventful first half of the year. However, the consequences of that surge have not received as much attention as the situation in Western countries. Bulgaria, even though it has been one of the most heavily affected countries, has been one of those neglected cases.

**Methods:** We use mortality and mobility data from Eurostat, official governmental and other sources to examine the development and impact of the COVID-19 pandemic in Bulgaria and other European countries.

**Results:** We find a very high level of excess mortality in Eastern European countries measured by several metrics including excess mortality rate (EMR), P-scores and potential years of life lost. By the last metric Eastern Europe emerges as the hardest hit region by the pandemic in Europe in 2020. With a record EMR at ∼0.25% and a strikingly large and mostly unique to it mortality rate in the working age demographics, Bulgaria emerges as one of the most affected countries in Eastern Europe. The high excess mortality in Bulgaria correlates with insufficient intensity of testing and with delayed imposition of “lockdown” measures. We also find major geographic and demographic disparities within the country, with considerably lower mortality observed in major cities relative to more remote areas (likely due to disparities in the availability of medical resources). Analysis of the course of the epidemic revealed that individual mobility measures were predictive of the eventual decline in cases and deaths. However, while mobility declined as a result of the imposition of a lockdown, it already trended downwards before such measures were introduced, which resulted in a reduction of deaths independent of the effect of restrictions.

**Conclusions:** Large excess mortality and high numbers of potential years of life lost are observed as a result of the COVID pandemic in Bulgaria, as well as in several other countries in Eastern Europe. Significant delays in the imposition of stringent mobility-reducing measures combined with a lack of medical resources likely caused a substantial loss of life, including in the working age population.

## Introduction

The SARS-CoV-2 virus and COVID-19, the disease it causes ^1–3^, have emerged as the most acute public health emergency in a century. The novel coronavirus spread rapidly before significant efforts at containment were implemented in much of the world, resulting in devastating early outbreaks in the United States and Western Europe, starting in late February and early March of 2020.

Some combination of lockdown measures, imposed in response to surging infections, voluntary changes in behavior, and the onset of the summer season is thought to have caused the major decline in COVID-19 cases in Europe in the summer of 2020. However, winter in the Southern hemisphere, during which large epidemics developed in South Africa and South America, together with the well-documented seasonality of common-cold coronaviruses ^4^, strongly suggested that a major second wave was to be expected in Europe with the arrival of winter ^5^, and when it eventually arrived expectation turned into reality.

During the early months of the pandemic, a dichotomy emerged between countries in Western and Eastern Europe (with the possible exception of Russia). Western Europe was heavily affected - by June 2020 official COVID mortality reached 600 to 800 deaths per million (DPM) in countries such as Spain, Italy, the UK, Belgium, France, and Sweden, with excess mortality rates even higher ^6–10^. In contrast, most Eastern European countries registered relatively few deaths, possibly because of much earlier implementation of social distancing measures relative to the development of the outbreak.

This dichotomy disappeared during the second wave at the end of 2020, with both countries in Western and Eastern Europe officially registering a large number of COVID-related fatalities, as well as in some cases considerably larger excess mortality. However, the development of the pandemic in Eastern Europe has so far generally received much less attention than that in the West even though multiple countries in the region were heavily affected by it. We show this using multiple excess mortality measures, which quantify the pandemic-related loss of life and allow for standardized comparisons between countries.

Among Eastern European countries, Bulgaria has emerged as perhaps the most heavily affected by the pandemic as suggested by excess mortality analysis ^6^. Here we analyze the development and impact of the pandemic on Bulgaria, in the broader European context, across demographic groups within the country, and for its regional subdivisions, as well as the influence of human mobility changes and government-imposed quarantine measure on the course of the pandemic. We use these analyses to identify correlate factors likely responsible for particularly high unexplained excess mortality in certain settings.

## Methodology

### Data Sources

All-cause mortality data for European countries, as well as NUTS-3 (Nomenclature of Territorial Units for Statistics) regions in Bulgaria, was obtained from Eurostat ^11,12^. The data presented in the datasets is sex- and age-stratified, with age groups split in increments of 5 years. Since not all countries submit data at the same time and in the same manner, only countries that have consistent weekly data for the period 2015-2020 (inclusive) were analyzed.

Country-level population data at the beginning of 2020 was collected through Eurostat ^13^, but was further supplemented by population data from the United Nations’ UN-data Data Service ^14^. We further elaborate on this topic in the subsequent section on Potential Years of Life Lost (PYLL) and Working Years of Life Lost (WYLL) estimates.

Life expectancy values at different ages were obtained from three separate sources. We acquire the full life tables for Bulgaria through the country’s National Statistical Institute ^15^, and for Czechia through the country’s Statistical Office ^16^. Abridged life tables for all European countries were obtained from the World Health Organization’s open data platform ^17^. This dataset is partitioned by age, in increments of 5 years.

COVID-related mortality and testing data for Bulgaria was collected through the resources available from the Ministry of Health ^18,19^. COVID-related mortality for Czechia was acquired from Czech Ministry of Health official website tracking the pandemic ^20^.

### Data Availability

All datasets and associated code can be found at https://github.com/Mlad-en/COV-BG.git.

### Excess mortality and P-scores

To calculate excess mortality across countries as well as across Bulgarian regions, we analyze the mortality observed between week 10 and 53 of 2020 and compare it to the mean mortality between the same period for the previous five years (2015–2019). We derive this mean both on a weekly and aggregate total basis. We then establish a 95% confidence interval for this mean. This range is used to calculate excess mortality as:

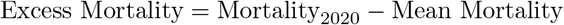

This calculation is done both as a sex- and age-stratified metric, as well as an aggregated total. To normalize excess mortality across countries, we calculate excess mortality per total population. To do this, we use population data from Eurostat for 2020.

Based on the excess mortality ranges we also compute a P-score value for each country/region. A P-score value is defined as “the percentage difference between the number of deaths in 2020[…] and the average number of deaths in the same period-week or month over the years 2015–2019.” ^21^

We calculate the P-score as follows:

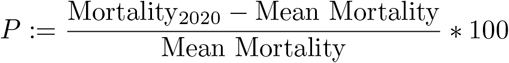

We also calculate the ratio between excess mortality and official COVID-19-attributed mortality. Due to the demonstrably low testing in Bulgaria ^22^ and other countries, this allows us to estimate under-reported COVID-19 fatalities. We also use the total positive tests per region reported at the end of 2020 to compute a Case Fatality Ratio (CFR) which estimates the proportion of COVID-19 fatalities among confirmed cases.

### Potential Years of Life Lost (PYLL), Aged-Standardized Years of life lost Rate (ASYR), and Working Years of Life Lost (WYLL) estimates

Potential Years of Life Lost (PYLL) is a metric that estimates the burden of disease on a given population by looking at premature mortality. It is derived as the difference between a person’s age at the time died and the expected years of life for people at that age in a given country. As such, the metric attributes more weight to people that have died at a younger age.

We compute the PYLL across countries by taking the positive all-cause excess mortality for all ages groups (in Eurostat they are aggregated at 5 year intervals). We use the abridged life expectancy tables by the WHO (also aggregated at 5 year intervals) and calculate a total and average PYLL value for all countries. To be more precise, for an age interval [*x, x* + 4] and sex *s* (if no sex is specified we assume it’s for both sexes) define by ED([*x, x* + 4], *s*) the excess deaths and by LE([*x, x* + 4], *s*) the life expectancy. Then the potential years of life lost are computed as

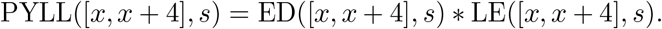

The total PYLL is computed by summing over all age intervals. In our computations we take into account the margin of error for each ED([*x, x* + 4], *s*).

A limitation on this approach is the upper-boundary aggregation value for the two datasets. The all-cause mortality dataset’s upper boundary is 90+, while the WHO’s abridged life tables only go up to the 85+ age bracket. To account for this, we attribute the life expectancy of the 85+ age group to the 85-89 mortality group. We have further excluded the 90+ mortality group from our analysis. This is further elaborated on in the Limitations subsection, where we also provide a way of correcting for this exclusion.

Two countries for which we have the exact ages and sex for each reported COVID-19 fatality are Bulgaria and Czechia. We also have full life tables (increments of one year) for both countries provided by their respective statistical institutes. This allows us to compute and compare the PYLLs for each country based on excess mortality data and official data for COVID-19 fatalities.

Finally, we standardize PYLL values across countries by diving the total sum value by the population and normalizing it per 100,000 people:

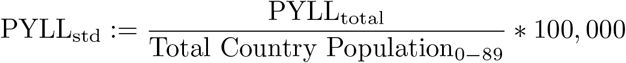

The data for country-level populations in Eurostat has a similar limitation in the upper boundary of the age distribution (a cut-off at 85+). To mitigate this limitation, we supplement the population data from Eurostat for ages 0-84 with population size data for the 85-89 age group from the UNdata Data Service.

To compare the impact of the pandemic across European populations with different age structures we compute the Age-Standardized Years of Life Lost Rate (ASYR) ^23,24^. Let ([*x, x* + 4], *s*) be an age interval for a sex *s* in a standard life expectancy table for a given population. Denote by *P* ([*x, x* + 4], *s*) the population size of ([*x, x* + 4], *s*). Define the PYLL rate for ([*x, x* + 4], *s*) as

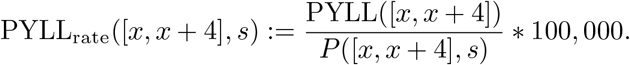

For the 2013 European Standard Population (ESP) denote by *W* ([*x, x*+4], *s*) the weight of ([*x, x*+4], *s*) in the standard population. Define

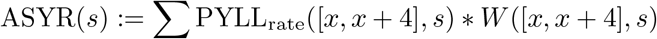

where the sum is taken over all age intervals. For a given population of sex *s* this measure is interpreted as the years of life lost per 100, 000 people (of sex *s*) if the population has the same age distribution as the ESP. ASYR allows for comparison of the pandemic impact on EU countries having different age distributions. Finally, we derive total, average and total standardized WYLL value approximations. To accomplish this, we first assume a common retirement age at 65 and thus exclude excess mortality for all age groups over 65. To calculate the remaining years of working life, we further assume a mean age for each age group, e.g. for the age interval 60-64 we assume a mean age at 62.5 years. This would leave this group with approximately 2.5 years until retirement. Limitations on this approach are discussed in the subsequent Limitations subsection.

### Stringency index and Mobility data

Metrics of population mobility were obtained from the Google COVID-19 Community Mobility Reports ^25^. These datasets contain data on how visits and length of stay at different places change compared to a baseline by generating anonymized metrics from data of Google users who have switched on “Location History” on their mobile devices.

To quantify governmental pandemic-response measures across countries, we used the Oxford COVID-19 Government Response Tracker ^26^, which systematically collects information on several different common policy responses that governments have taken to mitigate the effects of the pandemic ^27^. This allows a comparison of governmental measures between over 180 countries worldwide.

## Limitations

Each of the presented data sources and approaches to analysis have their own limitations. Below we discuss each one in detail.

### Limitation of scope

The current time frame that is analyzed creates a hard boundary between week 10 and week 53 of 2020. The exit conditions of different countries at these boundaries, however, are not equal. Some countries experienced subsequent surges in January 2021 and later months. Thus the current research provides a snapshot of the effects of the pandemic up to the end of 2020, not the totality of its effects.

### Limitation of Data

All cause mortality figures for 2020 are still provisional for most EU countries, so they are subject to readjustment in future time. Even so, they can provide a good estimate of the effect of COVID-19 in different countries up to this point.

### Limitation of Excess mortality and P-scores

Our mean mortality calculations do not account for population changes in the 2015-2019 period. For example, Bulgaria has one of the highest negative growth rates in the world. Between 2015 and 2020 Bulgaria’s population has decreased by around 250,000 or approximately 6.5% per year ^28^. The country also has a negative net migration and has seen approximately 25,000 people leave its borders in the last five years ^29^

Since the P-score metric we compute is derived from the excess mortality figures we calculate for each individual country, this metric also suffers from the issues we outline for excess mortality.

### Limitations of PYLL/ASYR/WYLL

Since PYLL, ASYR and WYLL data only take into account fatalities, these metrics do not provide information about any worsened quality of life of surviving individuals, reduced life expectancy of these individuals and working capacity. Metrics such as Disability-Adjusted Life Years (DALY), Quality-adjusted life year (QALY) and Healthy Years of Life (HALE) metrics may illuminate further the total disease burden on the European population, however, obtaining the necessary information for these measurements is not yet possible.

As mentioned before, due to data availability limitations from Eurostat in our computations of PYLLs and ASYRs we excluded the 90+ group. Given that countries like France, Italy and Spain have significant excess mortality in this age group, we also present a computation of the ASYRs including the 90+ age by assuming 4 years of life expectancy (the average life expectancy for the 90+ age group for the European population is 4.74, according to the UN-data Data Service) in Supplementary Tables 4 and 5. By linear interpolation, 4 is approximately the life expectancy for the age interval [90, 94] assuming that the average age of deaths for this interval is in the range 92.7 − 93 (the average age of the COVID-19 fatalities above 90 years of age is 92.7 in Czechia and 92.1 in Bulgaria, and likely it’s higher in Western countries which overall have higher life expectancy).

This rough approximation gives an upper bound of how large the ASYRs can go. It leads to 5% − 14% and −14% −22% increase in the ASYRs for the (0 − 89) population of Eastern and Western European countries, respectively, but it does not yield a decrease between the inequalities of the countries from the two groups or any significant change in their ranks (see Supplementary Fig. 3 and Supplementary Table 5).

The WYLL measure we present has some additional limitations. The first comes from the assumption that retirement age across European countries is 65. While it is most often assumed as a standard between European countries, there is actually some variation between individual member states ^30^. Furthermore, we assume that the mean age of people who have died in a given age group is the middle of the given range, e.g. for the age group 60-64 - mean age = 62.5. It may well be a fact that a majority of the fatalities are concentrated in the upper part of the age bracket. However, since we do not have data about the different causes of mortality, but rather an aggregate total, we cannot be certain that this trend will hold true for all age groups and across different countries.

### Google COVID-19 Community Mobility Reports

Bulgaria is below the EU average when it comes to use of mobile devices in the 16-74 age group. Still, a majority of the population within that group (∼64%) utilized mobile devices to access the internet in 2019^31^. However, it is possible that there might be a skew towards the younger half of this age range of users who are supplying data.

## Results

### Mortality during the COVID pandemic in Bulgaria

We analyzed overall excess mortality patterns in Bulgaria for the year 2020 and compared it to data for other European Union (EU) countries for the same period. We focus on excess mortality rather than officially registered COVID deaths because limited testing and varying standards for official reporting of COVID deaths can result in large disparities between public figures for COVID-related mortality and the actual burden the disease has imposed on the population ^6^. While some of the excess deaths are caused by the collapse of healthcare services during peak moments of COVID waves, when a particularly large discrepancy between official COVID deaths and excess deaths is observed, this is likely mostly due to underreporting of COVID deaths due to insufficient testing and other irregularities.

In total, we estimate that 17,352 lives have been lost in Bulgaria in 2020 in excess of the baseline from previous years (Figure 1A). This amounts to an EMR of 2,496 DPM, or∼0.25%, for the year and ranks the country as the most highly affected within the EU (Figure 1A; Supplementary Table 1; according to P-scores Spain, Poland and Belgium rank higher). COVID mortality is in most countries higher in males than in females ^32^, and this is also what is observed in Bulgaria and most other EU countries (Figure 1B-C; Supplementary Tables 2-3). For females, an EMR of 2,178 DPM is observed (P-score of 18.79), compared to an EMR of 3,198 DPM for males (P-score of 23.99) across all ages.

**Figure 1:**
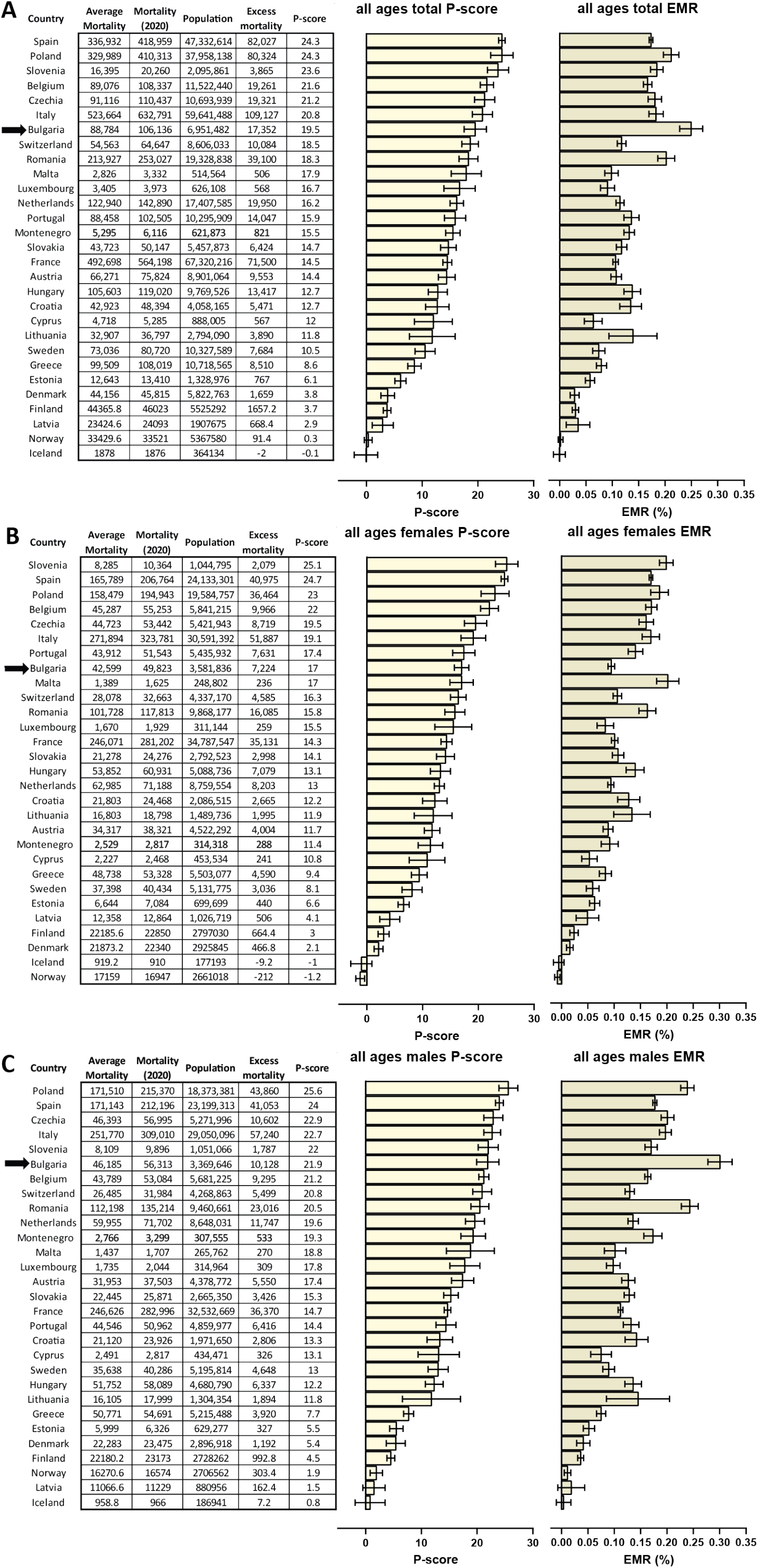
Excess mortality in Bulgaria and other EU countries in 2020. (A) Overall P-scores and excess mortality (in deaths per million; DPM) for all ages in Bulgaria (highlighted in red) and other EU countries; (B) P-scores and excess mortality for females of all ages; C) P-scores and excess mortality for males of all ages.

These observed EMR values are much higher than the officially reported COVID-attributed population fatality rate (PFR), by a factor of ∼2.3 ×. Examination of the EMR/PFR ratios in Europe showed that excess deaths are higher than official COVID death tolls in most countries (Figure 2). However, a clear dichotomy emerges between Eastern and Western Europe, with the EMR/PFR ratio being considerably higher in countries in Eastern Europe such as Bulgaria, Romania, Poland, Slovakia, Lithuania, and others.

**Figure 2:**
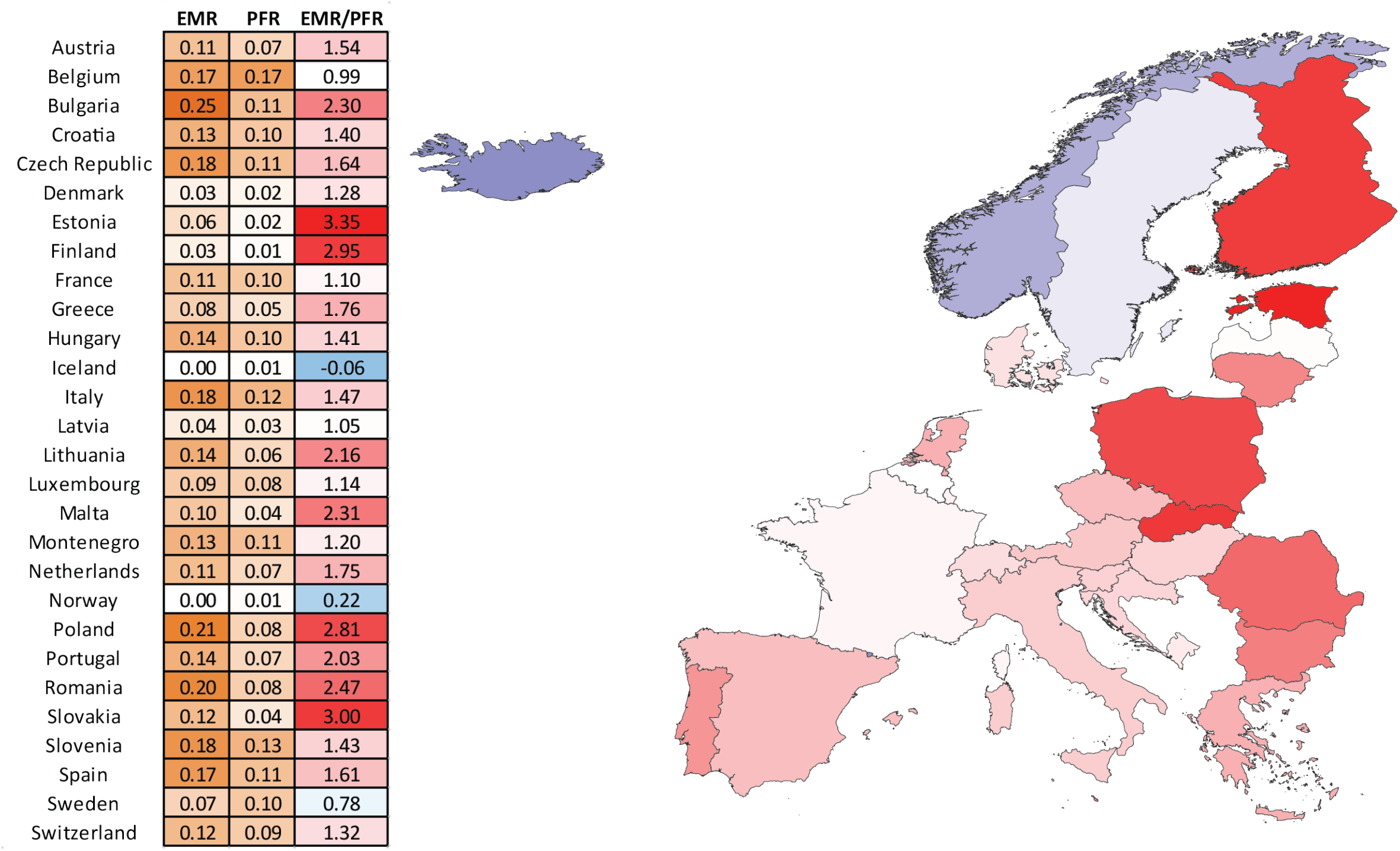
Ratio between excess mortality and official COVID-attributed deaths in European countries in 2020. Note that the high EMR/PFR ratios for 2020 in countries like Finland and Estonia might be an artifact of overall low both excess and COVID-attributed mortality.

These estimates and geographic patterns are in agreement with other recent analyses of excess mortality ^33,34^.

### Loss of life as a result of the COVID pandemic

We next examined the impact of the pandemic in terms of years of life lost using the PYLL and ASYR metrics based on excess mortality (Figure 3). Both metrics paint a similar picture, which is also consistent with the raw excess mortality measures.

**Figure 3:**
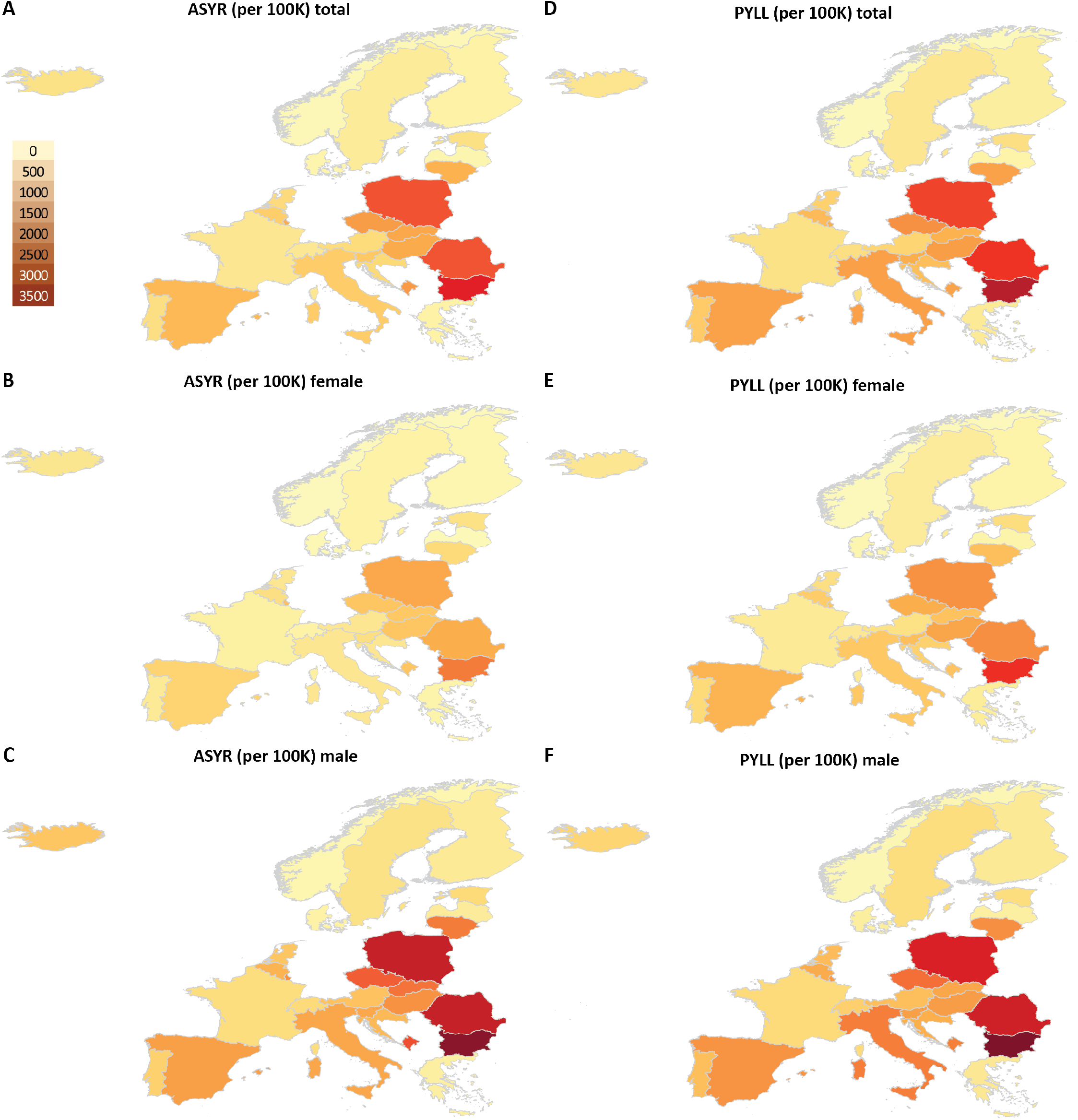
Geographic distribution of excess mortality-based ASYR and PYLL values for European countries in 2020. Shown are the total (per 100K people) values. (A) ASYR values for the whole population; (B) ASYR values for females; (C) ASYR values for males; (D) PYLL values for the whole population; (E) PYLL values for females; (F) PYLL values for males.

Using standardized ASYR and PYLL values (per 100,000 population; Supplementary Figures 2A-C and 1), we find that the highest total loss of life among the examined countries occurred in Bulgaria, for both males and females, followed by Romania, Poland, Hungary and Czechia. This higher loss of life burden in Eastern European countries is explained not only by their high EMRs but also by a large numbers of deaths in younger age groups. In Bulgaria and Romania, 28% and 29% of excess deaths, respectively, are of people under the age of 70. In Poland, 13% of the excess deaths are of people in the age ingerval 65 − 69 and 42% of the excess deaths are of people under 75. Moreover, 18% of all excess deaths in Bulgaria are of people under the age of 65, in particular in the 40-64 age group (Supplementary Figure 4). Calculation of WYLL values, which show the loss of working years of life, showed Bulgaria to have incurred the highest such loss within the set of examined countries (Figure 2D-E; note that the high total WYLL value for Iceland is possibly an artifact of the small population of the country). In contrast, in countries such as Italy, France and Spain, only 18% − 19% of excess deaths are under 75 years of age. In Italy and Spain, 34% − 35% of the excess deaths are of people older than 90. In France, 46% of excess deaths are of people older than 90. Remarkably, in Greece, one of the least affected countries in the EU (having a low EMR and P-score), 55% of all excess deaths are of people above 90 and 20% are people under 80.

For Bulgaria, we find an average PYLL value of 13.46 ± 0.11 in total, 12.56 ± 0.03 for males, and 13.68 ± 0.24 per female (Figure 2). Excluding outliers (note that average PYLL values based on excess mortality are very high in countries such as Iceland, Luxembourg due to stochasticity associated with the very low number of excess deaths), these values are generally higher than what is seen in Western Europe. The only three countries with an average PYLL greater than 13 are Bulgaria, Poland, and Romania, compared to values as low as in the 10 to 11 years range for countries such as Denmark, Switzerland and Sweden (Supplementary Table 17). Despite males exhibiting higher mortality due to COVID-19, the average PYLL based on excess deaths in Bulgaria is higher for females (it is also higher for females in several other European countries; Supplementary Figure 3).

Using official COVID-attributed deaths, for Bulgaria we obtain an average of 12.37 years lost for males and 14.01 years lost for females. Based on the official COVID-19 mortality data for Czechia (the other country for which exact data about the age of the diseased is available) we obtain 9.78 and 9.35 for males and females, respectively. In both cases, the estimates we obtain for the average PYLLs from excess mortality and official COVID-19 deaths data are in agreement (note, however, that there are substantial differences between Bulgaria and Czechia in other aspects – for example, the average age of officially registered COVID-19 deaths for women is 71 years in Bulgaria compared to a life expectancy of 78.4 years, while in Czechia, the average age of the female COVID-19 deaths is 80.81, which is very close to the 82.1 life expectancy for women in that country).

These observations suggest that the impact of the pandemic in hard hit in late 2020 countries in Eastern Europe was not only large in absolute terms but also heavily affected younger demographics than in Western Europe.

### Demographic-specific mortality patterns in Bulgaria

By the official statistics of the Ministry of Health ^18,19^ the average age of a deceased male and female from COVID-19 are 69 and 71, respectively. The leading comorbidity is cardiovascular disease (55%), followed by diabetes (17%), pulmonary disease (12%), obesity (3%), and 30% are listed with no known comorbidity. An overwhelming majority of 94.5% of all 7576 COVID-19 deaths occurred in the hospitals with working age deaths comprising 28% of all COVID-19 deaths. For the working age group females on average died at age 55.9 and males at age 55.7 with 45% of the deceased having a cardiovascular disease.

Data on excess mortality for people under 65 reveals a slightly different picture. The working age group excess deaths are 18.3% of all excess deaths with an average age of the deceased 55.65 ± 0.07 for men and 57.57 ± 0.28 for women. The reason for the higher average age for women is that our data does not reveal excess deaths in women under 40, whereas in the official statistics 5% of the casualties are of ages between 10 and 39.

Next, we examine mortality in Bulgaria within the working age population in detail. Due to the well-documented age-related skew of COVID fatalities, we focused on two subgroups of working age individuals – those in the 30-39 and those in the 40-64 age ranges.

We find no elevated mortality in females in the 30-39 age group, while mortality is elevated in males of the same age bracket, with P-scores of –0.39 and 9.37, respectively (Figure 4A-C).

**Figure 4:**
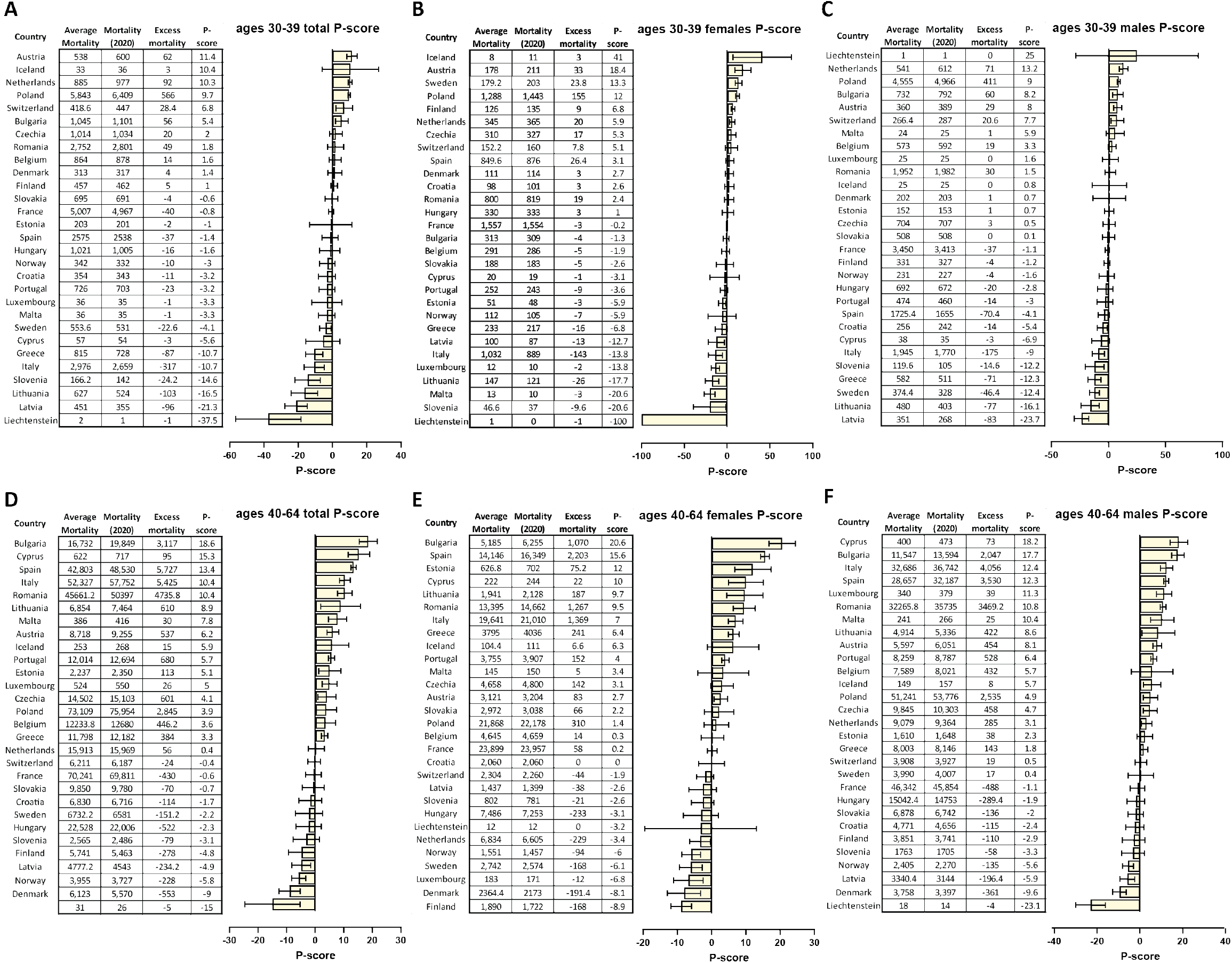
Excess mortality in working age populations in Bulgaria and other EU countries in 2020. (A) P-scores for the overall population in ages 30-39; (B) P-scores for females in ages 30-39; (C) P-scores for males in ages 30-39; (D) P-scores for the overall population in ages 40-64; (E) P-scores for females in ages 40-64; (F) P-scores for males in ages 40-64.

In contrast, we find highly elevated excess mortality in both males and females in the 40-64 age group, in which Bulgaria ranks highest in the EU (Figure 4D-F), with P-scores of 17.7 and 20.6 for males and females, respectively. The difference between males and females is remarkable, as, unlike the typical situation, elsewhere in the world in this group in Bulgaria excess mortality measured by P-scores is lower for males than for females. A similar reversal of the usual sex-specific mortality pattern is only also observed in Spain within the EU. We discuss the possible explanation for these observations in subsequent sections.

### Regional disparities in COVID pandemic-related mortality in Bulgaria

Following from the observation of considerable disparities between different European regions, we then analyzed regional differences in pandemic impacts within Bulgaria (Figure 5). As a reminder, the overall statistics for Bulgaria are an EMR of 0.25%, P-score of 19.5%, CFR of 3.7%, an EMR/PFR ratio of 2.3, and a percentage of population tested positive of 2.9%.

**Figure 5:**
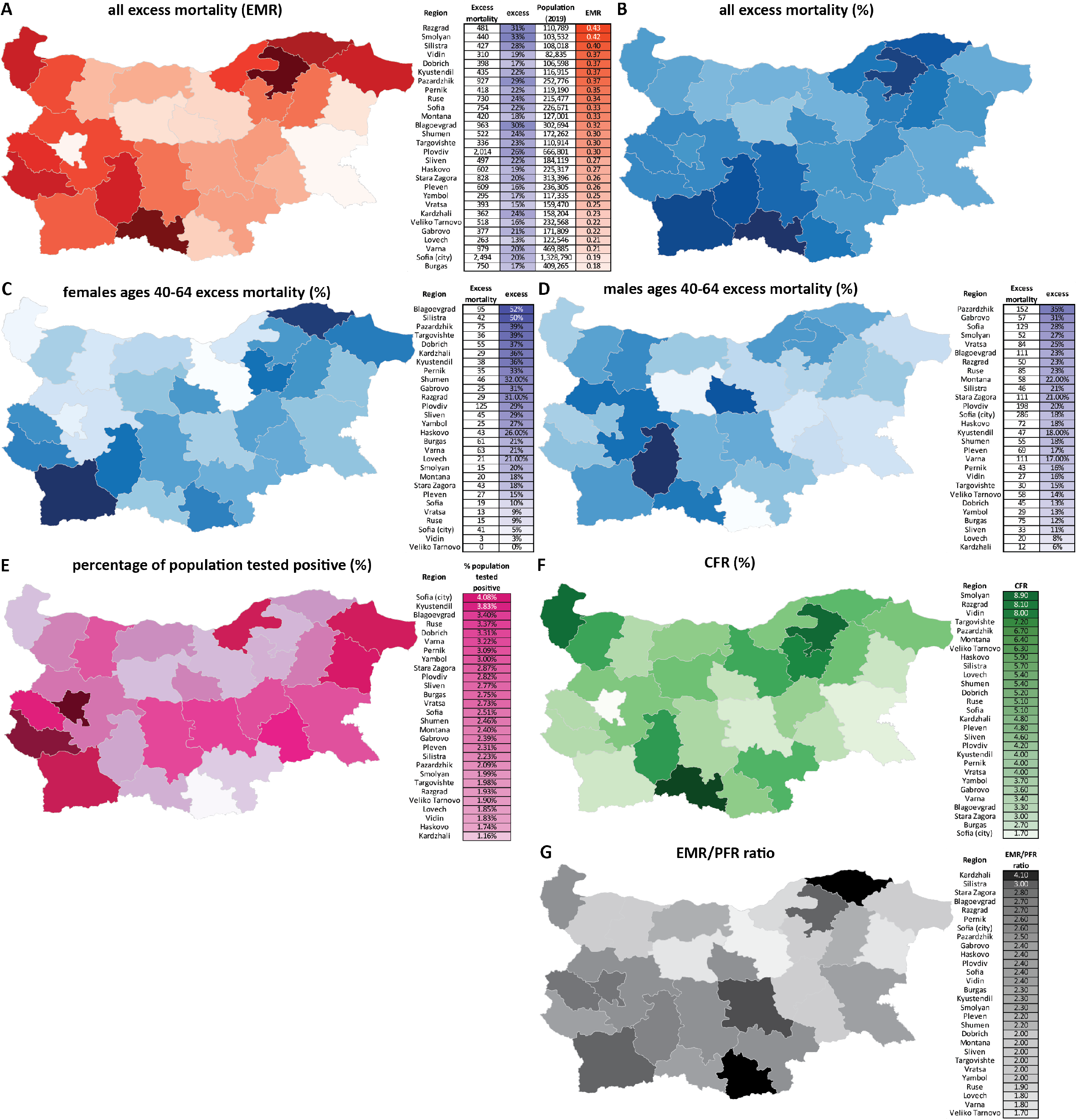
Regional disparities in the impacts of the COVID-19 pandemic in Bulgaria. (A) Overall excess mortality in Bulgarian regions (EMR units); (B) Overall excess mortality in Bulgarian regions (P-score); (C) Excess mortality in working age (40-64) females in Bulgarian regions (P-score); (D) Excess mortality in working age (40-64) males in Bulgarian regions (P-score); (E) Percentage of the population who have tested positive for SARS-CoV-2 in Bulgarian regions; (F) CFR values for Bulgarian regions; (G) Ratio between excess deaths (EMR) and official COVID-19-attributed deaths (PFR).

The first major such disparity we observe is that between the four most populated provinces and the rest of the country. The excess deaths in these four major regions – Sofia (city), Plovdiv, Varna and Burgas – account for just 34% of all excess deaths even though ≥50% of the Bulgarian population lives there. Moverover, Sofia (city), Varna and Burgas have the lowest EMR of all provinces (Figure 5A) and P-scores of up to 20% (Figure 5B). The provinces of Sofia (city) and Burgas also show the two lowest CFR values (Figure 5F).

In contrast, the more peripheral regions are among the most heavily affected. For example, the regions of Vidin and Silistra exhibit some of the highest EMRs – 0.37 and 0.40, respectively. Vidin also has the second highest CFR (8%). The relatively low P-score of 19% in Vidin is likely a result of already very high pre-pandemic mortality in the region (the region has one of the fastest aging populations in the EU and the death rate there is 22 per 1000 people per year, whereas the death rate for Bulgaria is 15 deaths per 1000 people per year). In Silistra, the EMR/PFR ratio is 3.00, the second highest in the country and the P-score is 28%. In Kardzhali, the EMR/PFR ratio is 4.00, the highest in the country, and the P-score is 24%. With a P-score of 31% and EMR of 0.42, Smolyan is the hardest hit region in the country. It also has a CFR of 8.9%, which is the highest among all provinces.

These regions also tend to show a lower percentage of the population that has tested positive, despite exhibiting the highest excess mortality (Figure 5E), often high CFRs (Figure 5F), and high EMR/PFR ratios (Figure 5G).

We also find curious disparities in regional patterns of male- and female-specific excess mortality (Figure 5C-D; Supplementary Tables 18-19). The highest male excess mortality was observed in Pazardzhik, Gabrovo, Sofia (region) and Smolyan, while the highest female excess mortality is seen in Blagoevgrad, Silistra, Pazardzhik and Targovishte.

Remarkably, only 30% of all excess deaths in the working age group occurred in Sofia (city), Plovdiv, Varna and Burgas. For women in the working age group only 27% of the deaths occurred in those regions. A regional analysis reveals that the regions that with the highest P-scores in this demographic category are Blagoevgrad, Silistra, Pazardzhik, Targovishte, Kardzhali, Kyustendil and Dobrich ranging from 52% ± 6% to 36% ± 7% (Figure 5C). Women in the age group 65 − 69, which includes working women in retirement age, were also heavily affected with an overall P-score of 23.7% and exceptionally high regional P-scores in the provinces of Sliven - 76% ± 10%, Kardzhali - 46% ± 8% Blagoevgrad - 45% ± 7%, Pazadzhik - 43% ± 6%, and Smolyan - 42% ± 11% and (Supplementary Figure 5; Supplementary Tables 20-21). We discuss the possible explanations for these observations in the Discussion section.

### The trajectory of the pandemic in Bulgaria and the effectiveness of implemented pandemic control measures

Finally, we mapped the trajectory of the pandemic in Bulgaria onto the timeline of imposition of social distancing measures and independent measures of actual changes in societal mobility to understand the relationship between those factors and its development (Figure 6).

**Figure 6:**
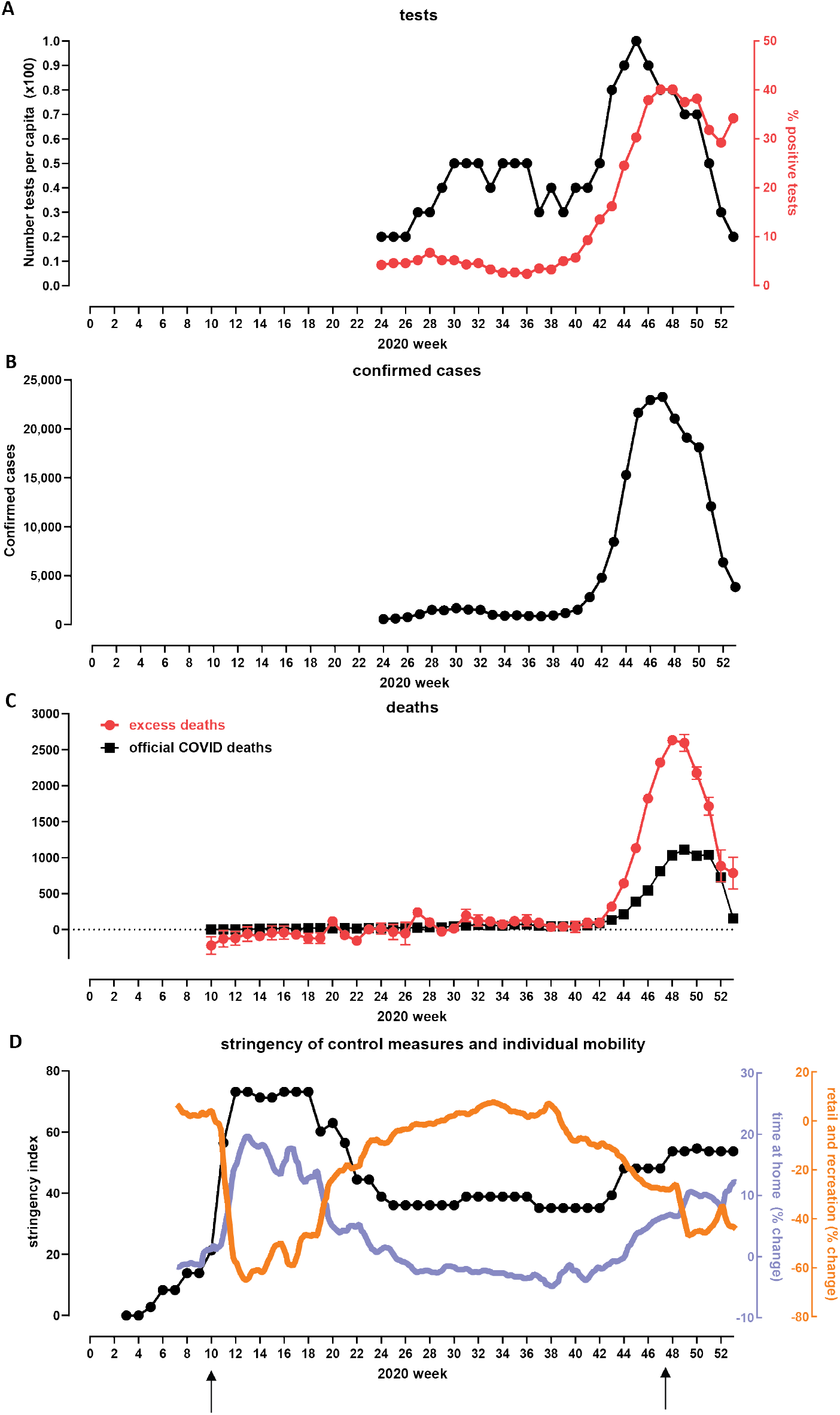
Development of the COVID pandemic in Bulgaria over 2020 and the effectives of measures implemented in order to control it. (A) Number of tests conduced and test positivity percentage over the second half of 2020^*a*^. (B) Officially registered COVID cases. Note that rapid antigen tests were only included in statistics starting from December 22nd 2020. (C) Officially registered weekly COVID deaths and overall weekly excess mortality over the course of 2020. (D) Social mobility changes and the timing of imposition of restrictions. Arrows indicate the time of imposition of “lockdown” measures. “Time Home” refers to the change of the number of visitors to residential areas relative to the period before the pandemic. “Time Retail and Recreation” refers to the change of the number of visitors to places of retail and recreation relative to the period before the pandemic. This includes restaurants, cafes, shopping centers, theme parks, museums, movie theatres, libraries. ^*a*^Official daily testing data was only made available from 06 Jun 2020 – Open Data Portal (https://data.egov.bg/data/resourceView/e59f95dd-afde-43af-83c8-ea2916badd19)

The first period of the COVID-19 pandemic in Bulgaria, from March to the end of September, was marked by a slight elevation of the new confirmed cases in the summer, peaking at 242 cases per day towards the end of July. The excess mortality for this period is around 300 people and the official COVID-19 death toll amounts to 820 people, with a daily death rate of up to 10 deaths until the middle of October.

Rapid growth in the number of new confirmed cases started around the end of September. Then an explosion of cases occurred in late October, November and the first half of December (Figure 6B). The peak in the 7-day moving average of the number of confirmed cases occurred on November 19th.

Official COVID-19 deaths peaked on December 6th with a 7-day moving average of 140, or ∼18 DPM/day; excess deaths started decreasing around the same time (Figure 6C). Excess deaths began diverging from official statistics with the start of the Fall surge, in the middle of October, and peaked at ∼54 DPM/day in the week ending on November 27th. This corresponds to a 112.3% increase in relative age-standardised mortality rates (rASMRs) according to the ONS ^35^; a higher number in Europe in 2020 was observed only in Spain for the week ending on April 3rd at at 142.9%.

One of the obvious candidate explanations for the discrepancy between official and excess deaths is insufficient testing ^36^.

Indeed, that appears to be the case for Bulgaria. Test positivity rates peaked ∼40% in late November. However, a curious pattern is observed in the number of tests recorded in official statistics, which actually began decreasing while the positivity was still increasing in the month of November (Figure 6A). An explanation for this pattern is that the results of rapid antigen tests were not included in official statistics until late December, and a considerable portion of testing shifted from PCR to antigen tests as the Fall wave developed. This likely accounts for at least some of the discrepancy between recorded and excess deaths.

We then examined the factors responsible for the Fall surge eventually receding using the stringency index and mobility metrics (see Methods), changes in which have been shown before to be predictive of the trajectory of COVID epidemics ^37–40^.

The stringency index was at 35.19 from mid September until October 29th (Figures 6D and 7B), when the Bulgarian government imposed some new restrictions (high schools and universities moved to remote learning; nightclubs, pubs and bars were closed), which is reflected by an increase in the stringency index to 48.15. No further substantial epidemiological measures were introduced until after the peak of the fall wave – on November 27th, restaurants, bars, malls, schools and gyms were closed. However, the stringency index, though now increased to 53.7%, remained considerably below the levels of restrictions imposed in other European countries (Figure 7A), as no stay-at-home orders or curfews were imposed, non-essential stores and hair-dressing salons remained open, and gatherings of up to 15 people were permitted.

**Figure 7:**
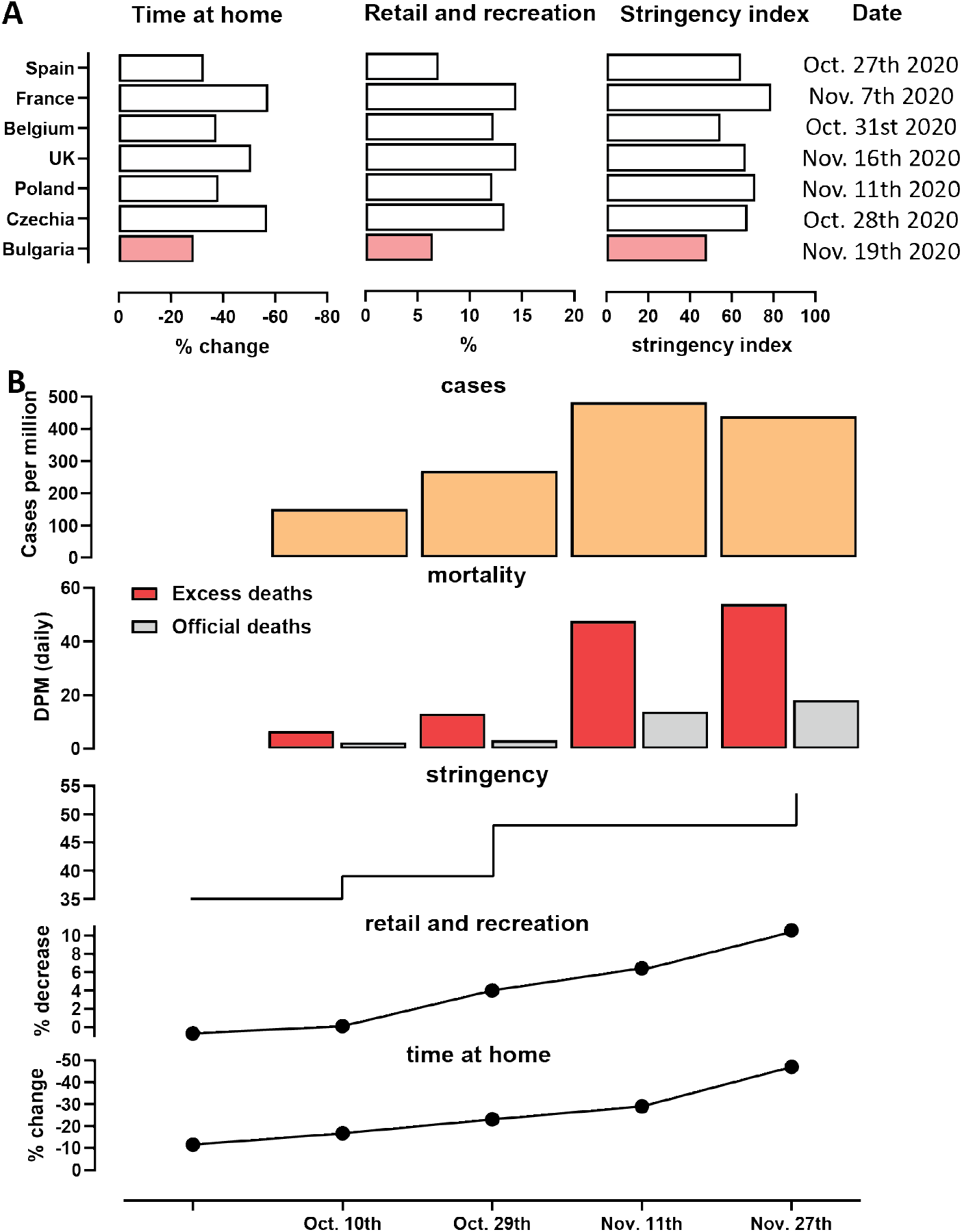
Mobility metrics, stringency of restrictions and mortality and cases at the peak of the late-2020 wave in Bulgaria. (A) Google Mobility Data and Stringency Index at the peak of the fall wave in Bulgaria and other EU countries. “Time Home” refers to the change of the number of visitors to residential areas relative to the period before the pandemic. “Time Retail and Recreation” refers to the change of the number of visitors to places of retail and recreation relative to the period before the pandemic. This includes restaurants, cafes, shopping centers, theme parks, museums, movie theatres, libraries. (B) Timeline of imposition of social distancing measures and of reductions in mobility in Bulgaria around the peak of the late-2020 wave. The peak occurred around November 11th 2020, as demonstrated by mortality data, which at any given moment reflects the dynamic of new cases in Bulgaria approximately 2.5 weeks prior to that moment.

As the peak of restrictions occurred around the time of the peak of excess mortality and thus after the peak of infections, it is likely that restrictions were not the main cause for the eventual decline in cases. Indeed, changes in people’s behavior as reflected in social mobility measures were observed much earlier than the imposition of restrictions, likely due to fear of becoming infected spreading among the population, a pattern previously noted elsewhere in the world ^41^. Our data supports this: 40% of the total increase from October 1st to November 27th of the time spent at home occurred in the period November 3rd to November 27th when no new substantial measures were introduced (see 7B).

The 7-day running average Google mobility data measured on November 19th shows a total decrease of −29% percent of time spent in retail and recreation and a 6.43% increase in time spent at home compared to the baseline (Figures 6D and 7B). However, as with the stringency index, these values are still the lowest among analyzed European countries (Figure 7B), which is likely a contributing factor to the very high excess mortality resulting from the pandemic.

Finally, we examine hospitalization trends in Bulgaria and several other European countries. We find that at their peak on December 12th, hospitalizations in Bulgaria reached a level of 0.1% of the population, which is one of the highest hospitalization rates up to date (it has since been exceed by Hungary and by Bulgaria itself during the subsequent March surge; Figure 8).

**Figure 8:**
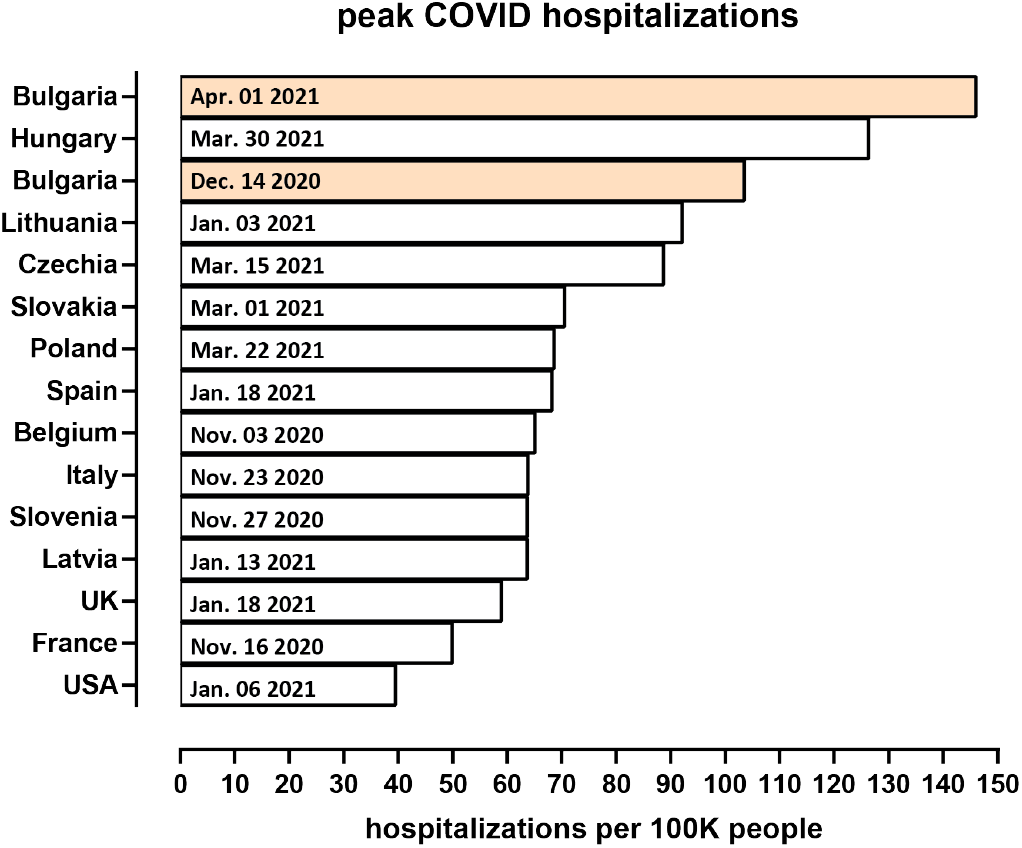
Peak hospitalizations in Bulgaria and other countries.

## Discussion

In this survey, we analyze the impact of the COVID-19 pandemic in 2020 in Bulgaria and the broader Eastern European context. After a relatively low level of COVID-19 cases and deaths prior to that, in the concentrated span of less than three months in October, November, and December 2020, Bulgaria recorded the largest (per capita) number of excess deaths among the examined countries. Similar, though somewhat lower, large excess mortality increases were observed in most other countries in Eastern Europe. However, official COVID-19-attributed deaths account for only less than half of the excess deaths.

This discrepancy is likely caused by a combination of multiple factors – COVID-19 cases leading to death that were not reported as such in official statistics, COVID-19 cases resulting in death some time after recovery due to longer-term complications from the disease, and deaths from other causes that increased as a result to the inability of the healthcare system to treat them due to it being overwhelmed by COVID-19 patients. As the disparity between excess deaths and official COVID-19 mortality is very large in the case of Bulgaria – excess deaths amount to ∼0.25% of the population while official COVID-19 are at ∼0.11%, a ∼0.14% difference – and excess mortality is highly temporally concentrated in a short time span of about ten weeks (i.e. the contribution of the latter two factors is unlikely to have been so large in such a brief period), it is most likely that the bulk of excess deaths were caused directly by COVID-19.

Why they were not recorded as such is also probably due to a multitude of factors. Testing in Bulgaria has been greatly insufficient throughout the pandemic and even more so during the late-2020 surge and the lowest among the examined countries (Supplementary Figure 7; in addition to that, the decision to not include rapid antigen tests in public statistics certainly has contributed to the underreporting. Most reported deaths occurred in hospitals, and many of those who could not be hospitalized due to healthcare systems being overwhelmed and died at home were not recorded as having died of COVID. Whether additional social and socioeconomic factors could have contributed, as has been suggested to be the case elsewhere in the world ^42^, is a subject for future investigations, as is the question of whether the reasons for underreporting are uniform across the more general Eastern European region. Lack of testing on its own in turn has probably contributed to the epidemic growing out of control and leading to such a number of excess deaths.

Another contributing factor to the high mortality rate in Eastern Europe is probably the very high prevalence of cardiovascular diseases in the region ^43^. In Bulgaria over half of the COVID-19 officially reported fatalities are listed with cardiovascular disease as a comorbidity.

Bulgaria also exhibits the most highly elevated working-age excess mortality, and it is also an outlier in terms of working-age excess mortality among females. We also observe significant regional disparities within the different regions in the country in total and in working age sex-specific excess mortality. A possible explanation for the latter is the development of outbreaks at workplaces where mostly women work – for example, garment, textile and shoe factories, which in Bulgaria almost exclusively employee women and which are major sectors of the economy in provinces such as Blagoevgrad, Kardzhali, Smolyan, Sliven, and Kyustendil ^44^. Indeed, there were numerous reports about outbreaks in such settings. Analogous causality might be behind regional disparities in working age male-specific excess mortality (Figure 5D). A list of reports about outbreaks in these regions can be found in our GitHub repository, which includes reports about outbreaks in battery, automotive parts, power transmission, sanitary ceramics, and other factories.

Of note, Spain is another European country with a notable substantial number of working-age female excess deaths. The likely cause in that case is the overrepresentation of middle-aged women among home care workers, who were identified as essential workers early on in the pandemic and were thus affected by it to a greater extent ^45^.

Regional disparities in overall excess mortality, in particular the clear dichotomy emerging between the major population centers, in which generally better outcomes are observed, and the more heavily affected peripheral regions, also warrant further investigation. COVID-19 is still often considered a disease that impacts highly populated big cities the most, where disease spread is thought to be facilitated by density; this is due to many of the most notable initial outbreaks affecting well-connected in terms of international travel large metropolitan areas. However, as the pandemic has spread throughout the countries that have not controlled it, it may be the case that previously established regional disparities in healthcare infrastructure are becoming a key factor determining differential outcomes between generally better resourced major cities on one hand, and the less equipped to test, track and treat COVID-19 patients countryside areas. There is evidence that such causation is at play in Bulgaria – many of the heavily affected regions have fewer ICU beds, fewer doctors, and fewer specialists in the most relevant to the treatment of COVID-19 specialties than the capital and a few other major cities (Supplementary Figure 6). For example, Vidin and Silistra have fewer than average hospital beds, Kardzhali has the lowest number of doctors, general practitioners and pulmonologists and the second to last number of ICU beds per capita in the country, and Smolyan has the lowest number of ICU beds (just 9 in total for the whole region) and a generally low number of doctors.

In addition, in some of these regions (e.g. Smolyan, the most heavily impacted in the country) there are purely geographic factors that may have complicated the timely treatment of patients due to the logistic challenges of transporting patients to the regional center (which is where the only ICU units are located) from remote small towns through mountainous terrain while the core city’s health infrastructure is itself under immense stress (as shown in Figure 8, Bulgaria recorded record hospitalization levels during the peaks of the pandemic). Whether similar regional patterns of pandemic-related excess mortality are observed in other areas of Europe will be informative and instructive for minimizing the impact of subsequent COVID-19 waves.

It should also be noted that healthcare disparities possibly play a role on a broader-level ^46–48^, as Eastern Europe’s healthcare systems as a whole are well-documented to be suffering from an outflow of skilled medical labor due to large numbers of doctors and nurses emigrating to Western Europe in recent years ^49^.

However, the main factor behind the very high levels of excess mortality is still most likely the late imposition of restrictions on social mobility and lax governmental efforts at controlling the spread of SARS-CoV-2, as our analysis shows. In Bulgaria these were adopted long after exponential growth in cases had commenced and was clearly going to overwhelm hospital resources, little testing was carried out and insufficient efforts were made to ensure the isolation of infected individuals, and even when restrictions were imposed, they were generally the most lax in Europe; furthermore, the late-2020 epidemic appears to have begun to trend downward due to changes in individual behavior, the onset of which actually preceded the imposition of restrictions by the government. The high levels of excess mortality are probably a natural consequence of following these policies.

## Data Availability

All datasets and associated code can be found at https://github.com/Mlad-en/COV-BG.git

## Notes

### Competing Interests

The authors declare no competing interests.

## Supplementary Materials

### Supplementary Figures

**Supplementary Figure 1:**
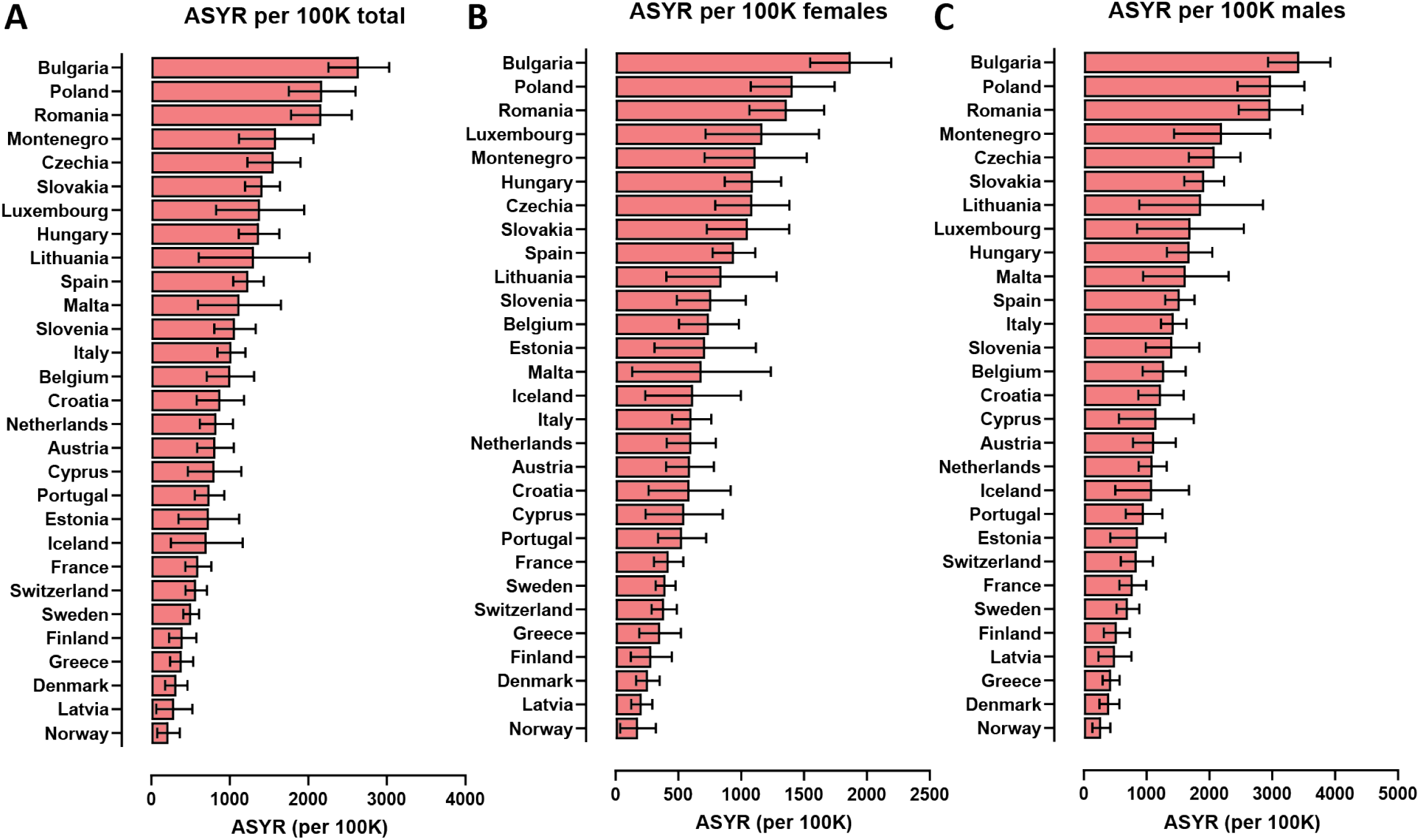
Excess mortality-based ASYR values for European countries in 2020. Shown are the total (per 100K people) values. (A) ASYR values for the whole population; (B) ASYR values for females; (C) ASYR values for males.

**Supplementary Figure 2:**
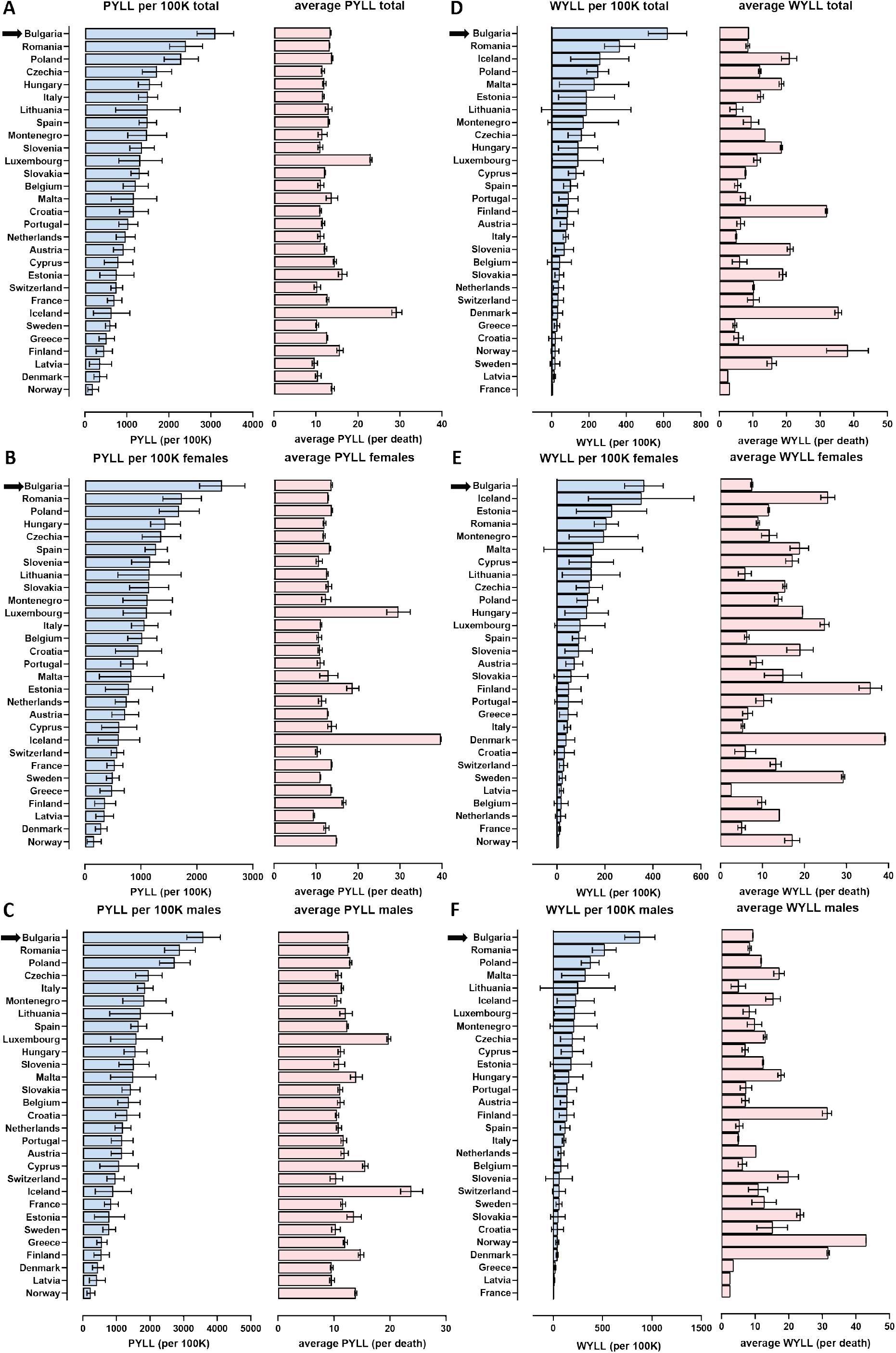
Excess mortality-based PYLLs and WYLL values for EU countries in 2020. Shown are the total (per 100K people) and average (per death) values. (A) PYLL values for the whole population; (B) PYLL values for females; (C) PYLL values for males; (D) WYLL values for the whole population; (E) WYLL values for females; (F) WYLL values for males.

**Supplementary Figure 3:**
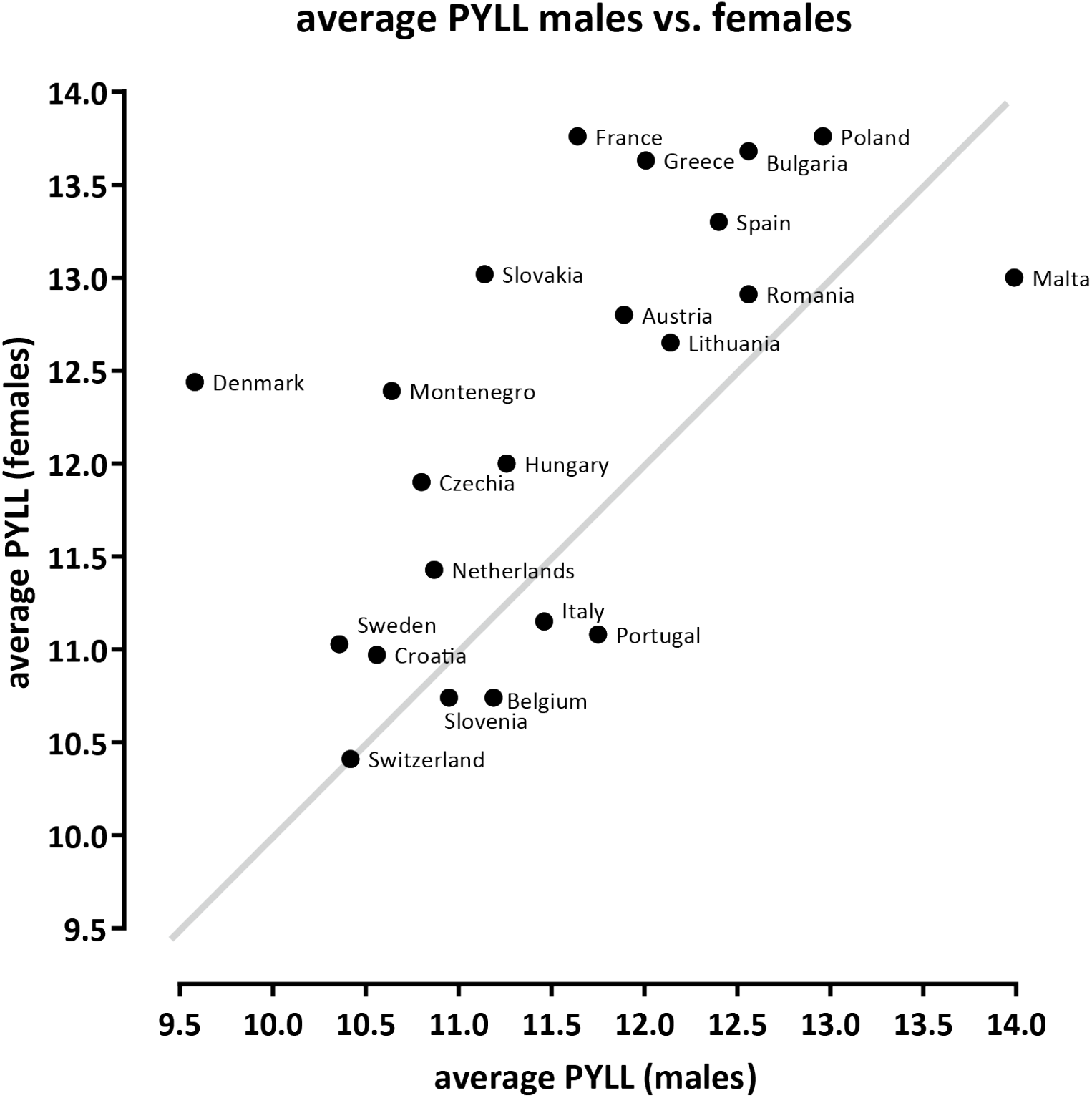
Comparison between male- and female-specific average PYLL values in European countries. Shown are the average PYLL values per excess death.

**Supplementary Figure 4:**
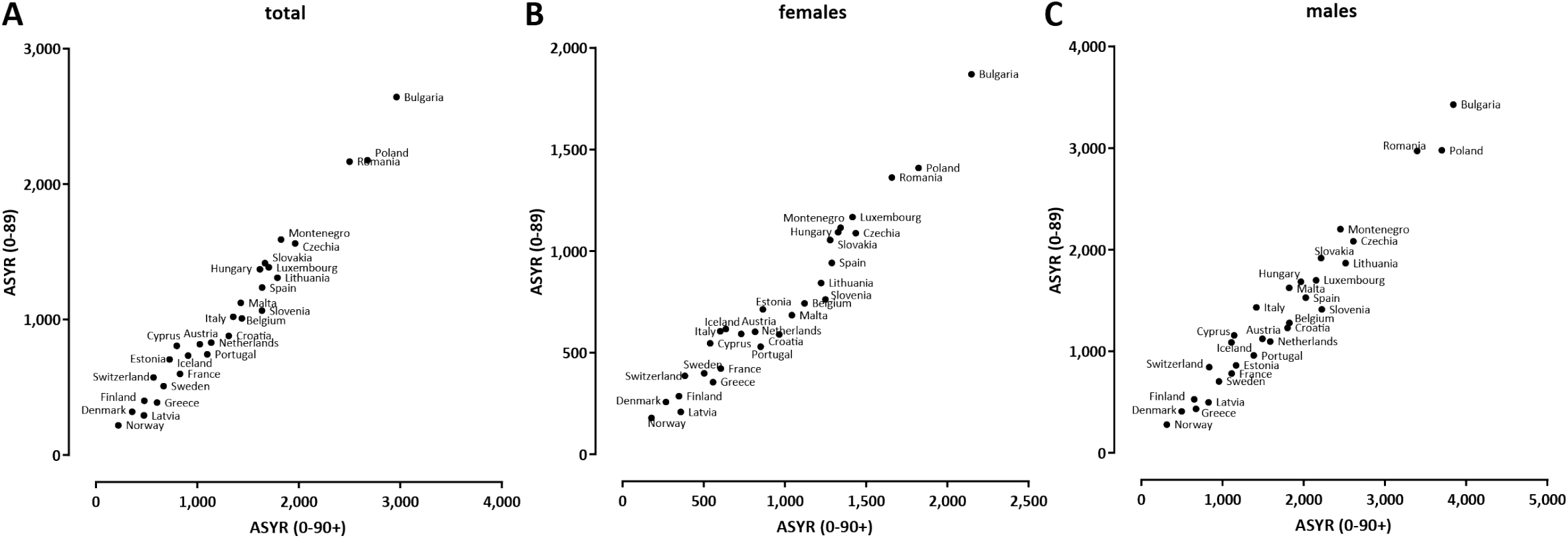
ASYR values computed over the 0-89 age range and ASYR values computed over the whole population (with 90+ year-olds included) are consistent. (A) ASYR total; (B) ASYR females; (C) ASYR males.

**Supplementary Figure 5:**
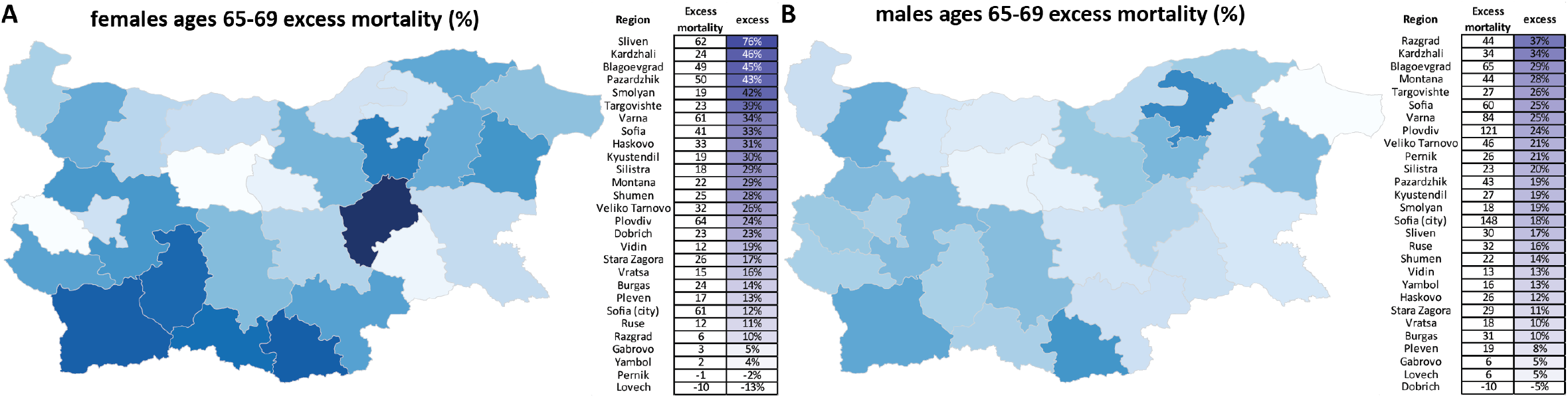
Excess mortality in the 65-69 age range in Bulgarian regions. (A) females, P-scores; (B) males, P-scores.

**Supplementary Figure 6:**
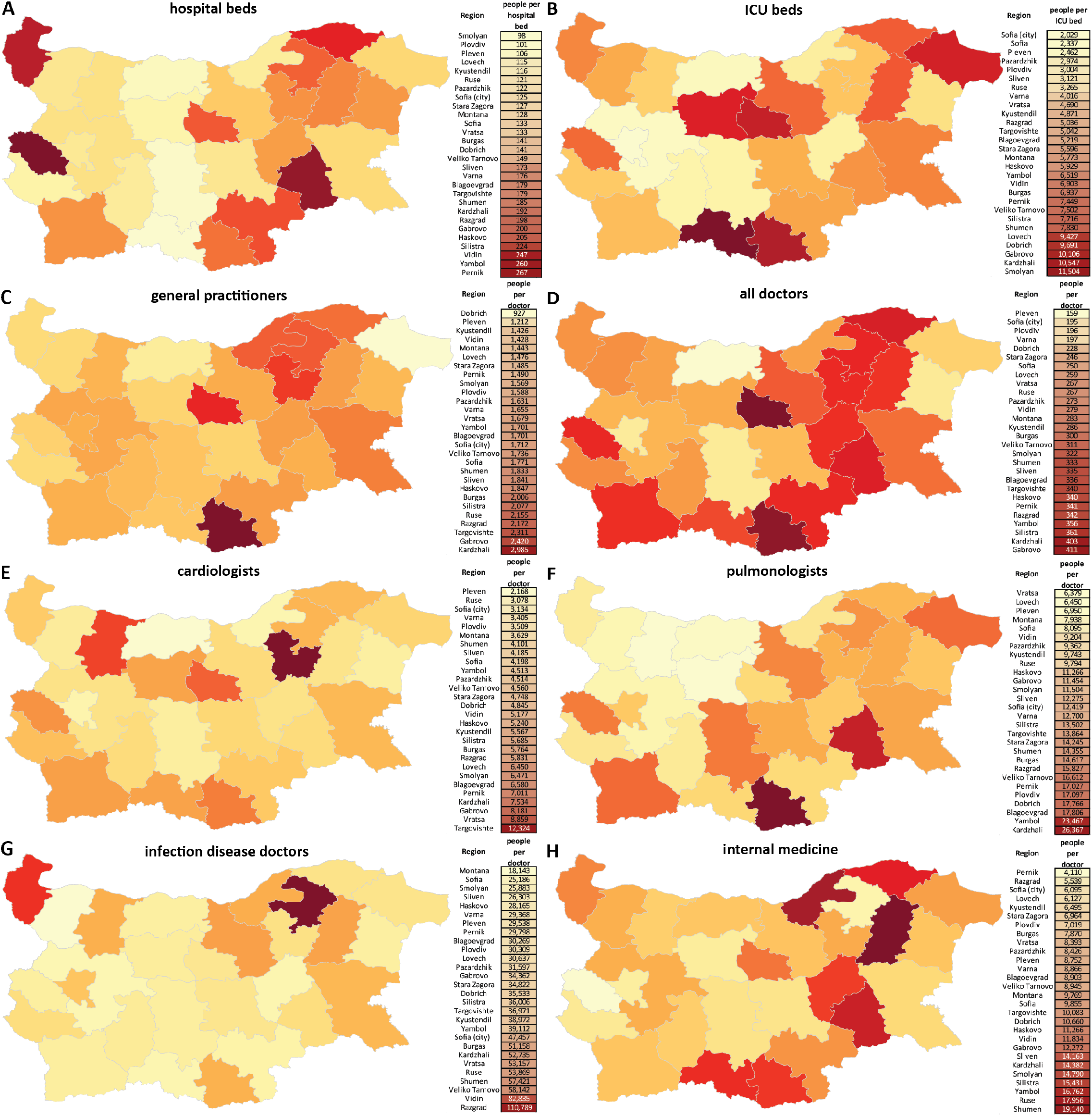
Availability of medical resources in Bulgarian regions. (A) Overall hospital beds (in units of people per bed); (B) ICU beds; (C) Total doctors; (D) General practitioners; (E) Cadiologists; (F) Pulmonologists; (G) Infection disease specialists; (H) Internal medicine specialists. Data from the Bulgarian National Statistical Institute (https://www.nsi.bg/bg)

**Supplementary Figure 7:**
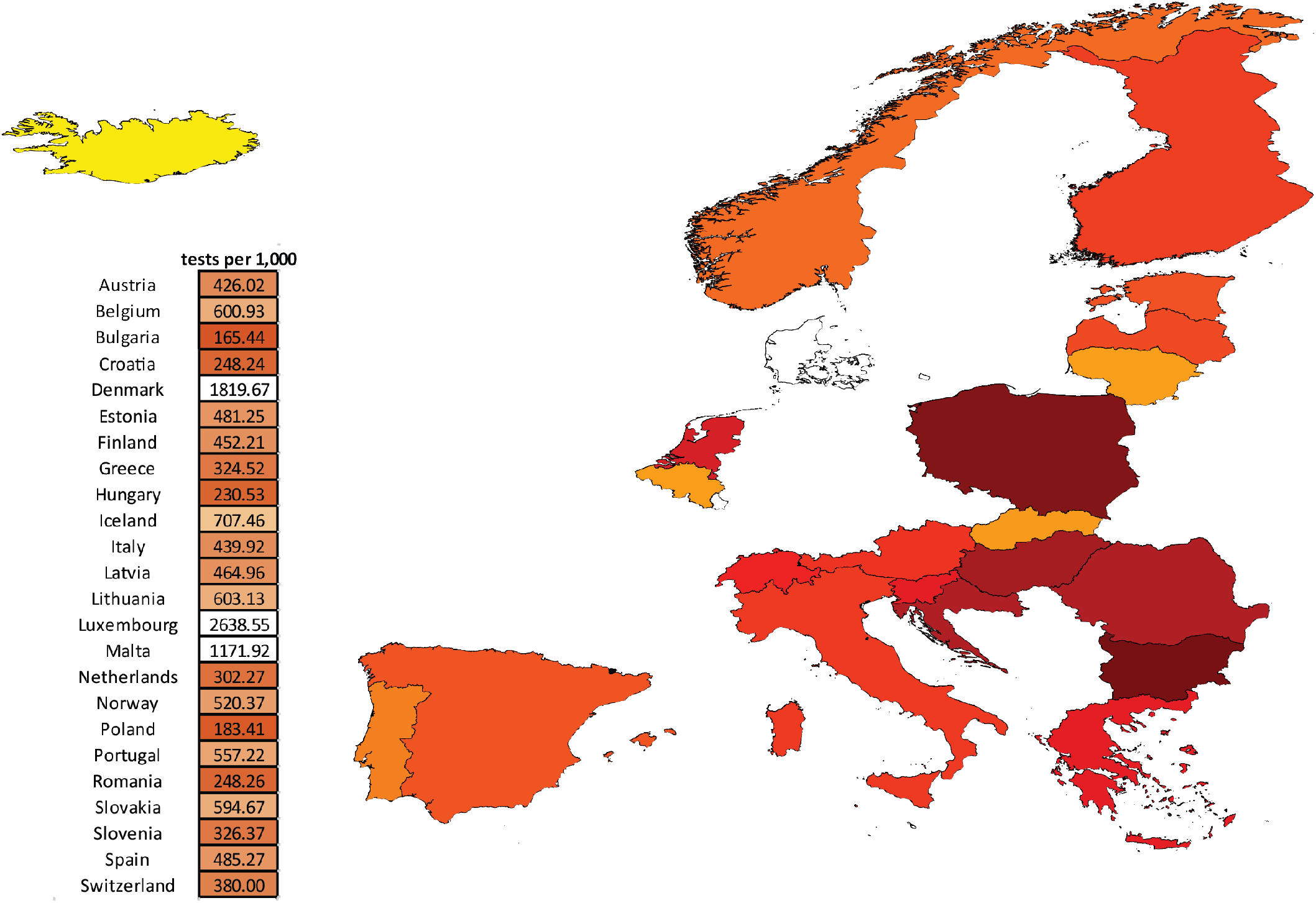
COVID testing in European countries. Shown is the number of tests per 1,000 population for the period up to December 31st 2020. Data from https://ourworldindata.org/coronavirus

### Supplementary Tables

**Supplementary Table 1:**
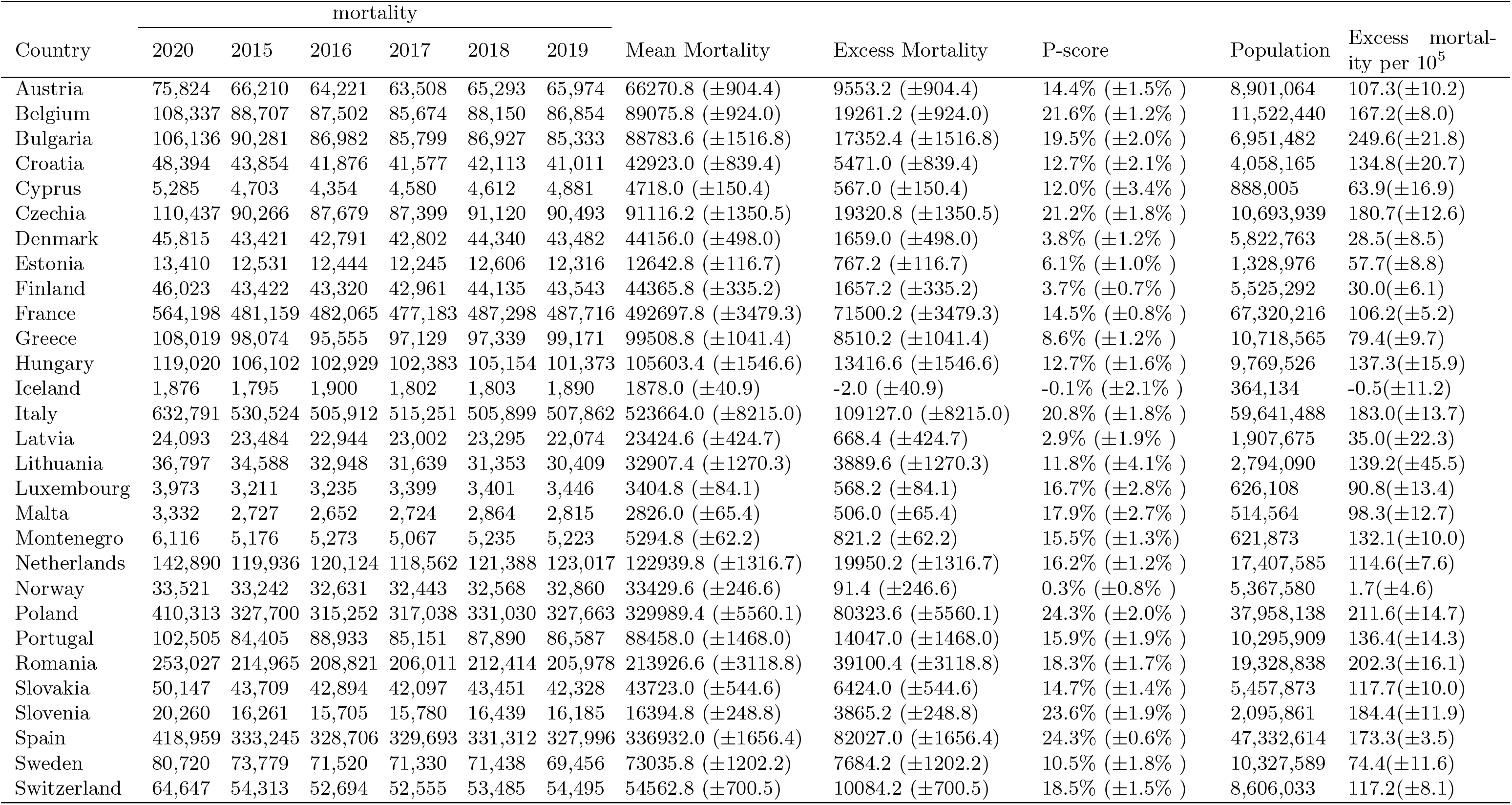
Total excess mortality in European countries in the year 2020.

**Supplementary Table 2:** Excess mortality in European countries in the year 2020 for females (all ages).

| Country | mortality |  |  |  |  |  |  |  |  |  | Excess mortality per 10<sup>5</sup> |
| --- | --- | --- | --- | --- | --- | --- | --- | --- | --- | --- | --- |
|  | 2020 | 2015 | 2016 | 2017 | 2018 | 2019 | Mean Mortality | Excess Mortality | P-score | Population |  |
| Austria | 1,689 | 1,703 | 1,701 | 1,593 | 1,577 | 1,623 | 1663.4 (±46.6) | 25.6 (±46.6) | 1.5% (±2.7%) | 237,161 | 10.8(±19.6) |
| Belgium | 2,625 | 2,379 | 2,267 | 2,261 | 2,325 | 2,332 | 2348.0 (±38.6) | 277.0 (±38.6) | 11.8% (±1.8%) | 321,281 | 86.2(±12.0) |
| Bulgaria | 3,975 | 3,292 | 3,154 | 3,218 | 3,186 | 3,101 | 3240.6 (±56.0) | 734.4 (±56.0) | 22.7% (±2.1%) | 258,560 | 284.0(±21.7) |
| Croatia | 1,466 | 1,195 | 1,285 | 1,314 | 1,272 | 1,176 | 1269.2 (±46.9) | 196.8 (±46.9) | 15.5% (±4.1%) | 142,394 | 138.2(±32.9) |
| Cyprus | 120 | 133 | 114 | 117 | 119 | 119 | 123.6 (±5.8) | -3.6 (±5.8) | -2.9% (±4.4%) | 23,116 | -15.6(±25.1) |
| Czechia | 3,539 | 3,556 | 3,399 | 3,319 | 3,318 | 3,109 | 3401.8 (±126.7) | 137.2 (±126.7) | 4.0% (±3.7%) | 363,248 | 37.8(±34.9) |
| Denmark | 1,385 | 1,566 | 1,478 | 1,319 | 1,421 | 1,400 | 1462.4 (±72.1) | -77.4 (±72.1) | -5.3% (±4.4%) | 163,541 | -47.3(±44.1) |
| Estonia | 403 | 358 | 372 | 370 | 443 | 335 | 379.6 (±31.7) | 23.4 (±31.7) | 6.2% (±8.2%) | 45,393 | 51.5(±69.9) |
| Finland | 1,224 | 1,411 | 1,240 | 1,295 | 1,237 | 1,228 | 1307.0 (±60.1) | -83.0 (±60.1) | -6.4% (±4.1%) | 185,685 | -44.7(±32.4) |
| France | 12,617 | 11,547 | 11,628 | 11,572 | 11,689 | 11,608 | 11827.2 (±42.9) | 789.8 (±42.9) | 6.7% (±0.4%) | 2,067,425 | 38.2(±2.1) |
| Greece | 2,204 | 2,045 | 1,920 | 1,913 | 1,910 | 1,836 | 1958.4 (±59.1) | 245.6 (±59.1) | 12.5% (±3.3%) | 323,057 | 76.0(±18.3) |
| Hungary | 5,359 | 4,130 | 4,062 | 4,286 | 4,414 | 4,421 | 4344.2 (±127.9) | 1014.8 (±127.9) | 23.4% (±3.6%) | 366,614 | 276.8(±34.9) |
| Iceland | 56 | 52 | 51 | 45 | 41 | 44 | 46.6 (±3.7) | 9.4 (±3.7) | 20.2% (±8.9%) | 8,431 | 111.5(±43.9) |
| Italy | 11,505 | 11,506 | 11,179 | 10,875 | 10,296 | 9,861 | 10983.4 (±521.1) | 521.6 (±521.1) | 4.7% (±4.7%) | 1,818,274 | 28.7(±28.6) |
| Latvia | 720 | 722 | 726 | 771 | 810 | 679 | 755.2 (±39.4) | -35.2 (±39.4) | -4.7% (±4.7%) | 65,922 | -53.4(±59.8) |
| Lithuania | 1,055 | 1,001 | 970 | 942 | 932 | 934 | 974.2 (±23.1) | 80.8 (±23.1) | 8.3% (±2.5%) | 92,402 | 87.4(±25.0) |
| Luxembourg | 108 | 101 | 84 | 104 | 89 | 85 | 93.4 (±7.3) | 14.6 (±7.3) | 15.6% (±8.4%) | 13,913 | 104.9(±52.5) |
| Malta | 87 | 111 | 104 | 82 | 83 | 80 | 95.2 (±11.3) | -8.2 (±11.3) | -8.6% (±9.7%) | 14,406 | -56.9(±78.4) |
| Montenegro | 2,817 | 2,542 | 2,482 | 2,390 | 2,539 | 2,445 | 2529.2 (±50.6) | 287.8 (±50.6) | 11.4% (±2.2%) | 314,318 | 91.6(±16.1) |
| Netherlands | 3,967 | 4,120 | 3,871 | 3,626 | 3,580 | 3,595 | 3833.6 (±183.6) | 133.4 (±183.6) | 3.5% (±4.7%) | 502,794 | 26.5(±36.5) |
| Norway | 897 | 951 | 969 | 932 | 910 | 855 | 941.0 (±34.5) | -44.0 (±34.5) | -4.7% (±3.3%) | 138,228 | -31.8(±24.9) |
| Poland | 16,084 | 12,019 | 12,246 | 12,834 | 13,235 | 13,509 | 12997.4 (±496.6) | 3086.6 (±496.6) | 23.7% (±4.5%) | 1,352,561 | 228.2(±36.7) |
| Portugal | 2,082 | 1,829 | 1,894 | 1,848 | 1,782 | 1,792 | 1863.4 (±35.4) | 218.6 (±35.4) | 11.7% (±2.1%) | 337,447 | 64.8(±10.5) |
| Romania | 9,792 | 7,001 | 7,444 | 7,489 | 7,890 | 7,618 | 7634.0 (±253.3) | 2158.0 (±253.3) | 28.3% (±4.2%) | 670,155 | 322.0(±37.8) |
| Slovakia | 2,003 | 1,546 | 1,680 | 1,625 | 1,751 | 1,681 | 1687.8 (±59.8) | 315.2 (±59.8) | 18.7% (±4.1%) | 181,339 | 173.8(±33.0) |
| Slovenia | 487 | 441 | 403 | 419 | 497 | 471 | 458.2 (±29.9) | 28.8 (±29.9) | 6.3% (±6.5%) | 68,823 | 41.8(±43.5) |
| Spain | 7,597 | 5,988 | 6,069 | 6,115 | 6,051 | 6,112 | 6192.6 (±40.8) | 1404.4 (±40.8) | 22.7% (±0.8%) | 1,271,580 | 110.4(±3.3) |
| Sweden | 1,758 | 2,152 | 1,928 | 1,894 | 1,859 | 1,678 | 1943.8 (±133.1) | -185.8 (±133.1) | -9.6% (±5.8%) | 272,942 | -68.1(±48.8) |
| Switzerland | 1,172 | 1,301 | 1,244 | 1,206 | 1,194 | 1,149 | 1246.8 (±44.8) | -74.8 (±44.8) | -6.0% (±3.3%) | 222,768 | -33.6(±20.1) |

**Supplementary Table 3:** Excess mortality in European countries in the year 2020 for males (all ages).

| Country | mortality |  |  |  |  |  |  |  |  |  | Excess mortality per 10<sup>5</sup> |
| --- | --- | --- | --- | --- | --- | --- | --- | --- | --- | --- | --- |
|  | 2020 | 2015 | 2016 | 2017 | 2018 | 2019 | Mean Mortality | Excess Mortality | P-score | Population |  |
| Austria | 2,889 | 2,854 | 2,871 | 2,629 | 2,656 | 2,606 | 2784.0 (±100.8) | 105.0 (±100.8) | 3.8% (±3.7%) | 213,220 | 49.2(±47.3) |
| Belgium | 4,569 | 4,040 | 4,027 | 3,922 | 3,882 | 3,729 | 3996.8 (±99.0) | 572.2 (±99.0) | 14.3% (±2.7%) | 302,248 | 189.3(±32.8) |
| Bulgaria | 7,029 | 6,059 | 5,979 | 5,849 | 5,835 | 5,645 | 5983.0 (±123.9) | 1046.0 (±123.9) | 17.5% (±2.4%) | 205,259 | 509.6(±60.4) |
| Croatia | 2,962 | 2,373 | 2,420 | 2,450 | 2,550 | 2,467 | 2501.6 (±51.3) | 460.4 (±51.3) | 18.4% (±2.4%) | 124,926 | 368.5(±41.1) |
| Cyprus | 224 | 224 | 186 | 221 | 228 | 221 | 220.0 (±13.3) | 4.0 (±13.3) | 1.8% (±5.8%) | 21,813 | 18.3(±61.0) |
| Czechia | 6,826 | 6,384 | 6,260 | 6,005 | 6,146 | 5,953 | 6269.6 (±139.5) | 556.4 (±139.5) | 8.9% (±2.4%) | 315,679 | 176.3(±44.1) |
| Denmark | 2,145 | 2,314 | 2,195 | 2,089 | 2,105 | 2,114 | 2205.0 (±73.4) | -60.0 (±73.4) | -2.7% (±3.2%) | 155,734 | -38.5(±47.1) |
| Estonia | 807 | 695 | 727 | 720 | 723 | 752 | 737.8 (±16.0) | 69.2 (±16.0) | 9.4% (±2.3%) | 32,108 | 215.5(±49.9) |
| Finland | 2,151 | 2,596 | 2,514 | 2,400 | 2,308 | 2,199 | 2438.6 (±124.1) | -287.6 (±124.1) | -11.8% (±4.3%) | 171,275 | -167.9(±72.4) |
| France | 24,034 | 23,344 | 22,913 | 22,656 | 22,558 | 21,980 | 23116.6 (±392.0) | 917.4 (±392.0) | 4.0% (±1.8%) | 1,835,511 | 50.0(±21.3) |
| Greece | 4,363 | 4,223 | 3,857 | 3,886 | 3,800 | 3,874 | 4015.2 (±131.8) | 347.8 (±131.8) | 8.7% (±3.5%) | 285,945 | 121.6(±46.1) |
| Hungary | 8,792 | 6,525 | 7,019 | 6,942 | 7,332 | 7,278 | 7139.2 (±252.5) | 1652.8 (±252.5) | 23.2% (±4.3%) | 277,065 | 596.5(±91.2) |
| Iceland | 86 | 78 | 73 | 80 | 69 | 87 | 79.8 (±5.4) | 6.2 (±5.4) | 7.8% (±6.9%) | 8,550 | 72.5(±63.2) |
| Italy | 20,940 | 19,839 | 19,094 | 18,098 | 17,400 | 16,617 | 18608.0 (±1010.2) | 2332.0 (±1010.2) | 12.5% (±5.8%) | 1,652,740 | 141.1(±61.1) |
| Latvia | 1,368 | 1,229 | 1,290 | 1,305 | 1,321 | 1,246 | 1305.4 (±30.8) | 62.6 (±30.8) | 4.8% (±2.4%) | 43,882 | 142.7(±70.1) |
| Lithuania | 2,078 | 1,878 | 1,734 | 1,690 | 1,737 | 1,752 | 1795.0 (±55.6) | 283.0 (±55.6) | 15.8% (±3.5%) | 62,138 | 455.4(±89.5) |
| Luxembourg | 168 | 136 | 161 | 210 | 160 | 150 | 165.8 (±21.9) | 2.2 (±21.9) | 1.3% (±11.8%) | 13,597 | 16.2(±161.0) |
| Malta | 147 | 165 | 141 | 153 | 158 | 153 | 159.6 (±6.8) | -12.6 (±6.8) | -7.9% (±3.8%) | 14,037 | -89.8(±48.5) |
| Montenegro | 3,299 | 2,634 | 2,791 | 2,677 | 2,696 | 2,778 | 2765.6 (±52.8) | 533.4 (±52.8) | 19.3% (±2.2%) | 307,555 | 173.4(±17.2) |
| Netherlands | 5,745 | 6,058 | 5,581 | 5,310 | 5,309 | 5,144 | 5589.2 (±281.4) | 155.8 (±281.4) | 2.8% (±4.9%) | 493,254 | 31.6(±57.0) |
| Norway | 1,374 | 1,503 | 1,460 | 1,406 | 1,336 | 1,295 | 1432.8 (±67.1) | -58.8 (±67.1) | -4.1% (±4.3%) | 137,044 | -42.9(±49.0) |
| Poland | 31,063 | 22,051 | 22,655 | 23,212 | 24,714 | 25,008 | 23980.0 (±1010.1) | 7083.0 (±1010.1) | 29.5% (±5.2%) | 1,095,263 | 646.7(±92.2) |
| Portugal | 4,148 | 3,524 | 3,730 | 3,629 | 3,660 | 3,724 | 3731.0 (±65.8) | 417.0 (±65.8) | 11.2% (±2.0%) | 285,465 | 146.1(±23.0) |
| Romania | 18,305 | 12,569 | 12,817 | 13,450 | 14,465 | 14,171 | 13771.2 (±646.2) | 4533.8 (±646.2) | 32.9% (±5.9%) | 531,739 | 852.6(±121.6) |
| Slovakia | 3,753 | 2,835 | 3,003 | 2,973 | 3,237 | 3,199 | 3102.2 (±131.0) | 650.8 (±131.0) | 21.0% (±4.9%) | 148,066 | 439.5(±88.5) |
| Slovenia | 1,106 | 798 | 862 | 847 | 960 | 986 | 903.4 (±62.2) | 202.6 (±62.2) | 22.4% (±7.9%) | 65,374 | 309.9(±95.2) |
| Spain | 15,883 | 13,492 | 13,126 | 13,309 | 13,057 | 12,782 | 13394.8 (±209.9) | 2488.2 (±209.9) | 18.6% (±1.9%) | 1,160,179 | 214.5(±18.1) |
| Sweden | 2,693 | 3,090 | 2,830 | 2,584 | 2,648 | 2,421 | 2780.2 (±200.6) | -87.2 (±200.6) | -3.1% (±6.6%) | 267,106 | -32.6(±75.1) |
| Switzerland | 2,082 | 2,137 | 2,033 | 1,916 | 1,943 | 1,872 | 2017.0 (±82.7) | 65.0 (±82.7) | 3.2% (±4.0%) | 207,844 | 31.3(±39.8) |

**Supplementary Table 4:**
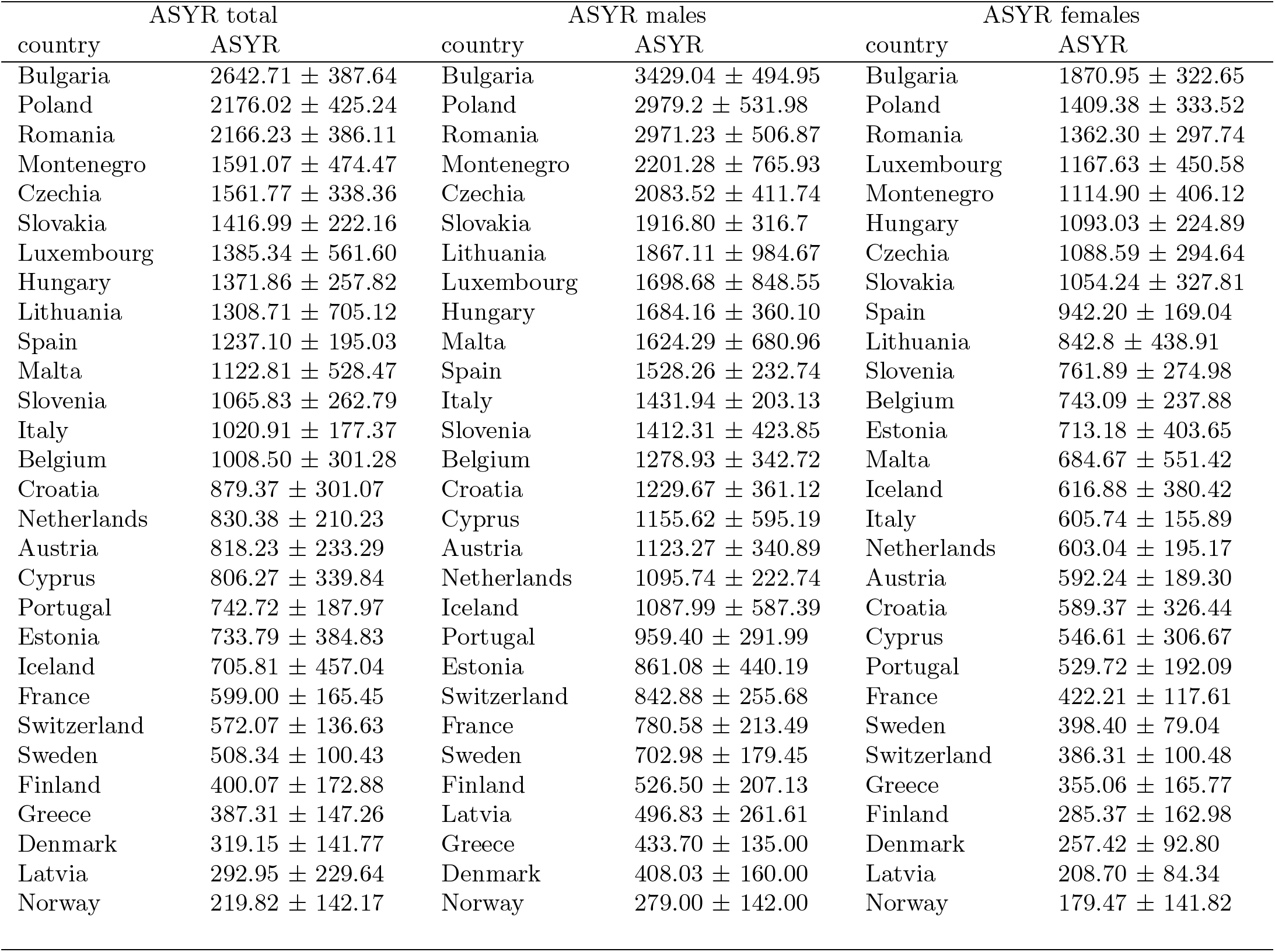
ASYR values for European countries for the year 2020, for ages 0-89.

**Supplementary Table 5:** ASYR values for European countries for the year 2020, for ages 0 to 90+.

| ASYR total |  | ASYR males |  | ASYR females |  |
| --- | --- | --- | --- | --- | --- |
| country | ASYR | country | ASYR | country | ASYR |
| Bulgaria | 2963.08 ± 423.25 | Bulgaria | 3842.60 ± 529.63 | Bulgaria | 2147.98 ± 362.37 |
| Poland | 2676.11 ± 479.93 | Poland | 3700.81 ± 597.50 | Poland | 1823.31 ± 387.70 |
| Romania | 2501.03 ± 430.02 | Romania | 3396.66 ± 546.60 | Romania | 1658.84 ± 344.88 |
| Czechia | 1963.81 ± 389.01 | Czechia | 2612.61 ± 476.53 | Czechia | 1435.77 ± 346.58 |
| Montenegro | 1825.12 ± 530.49 | Lithuania | 2517.08 ± 1058.43 | Luxembourg | 1416.52 ± 505.13 |
| Lithuania | 1789.84 ± 758.74 | Montenegro | 2451.80 ± 866.94 | Montenegro | 1342.87 ± 487.51 |
| Luxembourg | 1704.25 ± 628.43 | Slovenia | 2223.7 ± 534.70 | Hungary | 1326.87 ± 255.44 |
| Slovakia | 1666.51 ± 250.84 | Slovakia | 2213.56 ± 344.59 | Spain | 1288.61 ± 190.40 |
| Spain | 1638.01 ± 220.31 | Luxembourg | 2152.29 ± 961.87 | Slovakia | 1277.82 ± 358.38 |
| Slovenia | 1635.98 ± 320.95 | Spain | 2026.51 ± 275.46 | Slovenia | 1249.83 ± 328.03 |
| Hungary | 1616.83 ± 282.59 | Hungary | 1966.58 ± 380.22 | Lithuania | 1222.45 ± 495.15 |
| Belgium | 1438.89 ± 330.31 | Belgium | 1822.65 ± 392.28 | Belgium | 1120.08 ± 268.13 |
| Malta | 1427.67 ± 577.51 | Malta | 1820.27 ± 747.73 | Malta | 1043.07 ± 606.69 |
| Italy | 1354.28 ± 204.22 | Croatia | 1800.53 ± 448.09 | Croatia | 965.27 ± 368.26 |
| Croatia | 1309.57 ± 352.60 | Netherlands | 1587.07 ± 291.09 | Estonia | 863.81 ± 443.5 |
| Netherlands | 1136.98 ± 238.97 | Austria | 1492.68 ± 402.59 | Portugal | 850.83 ± 238.20 |
| Portugal | 1098.04 ± 245.12 | Italy | 1415.77 ± 200.74 | Netherlands | 816.38 ± 210.52 |
| Austria | 1024.27 ± 276.29 | Portugal | 1384.46 ± 374.22 | Austria | 730.49 ± 231.15 |
| Estonia | 909.88 ± 451.22 | Estonia | 1168.07 ± 571.62 | Iceland | 635.56 ± 436.84 |
| France | 829.05 ± 192.34 | Cyprus | 1142.61 ± 587.28 | France | 605.05 ± 145.93 |
| Cyprus | 796.80 ± 335.12 | France | 1112.10 ± 247.55 | Italy | 601.71 ± 154.81 |
| Iceland | 725.93 ± 507.83 | Iceland | 1109.31 ± 639.07 | Greece | 557.5 ± 228.32 |
| Sweden | 667.81 ± 126.16 | Sweden | 955.58 ± 203.81 | Cyprus | 539.87 ± 303.08 |
| Greece | 604.26 ± 207.60 | Switzerland | 837.75 ± 252.29 | Sweden | 503.02 ± 110.52 |
| Switzerland | 567.55 ± 134.87 | Latvia | 826.88 ± 373.38 | Switzerland | 384.27 ± 99.47 |
| Finland | 476.93 ± 198.87 | Greece | 674.05 ± 198.07 | Latvia | 357.84 ± 152.71 |
| Latvia | 474.65 ± 301.96 | Finland | 651.73 ± 265.71 | Finland | 346.96 ± 182.45 |
| Denmark | 358.89 ± 151.64 | Denmark | 496.97 ± 181.88 | Denmark | 267.38 ± 99.06 |
| Norway | 221.17 ± 146.99 | Norway | 314.10 ± 161.51 | Norway | 177.81 ± 140.37 |

**Supplementary Table 6:** Average PYLL values for European countries for the year 2020.

| PYLL total |  | PYLL males |  | PYLL females |  |
| --- | --- | --- | --- | --- | --- |
| country | PYLL | country | PYLL | country | PYLL |
| Iceland | 29.25 ± 1.22 | Iceland | 23.86 ± 1.99 | Iceland | 39.71 ± 0.13 |
| Luxembourg | 23.08 ± 0.26 | Luxembourg | 19.79 ± 0.37 | Luxembourg | 29.63 ± 2.83 |
| Estonia | 16.37 ± 1.03 | Cyprus | 15.58 ± 0.49 | Estonia | 18.73 ± 1.48 |
| Finland | 15.73 ± 0.77 | Finland | 14.82 ± 0.51 | Finland | 16.65 ± 0.48 |
| Cyprus | 14.48 ± 0.41 | Malta | 13.99 ± 1.09 | Norway | 14.92 ± 0.02 |
| Norway | 13.95 ± 0.39 | Norway | 13.88 ± 0.22 | Cyprus | 13.79 ± 1.04 |
| Poland | 13.87 ± 0.18 | Estonia | 13.58 ± 1.28 | France | 13.76 ± 0.1 |
| Malta | 13.76 ± 1.4 | Poland | 12.96 ± 0.21 | Poland | 13.76 ± 0.19 |
| Bulgaria | 13.46 ± 0.11 | Romania | 12.56 ± 0.11 | Bulgaria | 13.68 ± 0.24 |
| Romania | 13.29 ± 0.01 | Bulgaria | 12.56 ± 0.03 | Greece | 13.63 ± 0.14 |
| Spain | 13.08 ± 0.16 | Spain | 12.4 ± 0.15 | Spain | 13.3 ± 0.14 |
| Lithuania | 13.04 ± 0.82 | Lithuania | 12.14 ± 1.14 | Slovakia | 13.02 ± 0.7 |
| France | 12.77 ± 0.33 | Greece | 12.01 ± 0.38 | Malta | 13 ± 2.19 |
| Greece | 12.68 ± 0.1 | Austria | 11.89 ± 0.68 | Romania | 12.91 ± 0.05 |
| Austria | 12.21 ± 0.35 | Portugal | 11.75 ± 0.53 | Austria | 12.8 ± 0.07 |
| Slovakia | 12.17 ± 0.12 | France | 11.64 ± 0.46 | Lithuania | 12.65 ± 0.21 |
| Hungary | 11.99 ± 0.43 | Italy | 11.46 ± 0.25 | Denmark | 12.44 ± 0.66 |
| Italy | 11.72 ± 0.21 | Hungary | 11.26 ± 0.54 | Montenegro | 12.39 ± 1.07 |
| Portugal | 11.69 ± 0.38 | Belgium | 11.19 ± 0.59 | Hungary | 12 ± 0.37 |
| Czechia | 11.62 ± 0.37 | Slovakia | 11.14 ± 0.43 | Czechia | 11.9 ± 0.3 |
| Montenegro | 11.57 ± 1.08 | Slovenia | 10.95 ± 0.97 | Netherlands | 11.43 ± 0.9 |
| Belgium | 11.15 ± 0.72 | Netherlands | 10.87 ± 0.49 | Italy | 11.15 ± 0.22 |
| Netherlands | 11.14 ± 0.73 | Czechia | 10.8 ± 0.5 | Portugal | 11.08 ± 0.79 |
| Croatia | 11.06 ± 0.23 | Montenegro | 10.64 ± 0.58 | Sweden | 11.03 ± 0.02 |
| Slovenia | 11.05 ± 0.6 | Croatia | 10.56 ± 0.3 | Croatia | 10.97 ± 0.52 |
| Denmark | 10.52 ± 0.68 | Switzerland | 10.42 ± 1.15 | Belgium | 10.74 ± 0.6 |
| Switzerland | 10.32 ± 0.77 | Sweden | 10.36 ± 0.78 | Slovenia | 10.74 ± 0.77 |
| Sweden | 10.29 ± 0.37 | Latvia | 9.63 ± 0.44 | Switzerland | 10.41 ± 0.59 |
| Latvia | 9.71 ± 0.66 | Denmark | 9.58 ± 0.27 | Latvia | 9.47 ± 0.18 |

**Supplementary Table 7:** Average WYLL values for European countries for the year 2020.

| WYLL total |  | WYLL males |  | WYLL females |  |
| --- | --- | --- | --- | --- | --- |
| country | WYLL | country | WYLL | country | WYLL |
| Norway | 38.11 ± 6.23 | Norway | 43.02 ± 0.02 | Denmark | 39.15 ± 0.13 |
| Denmark | 35.38 ± 1.03 | Denmark | 31.69 ± 0.41 | Finland | 35.65 ± 2.69 |
| Finland | 31.87 ± 0.33 | Finland | 31.39 ± 1.39 | Sweden | 29.18 ± 0.4 |
| Slovenia | 21.09 ± 0.92 | Slovakia | 23.4 ± 1.04 | Iceland | 25.56 ± 1.69 |
| Iceland | 20.78 ± 2.31 | Slovenia | 19.81 ± 3.12 | Luxembourg | 24.77 ± 1.09 |
| Slovakia | 18.84 ± 1.04 | Hungary | 17.68 ± 1.01 | Hungary | 19.5 ± 0.06 |
| Malta | 18.47 ± 0.75 | Malta | 17.08 ± 1.6 | Slovenia | 18.89 ± 3.14 |
| Hungary | 18.43 ± 0.31 | Iceland | 15.32 ± 2.22 | Malta | 18.77 ± 2.23 |
| Sweden | 15.57 ± 1.43 | Croatia | 15.07 ± 4.6 | Norway | 17.03 ± 1.79 |
| Czechia | 13.54 ± 0 | Czechia | 12.96 ± 0.56 | Cyprus | 17.03 ± 1.54 |
| Estonia | 12.25 ± 0.9 | Sweden | 12.66 ± 3.65 | Czechia | 15.3 ± 0.47 |
| Poland | 12.04 ± 0.37 | Estonia | 12.37 ± 0.17 | Slovakia | 14.86 ± 4.48 |
| Luxembourg | 11.21 ± 1.06 | Poland | 11.74 ± 0.1 | Netherlands | 14.01 ± 0.47 |
| Netherlands | 10.17 ± 0.24 | Switzerland | 10.86 ± 2.88 | Poland | 13.72 ± 0.89 |
| Switzerland | 10.13 ± 1.81 | Netherlands | 10.26 ± 0.02 | Switzerland | 13.14 ± 1.34 |
| Montenegro | 9.4 ± 2.33 | Montenegro | 9.77 ± 2.16 | Montenegro | 11.59 ± 1.85 |
| Bulgaria | 8.68 ± 0 | Bulgaria | 9.35 ± 0.07 | Estonia | 11.42 ± 0.22 |
| Romania | 8.47 ± 0.58 | Romania | 8.32 ± 0.52 | Portugal | 10.27 ± 1.86 |
| Cyprus | 7.78 ± 0.21 | Luxembourg | 8.27 ± 1.91 | Belgium | 9.77 ± 0.95 |
| Portugal | 7.73 ± 1.52 | Portugal | 7.22 ± 1.75 | Romania | 8.91 ± 0.42 |
| Austria | 6.28 ± 1.16 | Austria | 7.09 ± 1.13 | Austria | 8.52 ± 1.49 |
| Belgium | 6.03 ± 2.23 | Cyprus | 6.98 ± 0.92 | Bulgaria | 7.43 ± 0.28 |
| Croatia | 5.69 ± 1.44 | Belgium | 6.2 ± 1.28 | Greece | 6.46 ± 1.22 |
| Spain | 5.42 ± 0.96 | Spain | 5.3 ± 1.1 | Spain | 6.25 ± 0.6 |
| Lithuania | 5.03 ± 1.97 | Lithuania | 4.97 ± 2.19 | Croatia | 5.89 ± 2.53 |
| Italy | 5 ± 0.18 | Italy | 4.93 ± 0.16 | Lithuania | 5.78 ± 1.57 |
| Greece | 4.61 ± 0.6 | Greece | 3.46 ± 0.32 | Italy | 5.29 ± 0.42 |
| France | 3.04 ± 0.67 | France | 2.5 ± 0 | France | 5 ± 0.91 |
| Latvia | 2.5 ± 0 | Latvia | 2.5 ± 0 | Latvia | 2.5 ± 0 |

**Supplementary Table 8:** Excess mortality in European countries in the year 2020, ages 30-39.

| Country | mortality |  |  |  |  |  |  |  |  |  | Excess mortality per 10<sup>5</sup> |
| --- | --- | --- | --- | --- | --- | --- | --- | --- | --- | --- | --- |
|  | 2020 | 2015 | 2016 | 2017 | 2018 | 2019 | Mean Mortality | Excess Mortality | P-score | Population |  |
| Austria | 600 | 538 | 520 | 510 | 560 | 532 | 538.4 (±14.9) | 61.6 (±14.9) | 11.4% (±3.0%) | 1,217,932 | 5.1(±1.2) |
| Belgium | 878 | 906 | 836 | 861 | 840 | 799 | 864.4 (±30.7) | 13.6 (±30.7) | 1.6% (±3.5%) | 1,499,416 | 0.9(±2.1) |
| Bulgaria | 1,101 | 1,068 | 1,065 | 962 | 1,032 | 978 | 1045.0 (±38.4) | 56.0 (±38.4) | 5.4% (±3.8%) | 956,388 | 5.9(±4.0) |
| Croatia | 343 | 391 | 333 | 343 | 344 | 348 | 354.2 (±17.7) | -11.2 (±17.7) | -3.2% (±4.6%) | 536,215 | -2.1(±3.3) |
| Cyprus | 54 | 60 | 46 | 50 | 58 | 68 | 57.2 (±6.7) | -3.2 (±6.7) | -5.6% (±9.9%) | 143,651 | -2.2(±4.6) |
| Czechia | 1,034 | 1,066 | 983 | 953 | 991 | 941 | 1014.0 (±38.3) | 20.0 (±38.3) | 2.0% (±3.7%) | 1,485,125 | 1.3(±2.6) |
| Denmark | 317 | 304 | 308 | 294 | 336 | 297 | 312.6 (±13.1) | 4.4 (±13.1) | 1.4% (±4.1%) | 686,808 | 0.6(±1.9) |
| Estonia | 201 | 232 | 249 | 191 | 176 | 163 | 203.0 (±28.8) | -2.0 (±28.8) | -1.0% (±12.3%) | 196,352 | -1.0(±14.6) |
| Finland | 462 | 447 | 447 | 440 | 429 | 460 | 457.4 (±8.9) | 4.6 (±8.9) | 1.0% (±1.9%) | 711,550 | 0.6(±1.3) |
| France | 4,967 | 5,047 | 4,843 | 4,824 | 4,954 | 4,927 | 5007.0 (±70.6) | -40.0 (±70.6) | -0.8% (±1.4%) | 8,329,503 | -0.5(±0.9) |
| Greece | 728 | 895 | 820 | 753 | 827 | 709 | 815.2 (±56.4) | -87.2 (±56.4) | -10.7% (±5.8%) | 1,333,233 | -6.5(±4.2) |
| Hungary | 1,005 | 1,120 | 994 | 1,054 | 941 | 917 | 1021.2 (±65.1) | -16.2 (±65.1) | -1.6% (±5.9%) | 1,271,926 | -1.3(±5.1) |
| Iceland | 36 | 22 | 33 | 41 | 37 | 30 | 32.6 (±5.7) | 3.4 (±5.7) | 10.4% (±16.4%) | 53,461 | 6.4(±10.6) |
| Italy | 2,659 | 3,198 | 3,044 | 2,865 | 2,837 | 2,618 | 2976.4 (±172.4) | -317.4 (±172.4) | -10.7% (±4.9%) | 6,854,632 | -4.6(±2.5) |
| Latvia | 355 | 490 | 425 | 480 | 433 | 359 | 451.0 (±40.9) | -96.0 (±40.9) | -21.3% (±6.5%) | 268,173 | -35.8(±15.3) |
| Lithuania | 524 | 730 | 644 | 563 | 582 | 521 | 627.2 (±63.7) | -103.2 (±63.7) | -16.5% (±7.7%) | 359,239 | -28.7(±17.7) |
| Luxembourg | 35 | 34 | 34 | 37 | 36 | 40 | 36.2 (±1.9) | -1.2 (±1.9) | -3.3% (±4.8%) | 98,584 | -1.2(±1.9) |
| Malta | 35 | 31 | 43 | 37 | 37 | 33 | 36.2 (±3.6) | -1.2 (±3.6) | -3.3% (±8.8%) | 88,901 | -1.3(±4.0) |
| Netherlands | 977 | 862 | 858 | 869 | 891 | 867 | 885.4 (±10.1) | 91.6 (±10.1) | 10.3% (±1.2%) | 2,147,931 | 4.3(±0.4) |
| Norway | 332 | 335 | 359 | 303 | 347 | 328 | 342.4 (±16.6) | -10.4 (±16.6) | -3.0% (±4.5%) | 730,547 | -1.4(±2.2) |
| Poland | 6,409 | 5,764 | 5,663 | 5,692 | 5,773 | 5,763 | 5843.0 (±39.3) | 566.0 (±39.3) | 9.7% (±0.7%) | 6,009,458 | 9.4(±0.7) |
| Portugal | 703 | 780 | 685 | 745 | 671 | 670 | 726.2 (±38.9) | -23.2 (±38.9) | -3.2% (±4.9%) | 1,239,016 | -1.9(±3.2) |
| Romania | 2,801 | 2,948 | 2,703 | 2,662 | 2,579 | 2,495 | 2751.8 (±134.1) | 49.2 (±134.1) | 1.8% (±4.7%) | 2,694,828 | 1.8(±5.0) |
| Slovakia | 691 | 731 | 693 | 651 | 647 | 679 | 695.4 (±26.9) | -4.4 (±26.9) | -0.6% (±3.7%) | 847,471 | -0.5(±3.2) |
| Slovenia | 142 | 186 | 173 | 136 | 146 | 162 | 166.2 (±15.8) | -24.2 (±15.8) | -14.6% (±7.4%) | 288,802 | -8.4(±5.5) |
| Spain | 2,538 | 2,740 | 2,593 | 2,551 | 2,415 | 2,316 | 2575.0 (±128.4) | -37.0 (±128.4) | -1.4% (±4.7%) | 6,103,321 | -0.6(±2.1) |
| Sweden | 531 | 558 | 548 | 538 | 567 | 501 | 553.6 (±20.1) | -22.6 (±20.1) | -4.1% (±3.3%) | 1,366,489 | -1.7(±1.5) |
| Switzerland | 447 | 428 | 399 | 445 | 379 | 406 | 418.6 (±20.2) | 28.4 (±20.2) | 6.8% (±4.9%) | 1,229,176 | 2.3(±1.7) |

**Supplementary Table 9:** Excess mortality in European countries in the year 2020, ages 30-39, females.

| Country | mortality |  |  |  |  |  |  |  |  |  | Excess mortality per 10<sup>5</sup> |
| --- | --- | --- | --- | --- | --- | --- | --- | --- | --- | --- | --- |
|  | 2020 | 2015 | 2016 | 2017 | 2018 | 2019 | Mean Mortality | Excess Mortality | P-score | Population |  |
| Austria | 211 | 171 | 165 | 175 | 208 | 156 | 178.2 ($\pm 15.5$) | 32.8 ($\pm 15.5$) | 18.4% ($\pm 9.5\%$) | 601,390 | 5.5($\pm 2.5$) |
| Belgium | 286 | 301 | 271 | 304 | 284 | 265 | 291.4 ($\pm 13.7$) | -5.4 ($\pm 13.7$) | -1.9% ($\pm 4.4\%$) | 749,098 | -0.7($\pm 1.8$) |
| Bulgaria | 309 | 328 | 296 | 293 | 305 | 299 | 313.0 ($\pm 11.0$) | -4.0 ($\pm 11.0$) | -1.3% ($\pm 3.3\%$) | 462,747 | -0.9($\pm 2.4$) |
| Croatia | 101 | 110 | 92 | 97 | 94 | 95 | 98.4 ($\pm 5.6$) | 2.6 ($\pm 5.6$) | 2.6% ($\pm 5.5\%$) | 260,574 | 1.0($\pm 2.1$) |
| Cyprus | 19 | 21 | 10 | 18 | 21 | 24 | 19.6 ($\pm 4.2$) | -0.6 ($\pm 4.2$) | -3.1% ($\pm 17.1\%$) | 72,453 | -0.8($\pm 5.8$) |
| Czechia | 327 | 299 | 332 | 280 | 290 | 299 | 310.4 ($\pm 15.3$) | 16.6 ($\pm 15.3$) | 5.3% ($\pm 4.9\%$) | 717,952 | 2.3($\pm 2.1$) |
| Denmark | 114 | 110 | 105 | 107 | 121 | 104 | 111.0 ($\pm 5.4$) | 3.0 ($\pm 5.4$) | 2.7% ($\pm 4.8\%$) | 337,996 | 0.9($\pm 1.6$) |
| Estonia | 48 | 42 | 74 | 40 | 48 | 51 | 51.0 ($\pm 10.7$) | -3.0 ($\pm 10.7$) | -5.9% ($\pm 16.3\%$) | 93,659 | -3.2($\pm 11.4$) |
| Finland | 135 | 130 | 121 | 123 | 125 | 125 | 126.4 ($\pm 2.6$) | 8.6 ($\pm 2.6$) | 6.8% ($\pm 2.1\%$) | 344,313 | 2.5($\pm 0.8$) |
| France | 1,554 | 1,539 | 1,533 | 1,512 | 1,532 | 1,532 | 1556.8 ($\pm 8.1$) | -2.8 ($\pm 8.1$) | -0.2% ($\pm 0.5\%$) | 4,280,714 | -0.1($\pm 0.2$) |
| Greece | 217 | 257 | 254 | 215 | 228 | 202 | 232.8 ($\pm 18.8$) | -15.8 ($\pm 18.8$) | -6.8% ($\pm 7.0\%$) | 665,385 | -2.4($\pm 2.9$) |
| Hungary | 333 | 334 | 338 | 362 | 285 | 305 | 329.6 ($\pm 23.6$) | 3.4 ($\pm 23.6$) | 1.0% ($\pm 6.7\%$) | 619,906 | 0.5($\pm 3.9$) |
| Iceland | 11 | 4 | 5 | 9 | 12 | 9 | 7.8 ($\pm 2.5$) | 3.2 ($\pm 2.5$) | 41.0% ($\pm 34.2\%$) | 24,653 | 13.0($\pm 10.1$) |
| Italy | 889 | 1,070 | 1,062 | 1,015 | 995 | 888 | 1031.6 ($\pm 57.3$) | -142.6 ($\pm 57.3$) | -13.8% ($\pm 4.6\%$) | 3,402,902 | -4.2($\pm 1.7$) |
| Latvia | 87 | 110 | 84 | 118 | 98 | 84 | 99.6 ($\pm 12.0$) | -12.6 ($\pm 12.0$) | -12.7% ($\pm 9.3\%$) | 130,421 | -9.7($\pm 9.2$) |
| Lithuania | 121 | 172 | 148 | 145 | 135 | 119 | 147.0 ($\pm 15.3$) | -26.0 ($\pm 15.3$) | -17.7% ($\pm 7.7\%$) | 170,267 | -15.3($\pm 9.0$) |
| Luxembourg | 10 | 10 | 12 | 10 | 13 | 13 | 11.6 ($\pm 1.2$) | -1.6 ($\pm 1.2$) | -13.8% ($\pm 8.1\%$) | 48,912 | -3.3($\pm 2.5$) |
| Malta | 10 | 5 | 18 | 12 | 16 | 12 | 12.6 ($\pm 3.9$) | -2.6 ($\pm 3.9$) | -20.6% ($\pm 18.8\%$) | 40,451 | -6.4($\pm 9.6$) |
| Netherlands | 365 | 332 | 359 | 345 | 326 | 330 | 344.8 ($\pm 10.6$) | 20.2 ($\pm 10.6$) | 5.9% ($\pm 3.2\%$) | 1,066,068 | 1.9($\pm 1.0$) |
| Norway | 105 | 119 | 109 | 105 | 103 | 106 | 111.6 ($\pm 4.9$) | -6.6 ($\pm 4.9$) | -5.9% ($\pm 4.0\%$) | 356,127 | -1.9($\pm 1.4$) |
| Poland | 1,443 | 1,235 | 1,298 | 1,292 | 1,306 | 1,231 | 1288.4 ($\pm 28.5$) | 154.6 ($\pm 28.5$) | 12.0% ($\pm 2.4\%$) | 2,952,254 | 5.2($\pm 1.0$) |
| Portugal | 243 | 246 | 254 | 262 | 222 | 252 | 252.0 ($\pm 11.9$) | -9.0 ($\pm 11.9$) | -3.6% ($\pm 4.3\%$) | 637,279 | -1.4($\pm 1.9$) |
| Romania | 819 | 844 | 793 | 795 | 752 | 690 | 799.6 ($\pm 45.1$) | 19.4 ($\pm 45.1$) | 2.4% ($\pm 5.4\%$) | 1,300,038 | 1.5($\pm 3.5$) |
| Slovakia | 183 | 173 | 229 | 180 | 153 | 184 | 187.8 ($\pm 21.9$) | -4.8 ($\pm 21.9$) | -2.6% ($\pm 10.1\%$) | 412,863 | -1.2($\pm 5.3$) |
| Slovenia | 37 | 53 | 46 | 41 | 41 | 44 | 46.6 ($\pm 3.9$) | -9.6 ($\pm 3.9$) | -20.6% ($\pm 6.1\%$) | 135,173 | -7.1($\pm 2.9$) |
| Spain | 876 | 882 | 894 | 853 | 807 | 764 | 849.6 ($\pm 42.4$) | 26.4 ($\pm 42.4$) | 3.1% ($\pm 4.9\%$) | 3,060,796 | 0.9($\pm 1.3$) |
| Sweden | 203 | 182 | 175 | 163 | 184 | 180 | 179.2 ($\pm 6.6$) | 23.8 ($\pm 6.6$) | 13.3% ($\pm 4.0\%$) | 665,413 | 3.6($\pm 1.0$) |
| Switzerland | 160 | 161 | 162 | 156 | 129 | 141 | 152.2 ($\pm 11.2$) | 7.8 ($\pm 11.2$) | 5.1% ($\pm 7.2\%$) | 608,327 | 1.3($\pm 1.8$) |

**Supplementary Table 10:** Excess mortality in European countries in the year 2020, ages 30-39, males.

| Country | mortality |  |  |  |  |  |  |  |  |  | Excess mortality per 10<sup>5</sup> |
| --- | --- | --- | --- | --- | --- | --- | --- | --- | --- | --- | --- |
|  | 2020 | 2015 | 2016 | 2017 | 2018 | 2019 | Mean Mortality | Excess Mortality | P-score | Population |  |
| Austria | 389 | 367 | 355 | 335 | 352 | 376 | 360.2 (±12.3) | 28.8 (±12.3) | 8.0% (±3.6%) | 616,542 | 4.7(±2.0) |
| Belgium | 592 | 605 | 565 | 557 | 556 | 534 | 573.0 (±20.3) | 19.0 (±20.3) | 3.3% (±3.5%) | 750,318 | 2.5(±2.7) |
| Bulgaria | 792 | 740 | 769 | 669 | 727 | 679 | 732.0 (±33.0) | 60.0 (±33.0) | 8.2% (±4.7%) | 493,641 | 12.2(±6.6) |
| Croatia | 242 | 281 | 241 | 246 | 250 | 253 | 255.8 (±12.3) | -13.8 (±12.3) | -5.4% (±4.3%) | 275,641 | -5.0(±4.5) |
| Cyprus | 35 | 39 | 36 | 32 | 37 | 44 | 37.6 (±3.4) | -2.6 (±3.4) | -6.9% (±7.7%) | 71,198 | -3.7(±4.8) |
| Czechia | 707 | 767 | 651 | 673 | 701 | 642 | 703.6 (±39.4) | 3.4 (±39.4) | 0.5% (±5.3%) | 767,173 | 0.4(±5.2) |
| Denmark | 203 | 194 | 203 | 187 | 215 | 193 | 201.6 (±8.5) | 1.4 (±8.5) | 0.7% (±4.1%) | 348,812 | 0.4(±2.4) |
| Estonia | 153 | 190 | 175 | 151 | 128 | 112 | 152.0 (±25.2) | 1.0 (±25.2) | 0.7% (±14.4%) | 102,693 | 1.0(±24.5) |
| Finland | 327 | 317 | 326 | 317 | 304 | 335 | 331.0 (±9.0) | -4.0 (±9.0) | -1.2% (±2.6%) | 367,237 | -1.1(±2.5) |
| France | 3,413 | 3,508 | 3,310 | 3,312 | 3,422 | 3,395 | 3450.2 (±65.0) | -37.2 (±65.0) | -1.1% (±1.8%) | 4,048,789 | -0.9(±1.6) |
| Greece | 511 | 638 | 566 | 538 | 599 | 507 | 582.4 (±40.1) | -71.4 (±40.1) | -12.3% (±5.6%) | 667,848 | -10.7(±6.0) |
| Hungary | 672 | 786 | 656 | 692 | 656 | 612 | 691.6 (±51.4) | -19.6 (±51.4) | -2.8% (±6.8%) | 652,020 | -3.0(±7.9) |
| Iceland | 25 | 18 | 28 | 32 | 25 | 21 | 24.8 (±4.4) | 0.2 (±4.4) | 0.8% (±15.2%) | 28,808 | 0.7(±15.3) |
| Italy | 1,770 | 2,128 | 1,982 | 1,850 | 1,842 | 1,730 | 1944.8 (±119.7) | -174.8 (±119.7) | -9.0% (±5.3%) | 3,451,730 | -5.1(±3.5) |
| Latvia | 268 | 380 | 341 | 362 | 335 | 275 | 351.4 (±31.2) | -83.4 (±31.2) | -23.7% (±6.3%) | 137,752 | -60.5(±22.6) |
| Lithuania | 403 | 558 | 496 | 418 | 447 | 402 | 480.2 (±49.8) | -77.2 (±49.8) | -16.1% (±7.9%) | 188,972 | -40.9(±26.4) |
| Luxembourg | 25 | 24 | 22 | 27 | 23 | 27 | 24.6 (±1.8) | 0.4 (±1.8) | 1.6% (±6.9%) | 49,672 | 0.8(±3.6) |
| Malta | 25 | 26 | 25 | 25 | 21 | 21 | 23.6 (±1.9) | 1.4 (±1.9) | 5.9% (±7.9%) | 48,450 | 2.9(±3.9) |
| Netherlands | 612 | 530 | 499 | 524 | 565 | 537 | 540.6 (±18.7) | 71.4 (±18.7) | 13.2% (±3.8%) | 1,081,863 | 6.6(±1.7) |
| Norway | 227 | 216 | 250 | 198 | 244 | 222 | 230.8 (±16.7) | -3.8 (±16.7) | -1.6% (±6.7%) | 374,420 | -1.0(±4.4) |
| Poland | 4,966 | 4,529 | 4,365 | 4,400 | 4,467 | 4,532 | 4554.6 (±58.9) | 411.4 (±58.9) | 9.0% (±1.4%) | 3,057,204 | 13.5(±1.9) |
| Portugal | 460 | 534 | 431 | 483 | 449 | 418 | 474.2 (±36.6) | -14.2 (±36.6) | -3.0% (±6.9%) | 601,737 | -2.4(±6.1) |
| Romania | 1,982 | 2,104 | 1,910 | 1,867 | 1,827 | 1,805 | 1952.2 (±93.7) | 29.8 (±93.7) | 1.5% (±4.6%) | 1,394,790 | 2.1(±6.8) |
| Slovakia | 508 | 558 | 464 | 471 | 494 | 495 | 507.6 (±29.1) | 0.4 (±29.1) | 0.1% (±5.4%) | 434,608 | 0.1(±6.7) |
| Slovenia | 105 | 133 | 127 | 95 | 105 | 118 | 119.6 (±12.3) | -14.6 (±12.3) | -12.2% (±8.2%) | 153,629 | -9.5(±8.0) |
| Spain | 1,655 | 1,858 | 1,699 | 1,698 | 1,608 | 1,552 | 1725.4 (±91.0) | -70.4 (±91.0) | -4.1% (±4.8%) | 3,042,525 | -2.3(±3.0) |
| Sweden | 328 | 376 | 373 | 375 | 383 | 321 | 374.4 (±19.8) | -46.4 (±19.8) | -12.4% (±4.4%) | 701,076 | -6.6(±2.8) |
| Switzerland | 287 | 267 | 237 | 289 | 250 | 265 | 266.4 (±15.3) | 20.6 (±15.3) | 7.7% (±5.8%) | 620,849 | 3.3(±2.5) |

**Supplementary Table 11:** Excess mortality in European countries in the year 2020, ages 40-64.

| Country | mortality |  |  |  |  |  |  |  |  |  | Excess mortality per 10<sup>5</sup> |
| --- | --- | --- | --- | --- | --- | --- | --- | --- | --- | --- | --- |
|  | 2020 | 2015 | 2016 | 2017 | 2018 | 2019 | Mean Mortality | Excess Mortality | P-score | Population |  |
| Austria | 9,255 | 8,875 | 8,602 | 8,347 | 8,582 | 8,358 | 8718.4 (±169.7) | 536.6 (±169.7) | 6.2% (±2.1%) | 3,149,418 | 17.0(±5.4) |
| Belgium | 12,680 | 12,880 | 12,151 | 11,810 | 11,712 | 11,364 | 12233.8 (±450.3) | 446.2 (±450.3) | 3.6% (±3.6%) | 3,823,073 | 11.7(±11.7) |
| Bulgaria | 19,849 | 17,125 | 16,808 | 16,204 | 16,293 | 15,587 | 16732.2 (±464.3) | 3116.8 (±464.3) | 18.6% (±3.2%) | 2,483,521 | 125.5(±18.7) |
| Croatia | 6,716 | 7,224 | 6,682 | 6,640 | 6,689 | 6,181 | 6830.4 (±289.8) | -114.4 (±289.8) | -1.7% (±4.0%) | 1,411,276 | -8.1(±20.5) |
| Cyprus | 717 | 631 | 575 | 636 | 591 | 625 | 622.0 (±21.1) | 95.0 (±21.1) | 15.3% (±3.8%) | 268,729 | 35.4(±7.8) |
| Czechia | 15,103 | 15,183 | 14,242 | 14,077 | 13,957 | 13,573 | 14502.4 (±469.6) | 600.6 (±469.6) | 4.1% (±3.2%) | 3,754,028 | 16.0(±12.5) |
| Denmark | 5,570 | 6,456 | 6,210 | 5,873 | 5,887 | 5,599 | 6122.6 (±260.5) | -552.6 (±260.5) | -9.0% (±3.7%) | 1,900,863 | -29.1(±13.7) |
| Estonia | 2,350 | 2,344 | 2,248 | 2,182 | 2,178 | 2,045 | 2237.0 (±85.7) | 113.0 (±85.7) | 5.1% (±3.9%) | 439,984 | 25.7(±19.5) |
| Finland | 5,463 | 6,078 | 5,743 | 5,651 | 5,564 | 5,064 | 5740.8 (±287.6) | -277.8 (±287.6) | -4.8% (±4.6%) | 1,745,132 | -15.9(±16.5) |
| France | 69,811 | 72,287 | 69,972 | 68,790 | 67,562 | 65,552 | 70241.4 (±1983.8) | -430.4 (±1983.8) | -0.6% (±2.7%) | 21,517,650 | -2.0(±9.2) |
| Greece | 12,182 | 11,816 | 11,530 | 11,637 | 11,480 | 11,419 | 11798.0 (±122.3) | 384.0 (±122.3) | 3.3% (±1.1%) | 3,797,626 | 10.1(±3.2) |
| Hungary | 22,006 | 23,851 | 22,517 | 21,719 | 21,973 | 20,138 | 22528.4 (±1054.2) | -522.4 (±1054.2) | -2.3% (±4.4%) | 3,474,333 | -15.0(±30.3) |
| Iceland | 268 | 233 | 236 | 233 | 254 | 277 | 253.0 (±15.0) | 15.0 (±15.0) | 5.9% (±5.9%) | 111,401 | 13.5(±13.4) |
| Italy | 57,752 | 53,841 | 51,123 | 51,312 | 50,453 | 49,508 | 52326.6 (±1264.3) | 5425.4 (±1264.3) | 10.4% (±2.6%) | 22,244,774 | 24.4(±5.7) |
| Latvia | 4,543 | 4,947 | 4,680 | 4,688 | 4,737 | 4,346 | 4777.2 (±169.2) | -234.2 (±169.2) | -4.9% (±3.3%) | 653,837 | -35.8(±25.9) |
| Lithuania | 7,464 | 7,520 | 7,158 | 6,356 | 6,265 | 6,197 | 6854.4 (±470.9) | 609.6 (±470.9) | 8.9% (±7.0%) | 985,469 | 61.9(±47.7) |
| Luxembourg | 550 | 482 | 517 | 510 | 515 | 527 | 523.8 (±13.2) | 26.2 (±13.2) | 5.0% (±2.6%) | 217,216 | 12.1(±6.0) |
| Malta | 416 | 389 | 374 | 374 | 396 | 357 | 386.0 (±11.8) | 30.0 (±11.8) | 7.8% (±3.2%) | 160,331 | 18.7(±7.4) |
| Netherlands | 15,969 | 16,399 | 15,905 | 15,460 | 15,357 | 14,892 | 15913.0 (±448.9) | 56.0 (±448.9) | 0.4% (±2.8%) | 5,858,292 | 1.0(±7.6) |
| Norway | 3,727 | 4,062 | 3,981 | 3,819 | 3,803 | 3,724 | 3955.4 (±109.0) | -228.4 (±109.0) | -5.8% (±2.5%) | 1,734,716 | -13.2(±6.3) |
| Poland | 75,954 | 76,731 | 72,326 | 70,737 | 70,473 | 67,728 | 73109.4 (±2596.1) | 2844.6 (±2596.1) | 3.9% (±3.6%) | 12,856,410 | 22.1(±20.2) |
| Portugal | 12,694 | 11,809 | 11,916 | 11,794 | 11,758 | 11,488 | 12013.8 (±125.0) | 680.2 (±125.0) | 5.7% (±1.1%) | 3,736,038 | 18.2(±3.4) |
| Romania | 50,397 | 45,898 | 45,219 | 44,143 | 44,875 | 43,323 | 45661.2 (±778.2) | 4735.8 (±778.2) | 10.4% (±1.9%) | 6,858,653 | 69.0(±11.4) |
| Slovakia | 9,780 | 10,409 | 9,769 | 9,596 | 9,570 | 9,001 | 9849.8 (±395.7) | -69.8 (±395.7) | -0.7% (±3.8%) | 1,918,677 | -3.6(±20.6) |
| Slovenia | 2,486 | 2,739 | 2,565 | 2,599 | 2,456 | 2,301 | 2564.8 (±128.6) | -78.8 (±128.6) | -3.1% (±4.6%) | 756,559 | -10.4(±17.0) |
| Spain | 48,530 | 42,710 | 41,641 | 41,697 | 41,975 | 41,862 | 42803.4 (±337.6) | 5726.6 (±337.6) | 13.4% (±0.9%) | 17,787,666 | 32.2(±1.9) |
| Sweden | 6,581 | 7,263 | 6,687 | 6,606 | 6,405 | 6,064 | 6732.2 (±344.5) | -151.2 (±344.5) | -2.2% (±4.8%) | 3,164,608 | -4.8(±10.9) |
| Switzerland | 6,187 | 6,378 | 6,040 | 6,087 | 6,109 | 5,874 | 6211.2 (±142.5) | -24.2 (±142.5) | -0.4% (±2.2%) | 3,008,509 | -0.8(±4.7) |

**Supplementary Table 12:** Excess mortality in European countries in the year 2020, ages 40-64, females.

| Country | mortality |  |  |  |  |  |  |  |  |  | Excess mortality per 10<sup>5</sup> |
| --- | --- | --- | --- | --- | --- | --- | --- | --- | --- | --- | --- |
|  | 2020 | 2015 | 2016 | 2017 | 2018 | 2019 | Mean Mortality | Excess Mortality | P-score | Population |  |
| Austria | 3,204 | 3,178 | 3,099 | 2,981 | 3,031 | 3,009 | 3121.2 (±62.1) | 82.8 (±62.1) | 2.7% (±2.0%) | 1,582,710 | 5.2(±4.0) |
| Belgium | 4,659 | 4,894 | 4,532 | 4,430 | 4,525 | 4,388 | 4645.0 (±156.7) | 14.0 (±156.7) | 0.3% (±3.3%) | 1,903,120 | 0.7(±8.3) |
| Bulgaria | 6,255 | 5,295 | 5,236 | 5,129 | 5,090 | 4,720 | 5185.2 (±176.0) | 1069.8 (±176.0) | 20.6% (±3.9%) | 1,243,628 | 86.0(±14.2) |
| Croatia | 2,060 | 2,172 | 1,963 | 2,059 | 2,003 | 1,897 | 2059.6 (±81.5) | 0.4 (±81.5) | 0.0% (±3.8%) | 713,346 | 0.1(±11.4) |
| Cyprus | 244 | 227 | 193 | 221 | 222 | 222 | 221.8 (±10.7) | 22.2 (±10.7) | 10.0% (±5.1%) | 136,643 | 16.2(±7.9) |
| Czechia | 4,800 | 4,887 | 4,580 | 4,467 | 4,488 | 4,386 | 4657.6 (±152.5) | 142.4 (±152.5) | 3.1% (±3.3%) | 1,853,549 | 7.7(±8.2) |
| Denmark | 2,173 | 2,519 | 2,444 | 2,220 | 2,250 | 2,121 | 2364.4 (±129.5) | -191.4 (±129.5) | -8.1% (±4.8%) | 949,220 | -20.2(±13.7) |
| Estonia | 702 | 679 | 620 | 626 | 582 | 583 | 626.8 (±31.1) | 75.2 (±31.1) | 12.0% (±5.3%) | 225,466 | 33.4(±13.7) |
| Finland | 1,722 | 1,938 | 1,843 | 1,879 | 1,840 | 1,714 | 1890.0 (±64.3) | -168.0 (±64.3) | -8.9% (±3.0%) | 868,038 | -19.4(±7.5) |
| France | 23,957 | 23,920 | 23,519 | 23,589 | 23,401 | 22,747 | 23899.2 (±337.3) | 57.8 (±337.3) | 0.2% (±1.4%) | 11,012,229 | 0.5(±3.1) |
| Greece | 4,036 | 3,627 | 3,794 | 3,797 | 3,726 | 3,679 | 3795.0 (±57.7) | 241.0 (±57.7) | 6.4% (±1.6%) | 1,965,849 | 12.3(±2.9) |
| Hungary | 7,253 | 8,123 | 7,413 | 7,240 | 7,212 | 6,650 | 7486.0 (±414.9) | -233.0 (±414.9) | -3.1% (±5.1%) | 1,772,947 | -13.1(±23.4) |
| Iceland | 111 | 110 | 107 | 85 | 101 | 107 | 104.4 (±7.9) | 6.6 (±7.9) | 6.3% (±7.5%) | 54,332 | 12.1(±14.6) |
| Italy | 21,010 | 20,143 | 19,100 | 19,191 | 19,001 | 18,772 | 19640.6 (±413.6) | 1369.4 (±413.6) | 7.0% (±2.2%) | 11,306,702 | 12.1(±3.7) |
| Latvia | 1,399 | 1,504 | 1,377 | 1,392 | 1,406 | 1,333 | 1436.8 (±49.4) | -37.8 (±49.4) | -2.6% (±3.3%) | 346,973 | -10.9(±14.2) |
| Lithuania | 2,128 | 2,099 | 1,955 | 1,842 | 1,841 | 1,782 | 1940.6 (±98.6) | 187.4 (±98.6) | 9.7% (±5.3%) | 519,477 | 36.1(±19.0) |
| Luxembourg | 171 | 166 | 171 | 195 | 181 | 184 | 183.4 (±8.9) | -12.4 (±8.9) | -6.8% (±4.3%) | 105,793 | -11.7(±8.4) |
| Malta | 150 | 133 | 134 | 161 | 153 | 128 | 145.0 (±11.2) | 5.0 (±11.2) | 3.4% (±7.4%) | 76,584 | 6.5(±14.7) |
| Netherlands | 6,605 | 7,087 | 6,838 | 6,612 | 6,631 | 6,315 | 6834.2 (±225.0) | -229.2 (±225.0) | -3.4% (±3.0%) | 2,931,284 | -7.8(±7.7) |
| Norway | 1,457 | 1,608 | 1,552 | 1,494 | 1,514 | 1,458 | 1550.8 (±45.1) | -93.8 (±45.1) | -6.0% (±2.7%) | 848,170 | -11.1(±5.4) |
| Poland | 22,178 | 23,099 | 21,517 | 20,973 | 21,181 | 20,375 | 21868.2 (±801.2) | 309.8 (±801.2) | 1.4% (±3.6%) | 6,529,236 | 4.7(±12.3) |
| Portugal | 3,907 | 3,623 | 3,695 | 3,744 | 3,643 | 3,626 | 3755.0 (±40.9) | 152.0 (±40.9) | 4.0% (±1.1%) | 1,975,105 | 7.7(±2.1) |
| Romania | 14,662 | 13,738 | 13,374 | 13,023 | 13,000 | 12,374 | 13395.4 (±396.9) | 1266.6 (±396.9) | 9.5% (±3.2%) | 3,423,435 | 37.0(±11.6) |
| Slovakia | 3,038 | 3,168 | 2,922 | 2,938 | 2,920 | 2,692 | 2972.0 (±132.1) | 66.0 (±132.1) | 2.2% (±4.3%) | 963,841 | 6.8(±13.8) |
| Slovenia | 781 | 822 | 782 | 847 | 792 | 730 | 801.8 (±34.7) | -20.8 (±34.7) | -2.6% (±4.0%) | 367,879 | -5.7(±9.5) |
| Spain | 16,349 | 13,933 | 13,524 | 13,950 | 13,874 | 14,055 | 14146.4 (±158.9) | 2202.6 (±158.9) | 15.6% (±1.3%) | 8,934,587 | 24.7(±1.7) |
| Sweden | 2,574 | 2,897 | 2,695 | 2,701 | 2,600 | 2,584 | 2741.8 (±97.7) | -167.8 (±97.7) | -6.1% (±3.3%) | 1,562,095 | -10.7(±6.2) |
| Switzerland | 2,260 | 2,347 | 2,271 | 2,185 | 2,331 | 2,188 | 2303.6 (±60.0) | -43.6 (±60.0) | -1.9% (±2.5%) | 1,496,087 | -2.9(±4.0) |

**Supplementary Table 13:** Excess mortality in European countries in the year 2020, ages 40-64, males.

| Country | mortality |  |  |  |  |  |  |  |  |  | Excess mortality per 10<sup>5</sup> |
| --- | --- | --- | --- | --- | --- | --- | --- | --- | --- | --- | --- |
|  | 2020 | 2015 | 2016 | 2017 | 2018 | 2019 | Mean Mortality | Excess Mortality | P-score | Population |  |
| Austria | 6,051 | 5,697 | 5,503 | 5,366 | 5,551 | 5,349 | 5597.2 ($\pm 112.2$) | 453.8 ($\pm 112.2$) | 8.1% ($\pm 2.1$%) | 1,566,708 | 29.0($\pm 7.1$) |
| Belgium | 8,021 | 7,986 | 7,619 | 7,380 | 7,187 | 6,976 | 7588.8 ($\pm 306.8$) | 432.2 ($\pm 306.8$) | 5.7% ($\pm 4.1$%) | 1,919,953 | 22.5($\pm 16.0$) |
| Bulgaria | 13,594 | 11,830 | 11,572 | 11,075 | 11,203 | 10,867 | 11547.0 ($\pm 304.2$) | 2047.0 ($\pm 304.2$) | 17.7% ($\pm 3.0$%) | 1,239,893 | 165.1($\pm 24.5$) |
| Croatia | 4,656 | 5,052 | 4,719 | 4,581 | 4,686 | 4,284 | 4770.8 ($\pm 216.6$) | -114.8 ($\pm 216.6$) | -2.4% ($\pm 4.2$%) | 697,930 | -16.4($\pm 31.0$) |
| Cyprus | 473 | 404 | 382 | 415 | 369 | 403 | 400.2 ($\pm 14.6$) | 72.8 ($\pm 14.6$) | 18.2% ($\pm 4.2$%) | 132,086 | 55.1($\pm 11.1$) |
| Czechia | 10,303 | 10,296 | 9,662 | 9,610 | 9,469 | 9,187 | 9844.8 ($\pm 319.9$) | 458.2 ($\pm 319.9$) | 4.7% ($\pm 3.3$%) | 1,900,479 | 24.1($\pm 16.8$) |
| Denmark | 3,397 | 3,937 | 3,766 | 3,653 | 3,637 | 3,478 | 3758.2 ($\pm 133.4$) | -361.2 ($\pm 133.4$) | -9.6% ($\pm 3.1$%) | 951,643 | -38.0($\pm 14.1$) |
| Estonia | 1,648 | 1,665 | 1,628 | 1,556 | 1,596 | 1,462 | 1610.2 ($\pm 61.1$) | 37.8 ($\pm 61.1$) | 2.3% ($\pm 3.7$%) | 214,518 | 17.6($\pm 28.5$) |
| Finland | 3,741 | 4,140 | 3,900 | 3,772 | 3,724 | 3,350 | 3850.8 ($\pm 225.9$) | -109.8 ($\pm 225.9$) | -2.9% ($\pm 5.3$%) | 877,094 | -12.5($\pm 25.7$) |
| France | 45,854 | 48,367 | 46,453 | 45,201 | 44,161 | 42,805 | 46342.2 ($\pm 1673.3$) | -488.2 ($\pm 1673.3$) | -1.1% ($\pm 3.4$%) | 10,505,421 | -4.6($\pm 15.9$) |
| Greece | 8,146 | 8,189 | 7,736 | 7,840 | 7,754 | 7,740 | 8003.0 ($\pm 151.5$) | 143.0 ($\pm 151.5$) | 1.8% ($\pm 1.9$%) | 1,831,777 | 7.8($\pm 8.3$) |
| Hungary | 14,753 | 15,728 | 15,104 | 14,479 | 14,761 | 13,488 | 15042.4 ($\pm 649.0$) | -289.4 ($\pm 649.0$) | -1.9% ($\pm 4.1$%) | 1,701,386 | -17.0($\pm 38.1$) |
| Iceland | 157 | 123 | 129 | 148 | 153 | 170 | 148.6 ($\pm 14.9$) | 8.4 ($\pm 14.9$) | 5.7% ($\pm 9.7$%) | 57,069 | 14.7($\pm 26.1$) |
| Italy | 36,742 | 33,698 | 32,023 | 32,121 | 31,452 | 30,736 | 32686.0 ($\pm 858.5$) | 4056.0 ($\pm 858.5$) | 12.4% ($\pm 2.9$%) | 10,938,072 | 37.1($\pm 7.8$) |
| Latvia | 3,144 | 3,443 | 3,303 | 3,296 | 3,331 | 3,013 | 3340.4 ($\pm 124.7$) | -196.4 ($\pm 124.7$) | -5.9% ($\pm 3.4$%) | 306,864 | -64.0($\pm 40.6$) |
| Lithuania | 5,336 | 5,421 | 5,203 | 4,514 | 4,424 | 4,415 | 4913.8 ($\pm 375.9$) | 422.2 ($\pm 375.9$) | 8.6% ($\pm 7.7$%) | 465,992 | 90.6($\pm 80.7$) |
| Luxembourg | 379 | 316 | 346 | 315 | 334 | 343 | 340.4 ($\pm 11.5$) | 38.6 ($\pm 11.5$) | 11.3% ($\pm 3.6$%) | 111,423 | 34.6($\pm 10.4$) |
| Malta | 266 | 256 | 240 | 213 | 243 | 229 | 241.0 ($\pm 12.6$) | 25.0 ($\pm 12.6$) | 10.4% ($\pm 5.5$%) | 83,747 | 29.9($\pm 15.0$) |
| Netherlands | 9,364 | 9,312 | 9,067 | 8,848 | 8,726 | 8,577 | 9078.8 ($\pm 226.8$) | 285.2 ($\pm 226.8$) | 3.1% ($\pm 2.5$%) | 2,927,008 | 9.7($\pm 7.8$) |
| Norway | 2,270 | 2,454 | 2,429 | 2,325 | 2,289 | 2,266 | 2404.6 ($\pm 66.1$) | -134.6 ($\pm 66.1$) | -5.6% ($\pm 2.5$%) | 886,546 | -15.2($\pm 7.5$) |
| Poland | 53,776 | 53,632 | 50,809 | 49,764 | 49,292 | 47,353 | 51241.2 ($\pm 1807.7$) | 2534.8 ($\pm 1807.7$) | 4.9% ($\pm 3.5$%) | 6,327,174 | 40.1($\pm 28.5$) |
| Portugal | 8,787 | 8,186 | 8,221 | 8,050 | 8,115 | 7,862 | 8258.8 ($\pm 111.2$) | 528.2 ($\pm 111.2$) | 6.4% ($\pm 1.4$%) | 1,760,933 | 30.0($\pm 6.3$) |
| Romania | 35,735 | 32,160 | 31,845 | 31,120 | 31,875 | 30,949 | 32265.8 ($\pm 411.7$) | 3469.2 ($\pm 411.7$) | 10.8% ($\pm 1.4$%) | 3,435,218 | 101.0($\pm 12.0$) |
| Slovakia | 6,742 | 7,241 | 6,847 | 6,658 | 6,650 | 6,309 | 6877.8 ($\pm 266.7$) | -135.8 ($\pm 266.7$) | -2.0% ($\pm 3.6$%) | 954,836 | -14.2($\pm 27.9$) |
| Slovenia | 1,705 | 1,917 | 1,783 | 1,752 | 1,664 | 1,571 | 1763.0 ($\pm 101.9$) | -58.0 ($\pm 101.9$) | -3.3% ($\pm 5.3$%) | 388,680 | -14.9($\pm 26.2$) |
| Spain | 32,187 | 28,777 | 28,117 | 27,747 | 28,101 | 27,807 | 28657.0 ($\pm 320.6$) | 3530.0 ($\pm 320.6$) | 12.3% ($\pm 1.2$%) | 8,853,079 | 39.9($\pm 3.6$) |
| Sweden | 4,007 | 4,366 | 3,992 | 3,905 | 3,805 | 3,480 | 3990.4 ($\pm 251.2$) | 16.6 ($\pm 251.2$) | 0.4% ($\pm 5.9$%) | 1,602,513 | 1.0($\pm 15.7$) |
| Switzerland | 3,927 | 4,031 | 3,769 | 3,902 | 3,778 | 3,686 | 3907.6 ($\pm 105.7$) | 19.4 ($\pm 105.7$) | 0.5% ($\pm 2.7$%) | 1,512,422 | 1.3($\pm 7.0$) |

**Supplementary Table 14:**
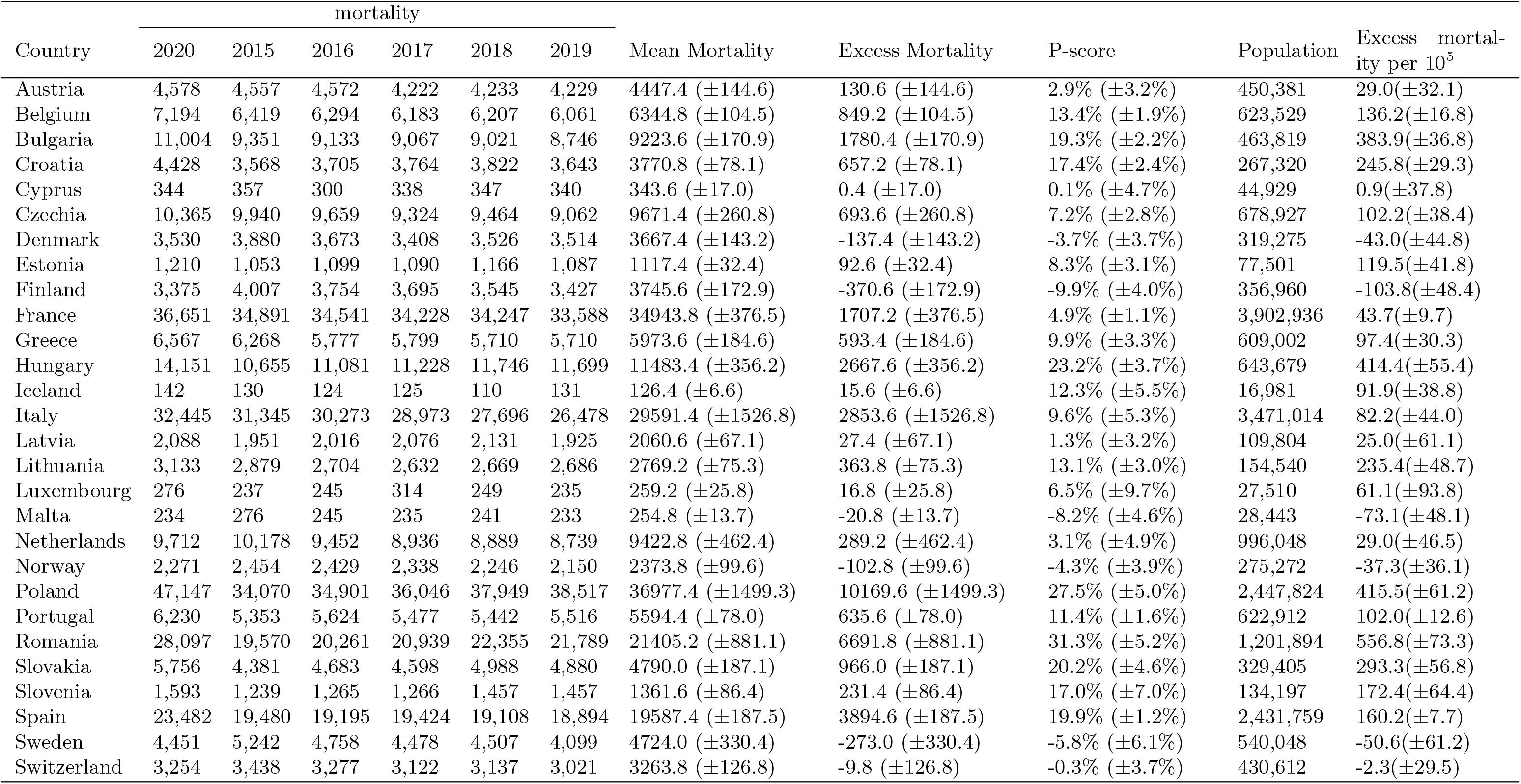
Excess mortality in European countries in the year 2020, ages 65-69.

**Supplementary Table 15:**
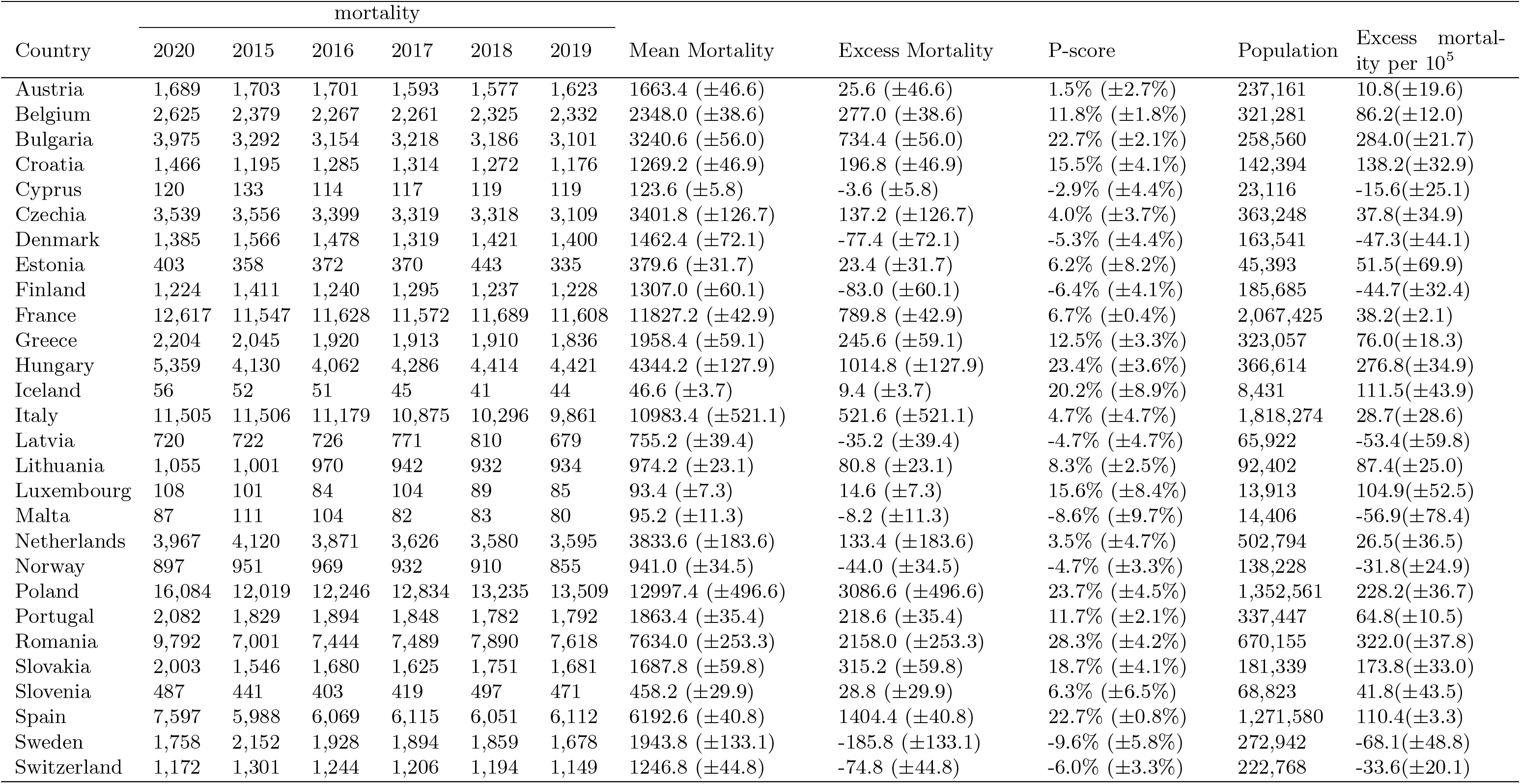
Excess mortality in European countries in the year 2020, ages 65-69, females.

| Country | mortality |  |  |  |  |  |  |  |  |  | Excess mortality per 10<sup>5</sup> |
| --- | --- | --- | --- | --- | --- | --- | --- | --- | --- | --- | --- |
|  | 2020 | 2015 | 2016 | 2017 | 2018 | 2019 | Mean Mortality | Excess Mortality | P-score | Population |  |
| Austria | 1,689 | 1,703 | 1,701 | 1,593 | 1,577 | 1,623 | 1663.4 (±46.6) | 25.6 (±46.6) | 1.5% (±2.7%) | 237,161 | 10.8(±19.6) |
| Belgium | 2,625 | 2,379 | 2,267 | 2,261 | 2,325 | 2,332 | 2348.0 (±38.6) | 277.0 (±38.6) | 11.8% (±1.8%) | 321,281 | 86.2(±12.0) |
| Bulgaria | 3,975 | 3,292 | 3,154 | 3,218 | 3,186 | 3,101 | 3240.6 (±56.0) | 734.4 (±56.0) | 22.7% (±2.1%) | 258,560 | 284.0(±21.7) |
| Croatia | 1,466 | 1,195 | 1,285 | 1,314 | 1,272 | 1,176 | 1269.2 (±46.9) | 196.8 (±46.9) | 15.5% (±4.1%) | 142,394 | 138.2(±32.9) |
| Cyprus | 120 | 133 | 114 | 117 | 119 | 119 | 123.6 (±5.8) | -3.6 (±5.8) | -2.9% (±4.4%) | 23,116 | -15.6(±25.1) |
| Czechia | 3,539 | 3,556 | 3,399 | 3,319 | 3,318 | 3,109 | 3401.8 (±126.7) | 137.2 (±126.7) | 4.0% (±3.7%) | 363,248 | 37.8(±34.9) |
| Denmark | 1,385 | 1,566 | 1,478 | 1,319 | 1,421 | 1,400 | 1462.4 (±72.1) | -77.4 (±72.1) | -5.3% (±4.4%) | 163,541 | -47.3(±44.1) |
| Estonia | 403 | 358 | 372 | 370 | 443 | 335 | 379.6 (±31.7) | 23.4 (±31.7) | 6.2% (±8.2%) | 45,393 | 51.5(±69.9) |
| Finland | 1,224 | 1,411 | 1,240 | 1,295 | 1,237 | 1,228 | 1307.0 (±60.1) | -83.0 (±60.1) | -6.4% (±4.1%) | 185,685 | -44.7(±32.4) |
| France | 12,617 | 11,547 | 11,628 | 11,572 | 11,689 | 11,608 | 11827.2 (±42.9) | 789.8 (±42.9) | 6.7% (±0.4%) | 2,067,425 | 38.2(±2.1) |
| Greece | 2,204 | 2,045 | 1,920 | 1,913 | 1,910 | 1,836 | 1958.4 (±59.1) | 245.6 (±59.1) | 12.5% (±3.3%) | 323,057 | 76.0(±18.3) |
| Hungary | 5,359 | 4,130 | 4,062 | 4,286 | 4,414 | 4,421 | 4344.2 (±127.9) | 1014.8 (±127.9) | 23.4% (±3.6%) | 366,614 | 276.8(±34.9) |
| Iceland | 56 | 52 | 51 | 45 | 41 | 44 | 46.6 (±3.7) | 9.4 (±3.7) | 20.2% (±8.9%) | 8,431 | 111.5(±43.9) |
| Italy | 11,505 | 11,506 | 11,179 | 10,875 | 10,296 | 9,861 | 10983.4 (±521.1) | 521.6 (±521.1) | 4.7% (±4.7%) | 1,818,274 | 28.7(±28.6) |
| Latvia | 720 | 722 | 726 | 771 | 810 | 679 | 755.2 (±39.4) | -35.2 (±39.4) | -4.7% (±4.7%) | 65,922 | -53.4(±59.8) |
| Lithuania | 1,055 | 1,001 | 970 | 942 | 932 | 934 | 974.2 (±23.1) | 80.8 (±23.1) | 8.3% (±2.5%) | 92,402 | 87.4(±25.0) |
| Luxembourg | 108 | 101 | 84 | 104 | 89 | 85 | 93.4 (±7.3) | 14.6 (±7.3) | 15.6% (±8.4%) | 13,913 | 104.9(±52.5) |
| Malta | 87 | 111 | 104 | 82 | 83 | 80 | 95.2 (±11.3) | -8.2 (±11.3) | -8.6% (±9.7%) | 14,406 | -56.9(±78.4) |
| Netherlands | 3,967 | 4,120 | 3,871 | 3,626 | 3,580 | 3,595 | 3833.6 (±183.6) | 133.4 (±183.6) | 3.5% (±4.7%) | 502,794 | 26.5(±36.5) |
| Norway | 897 | 951 | 969 | 932 | 910 | 855 | 941.0 (±34.5) | -44.0 (±34.5) | -4.7% (±3.3%) | 138,228 | -31.8(±24.9) |
| Poland | 16,084 | 12,019 | 12,246 | 12,834 | 13,235 | 13,509 | 12997.4 (±496.6) | 3086.6 (±496.6) | 23.7% (±4.5%) | 1,352,561 | 228.2(±36.7) |
| Portugal | 2,082 | 1,829 | 1,894 | 1,848 | 1,782 | 1,792 | 1863.4 (±35.4) | 218.6 (±35.4) | 11.7% (±2.1%) | 337,447 | 64.8(±10.5) |
| Romania | 9,792 | 7,001 | 7,444 | 7,489 | 7,890 | 7,618 | 7634.0 (±253.3) | 2158.0 (±253.3) | 28.3% (±4.2%) | 670,155 | 322.0(±37.8) |
| Slovakia | 2,003 | 1,546 | 1,680 | 1,625 | 1,751 | 1,681 | 1687.8 (±59.8) | 315.2 (±59.8) | 18.7% (±4.1%) | 181,339 | 173.8(±33.0) |
| Slovenia | 487 | 441 | 403 | 419 | 497 | 471 | 458.2 (±29.9) | 28.8 (±29.9) | 6.3% (±6.5%) | 68,823 | 41.8(±43.5) |
| Spain | 7,597 | 5,988 | 6,069 | 6,115 | 6,051 | 6,112 | 6192.6 (±40.8) | 1404.4 (±40.8) | 22.7% (±0.8%) | 1,271,580 | 110.4(±3.3) |
| Sweden | 1,758 | 2,152 | 1,928 | 1,894 | 1,859 | 1,678 | 1943.8 (±133.1) | -185.8 (±133.1) | -9.6% (±5.8%) | 272,942 | -68.1(±48.8) |
| Switzerland | 1,172 | 1,301 | 1,244 | 1,206 | 1,194 | 1,149 | 1246.8 (±44.8) | -74.8 (±44.8) | -6.0% (±3.3%) | 222,768 | -33.6(±20.1) |

**Supplementary Table 16:** Excess mortality in European countries in the year 2020, ages 65-69, males.

| Country | mortality |  |  |  |  |  |  |  |  |  | Excess mortality per 10<sup>5</sup> |
| --- | --- | --- | --- | --- | --- | --- | --- | --- | --- | --- | --- |
|  | 2020 | 2015 | 2016 | 2017 | 2018 | 2019 | Mean Mortality | Excess Mortality | P-score | Population |  |
| Austria | 2,889 | 2,854 | 2,871 | 2,629 | 2,656 | 2,606 | 2784.0 (±100.8) | 105.0 (±100.8) | 3.8% (±3.7%) | 213,220 | 49.2(±47.3) |
| Belgium | 4,569 | 4,040 | 4,027 | 3,922 | 3,882 | 3,729 | 3996.8 (±99.0) | 572.2 (±99.0) | 14.3% (±2.7%) | 302,248 | 189.3(±32.8) |
| Bulgaria | 7,029 | 6,059 | 5,979 | 5,849 | 5,835 | 5,645 | 5983.0 (±123.9) | 1046.0 (±123.9) | 17.5% (±2.4%) | 205,259 | 509.6(±60.4) |
| Croatia | 2,962 | 2,373 | 2,420 | 2,450 | 2,550 | 2,467 | 2501.6 (±51.3) | 460.4 (±51.3) | 18.4% (±2.4%) | 124,926 | 368.5(±41.1) |
| Cyprus | 224 | 224 | 186 | 221 | 228 | 221 | 220.0 (±13.3) | 4.0 (±13.3) | 1.8% (±5.8%) | 21,813 | 18.3(±61.0) |
| Czechia | 6,826 | 6,384 | 6,260 | 6,005 | 6,146 | 5,953 | 6269.6 (±139.5) | 556.4 (±139.5) | 8.9% (±2.4%) | 315,679 | 176.3(±44.1) |
| Denmark | 2,145 | 2,314 | 2,195 | 2,089 | 2,105 | 2,114 | 2205.0 (±73.4) | -60.0 (±73.4) | -2.7% (±3.2%) | 155,734 | -38.5(±47.1) |
| Estonia | 807 | 695 | 727 | 720 | 723 | 752 | 737.8 (±16.0) | 69.2 (±16.0) | 9.4% (±2.3%) | 32,108 | 215.5(±49.9) |
| Finland | 2,151 | 2,596 | 2,514 | 2,400 | 2,308 | 2,199 | 2438.6 (±124.1) | -287.6 (±124.1) | -11.8% (±4.3%) | 171,275 | -167.9(±72.4) |
| France | 24,034 | 23,344 | 22,913 | 22,656 | 22,558 | 21,980 | 23116.6 (±392.0) | 917.4 (±392.0) | 4.0% (±1.8%) | 1,835,511 | 50.0(±21.3) |
| Greece | 4,363 | 4,223 | 3,857 | 3,886 | 3,800 | 3,874 | 4015.2 (±131.8) | 347.8 (±131.8) | 8.7% (±3.5%) | 285,945 | 121.6(±46.1) |
| Hungary | 8,792 | 6,525 | 7,019 | 6,942 | 7,332 | 7,278 | 7139.2 (±252.5) | 1652.8 (±252.5) | 23.2% (±4.3%) | 277,065 | 596.5(±91.2) |
| Iceland | 86 | 78 | 73 | 80 | 69 | 87 | 79.8 (±5.4) | 6.2 (±5.4) | 7.8% (±6.9%) | 8,550 | 72.5(±63.2) |
| Italy | 20,940 | 19,839 | 19,094 | 18,098 | 17,400 | 16,617 | 18608.0 (±1010.2) | 2332.0 (±1010.2) | 12.5% (±5.8%) | 1,652,740 | 141.1(±61.1) |
| Latvia | 1,368 | 1,229 | 1,290 | 1,305 | 1,321 | 1,246 | 1305.4 (±30.8) | 62.6 (±30.8) | 4.8% (±2.4%) | 43,882 | 142.7(±70.1) |
| Lithuania | 2,078 | 1,878 | 1,734 | 1,690 | 1,737 | 1,752 | 1795.0 (±55.6) | 283.0 (±55.6) | 15.8% (±3.5%) | 62,138 | 455.4(±89.5) |
| Luxembourg | 168 | 136 | 161 | 210 | 160 | 150 | 165.8 (±21.9) | 2.2 (±21.9) | 1.3% (±11.8%) | 13,597 | 16.2(±161.0) |
| Malta | 147 | 165 | 141 | 153 | 158 | 153 | 159.6 (±6.8) | -12.6 (±6.8) | -7.9% (±3.8%) | 14,037 | -89.8(±48.5) |
| Netherlands | 5,745 | 6,058 | 5,581 | 5,310 | 5,309 | 5,144 | 5589.2 (±281.4) | 155.8 (±281.4) | 2.8% (±4.9%) | 493,254 | 31.6(±57.0) |
| Norway | 1,374 | 1,503 | 1,460 | 1,406 | 1,336 | 1,295 | 1432.8 (±67.1) | -58.8 (±67.1) | -4.1% (±4.3%) | 137,044 | -42.9(±49.0) |
| Poland | 31,063 | 22,051 | 22,655 | 23,212 | 24,714 | 25,008 | 23980.0 (±1010.1) | 7083.0 (±1010.1) | 29.5% (±5.2%) | 1,095,263 | 646.7(±92.2) |
| Portugal | 4,148 | 3,524 | 3,730 | 3,629 | 3,660 | 3,724 | 3731.0 (±65.8) | 417.0 (±65.8) | 11.2% (±2.0%) | 285,465 | 146.1(±23.0) |
| Romania | 18,305 | 12,569 | 12,817 | 13,450 | 14,465 | 14,171 | 13771.2 (±646.2) | 4533.8 (±646.2) | 32.9% (±5.9%) | 531,739 | 852.6(±121.6) |
| Slovakia | 3,753 | 2,835 | 3,003 | 2,973 | 3,237 | 3,199 | 3102.2 (±131.0) | 650.8 (±131.0) | 21.0% (±4.9%) | 148,066 | 439.5(±88.5) |
| Slovenia | 1,106 | 798 | 862 | 847 | 960 | 986 | 903.4 (±62.2) | 202.6 (±62.2) | 22.4% (±7.9%) | 65,374 | 309.9(±95.2) |
| Spain | 15,883 | 13,492 | 13,126 | 13,309 | 13,057 | 12,782 | 13394.8 (±209.9) | 2488.2 (±209.9) | 18.6% (±1.9%) | 1,160,179 | 214.5(±18.1) |
| Sweden | 2,693 | 3,090 | 2,830 | 2,584 | 2,648 | 2,421 | 2780.2 (±200.6) | -87.2 (±200.6) | -3.1% (±6.6%) | 267,106 | -32.6(±75.1) |
| Switzerland | 2,082 | 2,137 | 2,033 | 1,916 | 1,943 | 1,872 | 2017.0 (±82.7) | 65.0 (±82.7) | 3.2% (±4.0%) | 207,844 | 31.3(±39.8) |

**Supplementary Table 17:**
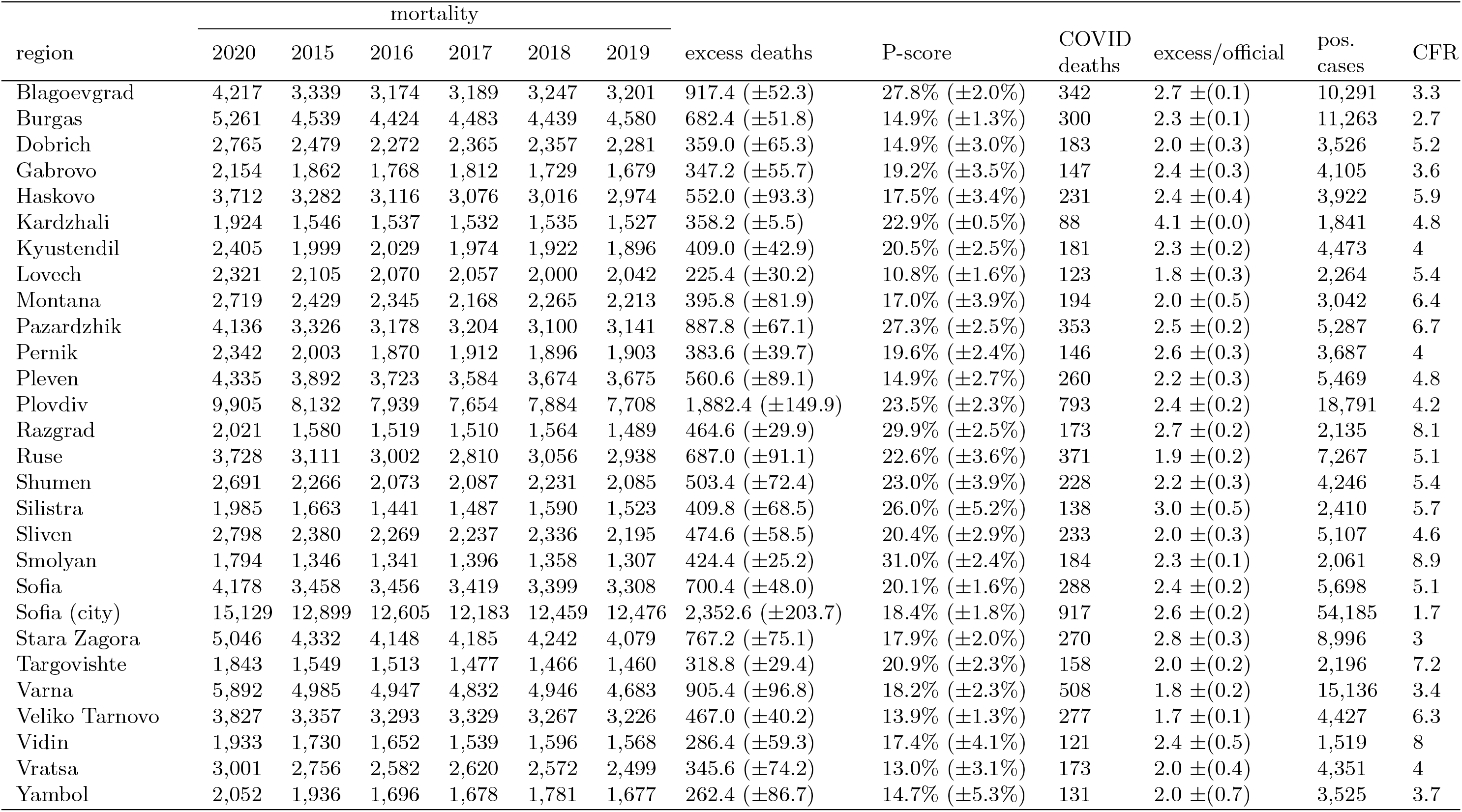
Excess mortality and official COVID-attributed deaths in Bulgarian regions for the year 2020.

**Supplementary Table 18:** Excess mortality in Bulgarian regions for females ages 40-64 for the year 2020.

| region | mortality |  |  |  |  |  | excess deaths | P-score |
| --- | --- | --- | --- | --- | --- | --- | --- | --- |
|  | 2020 | 2015 | 2016 | 2017 | 2018 | 2019 |  |  |
| Blagoevgrad | 281 | 187 | 169 | 181 | 194 | 178 | 96.8 ($\pm 7.4$) | 52.6% ($\pm 5.9\%$) |
| Burgas | 356 | 302 | 300 | 270 | 288 | 299 | 59.4 ($\pm 10.4$) | 20.0% ($\pm 4.0\%$) |
| Dobrich | 207 | 158 | 139 | 158 | 151 | 136 | 54.6 ($\pm 8.2$) | 35.8% ($\pm 6.9\%$) |
| Gabrovo | 106 | 70 | 82 | 94 | 84 | 72 | 24.8 ($\pm 7.6$) | 30.5% ($\pm 11.1\%$) |
| Haskovo | 207 | 172 | 187 | 151 | 141 | 168 | 40.8 ($\pm 14.2$) | 24.5% ($\pm 9.8\%$) |
| Kardzhali | 113 | 89 | 93 | 79 | 73 | 79 | 30.4 ($\pm 6.4$) | 36.8% ($\pm 9.8\%$) |
| Kyustendil | 143 | 113 | 116 | 95 | 102 | 91 | 38.8 ($\pm 8.6$) | 37.2% ($\pm 10.4\%$) |
| Lovech | 120 | 99 | 93 | 105 | 102 | 92 | 20.2 ($\pm 4.4$) | 20.2% ($\pm 5.0\%$) |
| Montana | 140 | 113 | 119 | 124 | 117 | 108 | 23.8 ($\pm 4.7$) | 20.5% ($\pm 4.7\%$) |
| Pazardzhik | 273 | 192 | 183 | 201 | 198 | 196 | 76.6 ($\pm 5.4$) | 39.0% ($\pm 3.7\%$) |
| Pernik | 141 | 121 | 94 | 103 | 99 | 112 | 31.2 ($\pm 8.4$) | 28.4% ($\pm 9.1\%$) |
| Pleven | 217 | 189 | 205 | 181 | 191 | 164 | 27.8 ($\pm 11.7$) | 14.7% ($\pm 6.7\%$) |
| Plovdiv | 561 | 441 | 484 | 428 | 419 | 388 | 122.6 ($\pm 27.4$) | 28.0% ($\pm 7.6\%$) |
| Razgrad | 125 | 103 | 102 | 110 | 77 | 73 | 28.8 ($\pm 13.1$) | 29.9% ($\pm 15.5\%$) |
| Ruse | 185 | 176 | 166 | 160 | 179 | 159 | 13.8 ($\pm 7.2$) | 8.1% ($\pm 4.4\%$) |
| Shumen | 190 | 170 | 133 | 158 | 132 | 127 | 42.0 ($\pm 14.8$) | 28.4% ($\pm 11.7\%$) |
| Silistra | 130 | 97 | 72 | 96 | 85 | 73 | 43.8 ($\pm 9.4$) | 50.8% ($\pm 14.8\%$) |
| Sliven | 204 | 177 | 148 | 175 | 145 | 141 | 42.8 ($\pm 13.6$) | 26.6% ($\pm 9.9\%$) |
| Smolyan | 90 | 90 | 66 | 66 | 78 | 74 | 14.4 ($\pm 7.8$) | 19.0% ($\pm 11.1\%$) |
| Sofia | 215 | 175 | 221 | 182 | 210 | 175 | 19.2 ($\pm 16.8$) | 9.8% ($\pm 8.7\%$) |
| Sofia (stolitsa) | 907 | 871 | 888 | 856 | 884 | 760 | 37.6 ($\pm 41.4$) | 4.3% ($\pm 4.7\%$) |
| Stara Zagora | 292 | 260 | 267 | 243 | 240 | 215 | 43.0 ($\pm 15.9$) | 17.3% ($\pm 7.1\%$) |
| Targovishte | 132 | 89 | 98 | 82 | 98 | 101 | 35.2 ($\pm 6.2$) | 36.4% ($\pm 8.2\%$) |
| Varna | 372 | 306 | 312 | 304 | 304 | 273 | 66.6 ($\pm 12.0$) | 21.8% ($\pm 4.6\%$) |
| Veliko Tarnovo | 180 | 184 | 174 | 190 | 181 | 172 | -3.4 ($\pm 5.8$) | -1.9% ($\pm 3.0\%$) |
| Vidin | 79 | 73 | 77 | 79 | 70 | 83 | 2.6 ($\pm 3.9$) | 3.4% ($\pm 5.0\%$) |
| Vratsa | 170 | 164 | 168 | 165 | 156 | 130 | 10.2 ($\pm 12.2$) | 6.4% ($\pm 7.6\%$) |
| Yambol | 119 | 114 | 80 | 93 | 92 | 81 | 25.4 ($\pm 10.7$) | 27.1% ($\pm 13.0\%$) |

**Supplementary Table 19:** Excess mortality in Bulgarian regions for males ages 40-64 for the year 2020.

| region | mortality |  |  |  |  |  | excess deaths | P-score |
| --- | --- | --- | --- | --- | --- | --- | --- | --- |
|  | 2020 | 2015 | 2016 | 2017 | 2018 | 2019 |  |  |
| Blagoevgrad | 597 | 508 | 476 | 487 | 471 | 451 | 108.8 ($\pm 16.5$) | 22.3% ($\pm 4.0\%$) |
| Burgas | 720 | 679 | 610 | 621 | 633 | 640 | 69.0 ($\pm 20.7$) | 10.6% ($\pm 3.4\%$) |
| Dobrich | 395 | 378 | 356 | 347 | 320 | 327 | 42.2 ($\pm 18.2$) | 12.0% ($\pm 5.5\%$) |
| Gabrovo | 246 | 214 | 180 | 185 | 163 | 191 | 53.0 ($\pm 14.6$) | 27.5% ($\pm 9.0\%$) |
| Haskovo | 477 | 431 | 395 | 392 | 410 | 388 | 61.0 ($\pm 13.8$) | 14.7% ($\pm 3.7\%$) |
| Kardzhali | 228 | 205 | 216 | 201 | 219 | 203 | 13.6 ($\pm 6.4$) | 6.3% ($\pm 3.0\%$) |
| Kyustendil | 311 | 268 | 268 | 258 | 260 | 267 | 41.2 ($\pm 3.8$) | 15.3% ($\pm 1.6\%$) |
| Lovech | 264 | 255 | 273 | 229 | 229 | 227 | 15.0 ($\pm 16.1$) | 6.0% ($\pm 6.4\%$) |
| Montana | 325 | 252 | 308 | 249 | 250 | 260 | 57.2 ($\pm 19.6$) | 21.4% ($\pm 8.3\%$) |
| Pazardzhik | 595 | 480 | 418 | 444 | 416 | 422 | 147.8 ($\pm 21.2$) | 33.1% ($\pm 6.1\%$) |
| Pernik | 305 | 265 | 260 | 256 | 279 | 241 | 40.0 ($\pm 10.9$) | 15.1% ($\pm 4.6\%$) |
| Pleven | 486 | 432 | 424 | 379 | 428 | 409 | 69.2 ($\pm 16.9$) | 16.6% ($\pm 4.5\%$) |
| Plovdiv | 1218 | 1056 | 1069 | 937 | 1052 | 952 | 184.0 ($\pm 49.6$) | 17.8% ($\pm 5.4\%$) |
| Razgrad | 276 | 224 | 225 | 259 | 210 | 196 | 50.8 ($\pm 18.4$) | 22.6% ($\pm 9.3\%$) |
| Ruse | 461 | 372 | 387 | 367 | 368 | 358 | 77.8 ($\pm 8.3$) | 20.3% ($\pm 2.5\%$) |
| Shumen | 361 | 328 | 299 | 260 | 323 | 304 | 53.4 ($\pm 21.0$) | 17.4% ($\pm 7.5\%$) |
| Silistra | 269 | 244 | 207 | 225 | 213 | 207 | 43.4 ($\pm 12.3$) | 19.2% ($\pm 6.1\%$) |
| Sliven | 355 | 292 | 342 | 340 | 308 | 296 | 35.4 ($\pm 18.8$) | 11.1% ($\pm 6.2\%$) |
| Smolyan | 241 | 201 | 172 | 190 | 188 | 195 | 50.2 ($\pm 8.5$) | 26.3% ($\pm 5.4\%$) |
| Sofia | 589 | 463 | 464 | 470 | 459 | 426 | 126.2 ($\pm 13.7$) | 27.3% ($\pm 3.7\%$) |
| Sofia (stolitsa) | 1885 | 1680 | 1550 | 1566 | 1512 | 1516 | 286.6 ($\pm 53.6$) | 17.9% ($\pm 3.8\%$) |
| Stara Zagora | 646 | 534 | 597 | 483 | 536 | 503 | 98.6 ($\pm 33.9$) | 18.0% ($\pm 6.9\%$) |
| Targovishte | 231 | 209 | 199 | 187 | 190 | 199 | 28.6 ($\pm 6.8$) | 14.1% ($\pm 3.7\%$) |
| Varna | 778 | 688 | 698 | 640 | 632 | 631 | 110.6 ($\pm 25.5$) | 16.6% ($\pm 4.3\%$) |
| Veliko Tarnovo | 465 | 417 | 420 | 405 | 404 | 366 | 49.8 ($\pm 16.9$) | 12.0% ($\pm 4.4\%$) |
| Vidin | 202 | 179 | 169 | 161 | 181 | 175 | 25.8 ($\pm 6.4$) | 14.6% ($\pm 4.0\%$) |
| Vratsa | 418 | 355 | 362 | 321 | 310 | 314 | 83.2 ($\pm 19.0$) | 24.9% ($\pm 6.8\%$) |
| Yambol | 250 | 221 | 228 | 216 | 239 | 203 | 24.6 ($\pm 10.5$) | 10.9% ($\pm 4.9\%$) |

**Supplementary Table 20:** Excess mortality in Bulgarian regions for females ages 65-69 for the year 2020.

| region | mortality |  |  |  |  |  | excess deaths | P-score |
| --- | --- | --- | --- | --- | --- | --- | --- | --- |
|  | 2020 | 2015 | 2016 | 2017 | 2018 | 2019 |  |  |
| Blagoevgrad | 157 | 106 | 101 | 97 | 115 | 106 | 48.8 ($\pm 5.3$) | 45.1% ($\pm 6.8\%$) |
| Burgas | 194 | 187 | 154 | 171 | 163 | 171 | 24.0 ($\pm 9.6$) | 14.1% ($\pm 6.1\%$) |
| Dobrich | 123 | 103 | 116 | 91 | 106 | 85 | 22.8 ($\pm 9.6$) | 22.8% ($\pm 10.8\%$) |
| Gabrovo | 70 | 77 | 67 | 71 | 54 | 65 | 3.2 ($\pm 6.7$) | 4.8% ($\pm 9.6\%$) |
| Haskovo | 140 | 124 | 91 | 102 | 102 | 105 | 32.8 ($\pm 9.4$) | 30.6% ($\pm 10.5\%$) |
| Kardzhali | 78 | 51 | 56 | 59 | 48 | 54 | 24.4 ($\pm 3.3$) | 45.5% ($\pm 8.4\%$) |
| Kyustendil | 82 | 66 | 66 | 63 | 69 | 51 | 19.0 ($\pm 5.5$) | 30.2% ($\pm 10.5\%$) |
| Lovech | 63 | 85 | 61 | 52 | 82 | 71 | -9.6 ($\pm 10.9$) | -13.2% ($\pm 11.4\%$) |
| Montana | 100 | 72 | 69 | 81 | 83 | 80 | 22.2 ($\pm 4.8$) | 28.5% ($\pm 7.4\%$) |
| Pazardzhik | 166 | 124 | 113 | 110 | 106 | 110 | 50.2 ($\pm 5.3$) | 43.4% ($\pm 6.3\%$) |
| Pernik | 68 | 62 | 77 | 73 | 68 | 62 | -1.2 ($\pm 5.3$) | -1.7% ($\pm 7.0\%$) |
| Pleven | 149 | 137 | 115 | 150 | 133 | 107 | 17.4 ($\pm 13.6$) | 13.2% ($\pm 10.6\%$) |
| Plovdiv | 331 | 264 | 270 | 281 | 254 | 244 | 64.4 ($\pm 11.2$) | 24.2% ($\pm 5.0\%$) |
| Razgrad | 70 | 60 | 77 | 58 | 54 | 70 | 6.2 ($\pm 7.4$) | 9.7% ($\pm 11.4\%$) |
| Ruse | 120 | 101 | 109 | 103 | 112 | 99 | 12.0 ($\pm 4.3$) | 11.1% ($\pm 4.2\%$) |
| Shumen | 115 | 108 | 77 | 101 | 76 | 82 | 25.4 ($\pm 11.6$) | 28.3% ($\pm 14.7\%$) |
| Silistra | 78 | 73 | 50 | 65 | 56 | 54 | 17.6 ($\pm 7.3$) | 29.1% ($\pm 13.9\%$) |
| Sliven | 145 | 79 | 86 | 77 | 91 | 76 | 62.4 ($\pm 5.1$) | 75.5% ($\pm 10.2\%$) |
| Smolyan | 65 | 46 | 48 | 51 | 47 | 37 | 19.2 ($\pm 4.1$) | 41.9% ($\pm 11.6\%$) |
| Sofia | 164 | 130 | 113 | 119 | 110 | 117 | 40.6 ($\pm 6.0$) | 32.9% ($\pm 6.2\%$) |
| Sofia (stolitsa) | 594 | 509 | 551 | 518 | 509 | 544 | 61.4 ($\pm 15.6$) | 11.5% ($\pm 3.1\%$) |
| Stara Zagora | 178 | 148 | 130 | 167 | 168 | 142 | 26.2 ($\pm 12.9$) | 17.3% ($\pm 9.2\%$) |
| Targovishte | 82 | 46 | 63 | 63 | 62 | 61 | 23.0 ($\pm 5.7$) | 39.0% ($\pm 12.3\%$) |
| Varna | 241 | 164 | 172 | 175 | 201 | 177 | 60.8 ($\pm 10.9$) | 33.7% ($\pm 7.6\%$) |
| Veliko Tarnovo | 158 | 116 | 125 | 119 | 126 | 126 | 32.4 ($\pm 3.6$) | 25.8% ($\pm 3.5\%$) |
| Vidin | 73 | 73 | 72 | 48 | 53 | 57 | 11.6 ($\pm 8.9$) | 18.9% ($\pm 15.1\%$) |
| Vratsa | 109 | 113 | 81 | 97 | 84 | 79 | 15.0 ($\pm 11.1$) | 16.0% ($\pm 12.3\%$) |
| Yambol | 62 | 68 | 44 | 56 | 54 | 69 | 2.2 ($\pm 8.2$) | 3.7% ($\pm 12.5\%$) |

**Supplementary Table 21:** Excess mortality in Bulgarian regions for males ages 65-69 for the year 2020.

| region | mortality |  |  |  |  |  | excess deaths | P-score |
| --- | --- | --- | --- | --- | --- | --- | --- | --- |
|  | 2020 | 2015 | 2016 | 2017 | 2018 | 2019 |  |  |
| Blagoevgrad | 287 | 198 | 207 | 218 | 253 | 198 | 65.0 ($\pm 18.0$) | 29.3% ($\pm 9.7\%$) |
| Burgas | 355 | 308 | 332 | 334 | 324 | 299 | 30.8 ($\pm 12.0$) | 9.5% ($\pm 3.9\%$) |
| Dobrich | 186 | 196 | 192 | 189 | 185 | 190 | -10.0 ($\pm 3.2$) | -5.1% ($\pm 1.5\%$) |
| Gabrovo | 122 | 116 | 120 | 120 | 116 | 100 | 6.0 ($\pm 6.5$) | 5.2% ($\pm 5.6\%$) |
| Haskovo | 249 | 218 | 216 | 239 | 196 | 212 | 26.4 ($\pm 12.1$) | 11.9% ($\pm 5.8\%$) |
| Kardzhali | 135 | 96 | 100 | 89 | 96 | 108 | 34.0 ($\pm 5.4$) | 33.7% ($\pm 6.8\%$) |
| Kyustendil | 170 | 136 | 151 | 145 | 126 | 142 | 26.8 ($\pm 7.5$) | 18.7% ($\pm 5.9\%$) |
| Lovech | 133 | 129 | 129 | 131 | 127 | 108 | 5.8 ($\pm 7.5$) | 4.6% ($\pm 5.9\%$) |
| Montana | 201 | 178 | 156 | 151 | 157 | 134 | 44.2 ($\pm 12.4$) | 28.2% ($\pm 9.4\%$) |
| Pazardzhik | 271 | 247 | 223 | 211 | 218 | 218 | 42.8 ($\pm 10.9$) | 18.8% ($\pm 5.5\%$) |
| Pernik | 153 | 138 | 125 | 125 | 107 | 122 | 26.4 ($\pm 8.7$) | 20.9% ($\pm 7.8\%$) |
| Pleven | 270 | 261 | 226 | 247 | 264 | 225 | 19.0 ($\pm 14.6$) | 7.6% ($\pm 5.9\%$) |
| Plovdiv | 629 | 486 | 514 | 525 | 472 | 493 | 120.6 ($\pm 16.7$) | 23.7% ($\pm 3.9\%$) |
| Razgrad | 165 | 124 | 117 | 100 | 120 | 131 | 44.2 ($\pm 9.0$) | 36.6% ($\pm 9.5\%$) |
| Ruse | 237 | 215 | 194 | 169 | 237 | 199 | 31.8 ($\pm 19.8$) | 15.5% ($\pm 10.2\%$) |
| Shumen | 179 | 157 | 155 | 157 | 148 | 164 | 22.0 ($\pm 4.5$) | 14.0% ($\pm 3.2\%$) |
| Silistra | 137 | 119 | 111 | 116 | 107 | 106 | 22.8 ($\pm 4.4$) | 20.0% ($\pm 4.5\%$) |
| Sliven | 200 | 175 | 170 | 160 | 190 | 145 | 29.6 ($\pm 13.1$) | 17.4% ($\pm 8.4\%$) |
| Smolyan | 118 | 83 | 107 | 104 | 109 | 91 | 18.4 ($\pm 8.9$) | 18.5% ($\pm 9.7\%$) |
| Sofia | 299 | 242 | 250 | 261 | 230 | 206 | 59.6 ($\pm 16.6$) | 24.9% ($\pm 8.1\%$) |
| Sofia (stolitsa) | 951 | 830 | 823 | 733 | 790 | 769 | 147.6 ($\pm 31.3$) | 18.4% ($\pm 4.5\%$) |
| Stara Zagora | 309 | 274 | 297 | 261 | 273 | 277 | 29.4 ($\pm 10.3$) | 10.5% ($\pm 3.9\%$) |
| Targovishte | 130 | 113 | 122 | 110 | 72 | 84 | 26.6 ($\pm 16.6$) | 25.7% ($\pm 17.4\%$) |
| Varna | 424 | 358 | 327 | 312 | 334 | 341 | 84.0 ($\pm 13.3$) | 24.7% ($\pm 4.7\%$) |
| Veliko Tarnovo | 262 | 224 | 213 | 218 | 209 | 201 | 45.8 ($\pm 6.8$) | 21.2% ($\pm 3.7\%$) |
| Vidin | 115 | 123 | 103 | 101 | 85 | 86 | 13.0 ($\pm 12.2$) | 12.7% ($\pm 12.0\%$) |
| Vratsa | 203 | 189 | 188 | 192 | 171 | 174 | 17.8 ($\pm 7.5$) | 9.6% ($\pm 4.3\%$) |
| Yambol | 139 | 126 | 111 | 131 | 119 | 122 | 15.6 ($\pm 5.9$) | 12.6% ($\pm 5.1\%$) |

## Notes

### Competing Interest Statement

The authors have declared no competing interest.

### Funding Statement

This work was not directly funded by a third party

### Author Declarations

No IRB approval was needed for this research

